# Antithrombin, protein C and protein S: Genome and transcriptome wide association studies identify 7 novel loci regulating plasma levels

**DOI:** 10.1101/2022.11.01.22281689

**Authors:** Yuekai Ji, Gerard Temprano-Sagrera, Lori A Holle, Allison Bebo, Jennifer Brody, Ngoc-Quynh Le, Michael R Brown, Angel Martinez-Perez, Colleen M Sitlani, Pierre Suchon, Marcus E Kleber, David B Emmert, Ayse Bilge Ozel, Dre’Von A Dobson, Weihong Tang, Dolors Llobet, Russell P Tracy, Jean-François Deleuze, Graciela E Delgado, Martin Gögele, Kerri L Wiggins, Juan Carlos Souto, James S Pankow, Kent D Taylor, David-Alexandre Trégouët, Angela P Moissl, Christian Fuchsberger, Frits R Rosendaal, Alanna C Morrison, Jose Manuel Soria, Mary Cushman, Pierre-Emmanuel Morange, Winfried März, Andrew A Hicks, Karl C Desch, Andrew D Johnson, Paul S de Vries, CHARGE Consortium Hemostasis Working Group, INVENT Consortium, Alisa S Wolberg, Nicholas L Smith, Maria Sabater-Lleal

**Affiliations:** Cardiovascular Division, Department of Medicine, University of Minnesota, MN, USA; Unit of genomics of Complex Disease, Institut d’Investigació Biomedica Sant Pau (IIB SANT PAU), Barcelona, Spain; Department of Pathology and Laboratory Medicine and UNC Blood Research Center, University of North Carolina at Chapel Hill, NC, USA; Human Genetics Center, Department of Epidemiology, Human Genetics, and Environmental Sciences, School of Public Health, The University of Texas Health Science Center at Houston, TX, USA; Department of Medicine, University of Washington, WA, USA; Cardiovascular Health Research Unit, Department of Medicine, University of Washington, WA, USA; C2VN, INSERM, INRAE, Aix Marseille Univ, France; Laboratory of Haematology, La Timone Hospital, France; SYNLAB MVZ für Humangenetik Mannheim, Germany; Vth Department of Medicine, Medical Faculty Mannheim, Heidelberg University, Germany; Institute for Biomedicine (affiliated to the University of Lübeck), Eurac Research, Italy; Department of Human Genetics, University of Michigan, C.S. Mott Children’s Hospital, MI, USA; Division of Epidemiology and Community Health, School of Public Health, University of Minnesota, MN, USA; Unit of Thrombosis and Hemostasis, Hospital de la Santa Creu i Sant Pau, Barcelona, Spain; Department of Pathology and Laboratory Medicine, University of Vermont College of Medicine, VT, USA; Centre National de Recherche en Génomique Humaine, CEA, France; Centre d’Etude du Polymorphisme Humain, Fondation Jean Dausset, France; Laboratory of Excellence on Medical Genomics (GenMed), France; The Institute for Translational Genomics and Population Sciences, Department of Pediatrics, The Lundquist Institute for Biomedical Innovation at Harbor-UCLA Medical Center, CA, USA; INSERM UMR 1219, Bordeaux Population Health Research Center, France; Institute of Nutritional Sciences, Friedrich Schiller University Jena, Germany; Competence Cluster for Nutrition and Cardiovascluar Health(nutriCARD) Halle-Jena-Leipzig, Germany; Department of Clinical Epidemiology, Leiden University Medical Center, the Netherlands; Larner College of Medicine, University of Vermont, VT, USA; Synlab Academy, Synlab Holding Deutschland GmbH, Germany; Department of Pediatrics, University of Michigan, C.S. Mott Children’s Hospital, MI, USA; National Heart Lung and Blood Institute, Division of Intramural Research, Population Sciences Branch, The Framingham Heart Study, MA, USA; Department of Epidemiology, University of Washington, WA, USA; Kaiser Permanente Washington Health Research Institute, Kaiser Permanente, WA, USA; Seattle Epidemiologic Research and Information Center, Department of Veterans Affairs Office of Research and Development, WA, USA; Cardiovascular Medicine Unit, Department of Medicine, Karolinska Institutet, Center for Molecular Medicine, Stockholm, Sweden

**Author notes:** These authors contributed equally as first authors (Yuekai Ji, Gerard Temprano-Sagrera). These authors contributed equally as last authors (Nicholas L Smith, Maria Sabater-Lleal). Corresponding authors: Maria Sabater-Lleal, PhD, Genomics of Complex Disease Unit, Sant Pau Biomedical Research Institute (IIB Sant Pau), St Quintí 77-79, 08041, Barcelona, Nicholas L. Smith, PhD, Cardiovascular Health Research Unit, University of Washington, 1730 Minor Ave, Suite 1360, Seattle WA 98101.

## Abstract

**Objective:** Antithrombin, protein C (PC) and protein S (PS) are circulating natural-anticoagulant proteins that regulate hemostasis and of which partial deficiencies are causes of venous thromboembolism. Previous genetic association studies involving antithrombin, PC, and PS were limited by modest sample sizes or by being restricted to candidate genes. In the setting of the Cohorts for Heart and Aging Research in Genomic Epidemiology consortium, we meta-analyzed across ancestries the results from 10 genome-wide association studies (GWAS) of plasma levels of antithrombin, PC, PS free and PS total.

**Approach and Results:** Study participants were of European and African ancestries and genotype data were imputed to TOPMed, a dense multi-ancestry reference panel. Each of 10 studies conducted a GWAS for each phenotype and summary results were meta-analyzed, stratified by ancestry. We also conducted transcriptome-wide association analyses and multi-phenotype analysis to discover additional associations. Novel GWAS findings were validated by *in vitro* functional experiments. Mendelian randomization was performed to assess the causal relationship between these proteins and cardiovascular outcomes.

GWAS meta-analyses identified 4 newly associated loci: 3 with antithrombin levels (*GCKR, BAZ1B*, and *HP-TXNL4B*) and 1 with PS levels (*ORM1*-*ORM2*). TWAS identified 3 newly associated genes: 1 with antithrombin level (*FCGRT*), 1 with PC (*GOLM2*), and 1 with PS (*MYL7*). In addition, we replicated 7 independent loci reported in previous studies. Functional experiments provided evidence for the involvement of *GCKR, SNX17*, and *HP* genes in antithrombin regulation.

**Conclusion:** The use of larger sample sizes, diverse populations, and a denser imputation reference panel allowed the detection of 7 novel genomic loci associated with plasma antithrombin, PC, and PS levels.

## INTRODUCTION

Antithrombin, protein C (PC), and protein S (PS) are circulating anticoagulant proteins, and low levels or low activity of these proteins are associated with the risk of venous thromboembolism (VTE)^1-5^. Variation in the protein-coding genes for antithrombin, PC, and PS (*SERPINC1, PROC*, and *PROS1*, respectively)^6-8^ has been studied for decades, and rare mutations have been associated both with low protein levels and with risk of VTE^6,9-12^. There have been at least 6 agnostic genome-wide association studies (GWAS) for antithrombin, PC, and PS, with sample sizes ranging from 351 (GAIT, antithrombin) to 13,968 (ARIC, PC). For antithrombin, no additional genome-wide significant loci beyond *SERPINC1* were identified^13,14^. For PC, significant loci at the *GCKR* and *BAZ1B* genes had been identified in European ancestry (EA) populations^15,16^, and the *CELSR2-PSRC1-SORT1, PROC* and *PROCR* loci were identified in both EA and African ancestry (AA) populations^14,16-18^. For PS, no genome-wide significant associations have been found. In this report, using larger sample sizes, diverse populations, and a denser imputation reference panel, we sought to identify novel genomic loci associated with plasma antithrombin, PC, and PS levels.

## METHODS

### Overview

We used densely imputed genotypes to perform cross-ancestry (antithrombin and PC) and EA-only (PS) GWAS meta-analyses and attempted replication of the lead variants using available summary data from a proteomics-based study^19^. This was followed by a multi-phenotype analysis and transcriptome-wide association analyses (TWAS) in EA individuals. For characterization and prioritization of genes, we used colocalization and fine-mapping analyses, and novel GWAS findings were functionally interrogated. Last, we conducted Mendelian randomization (MR) analyses to assess causal relationships with cardiovascular clinical events. **Figure 1** is a schematic summarizing our approach.

**Figure 1.**
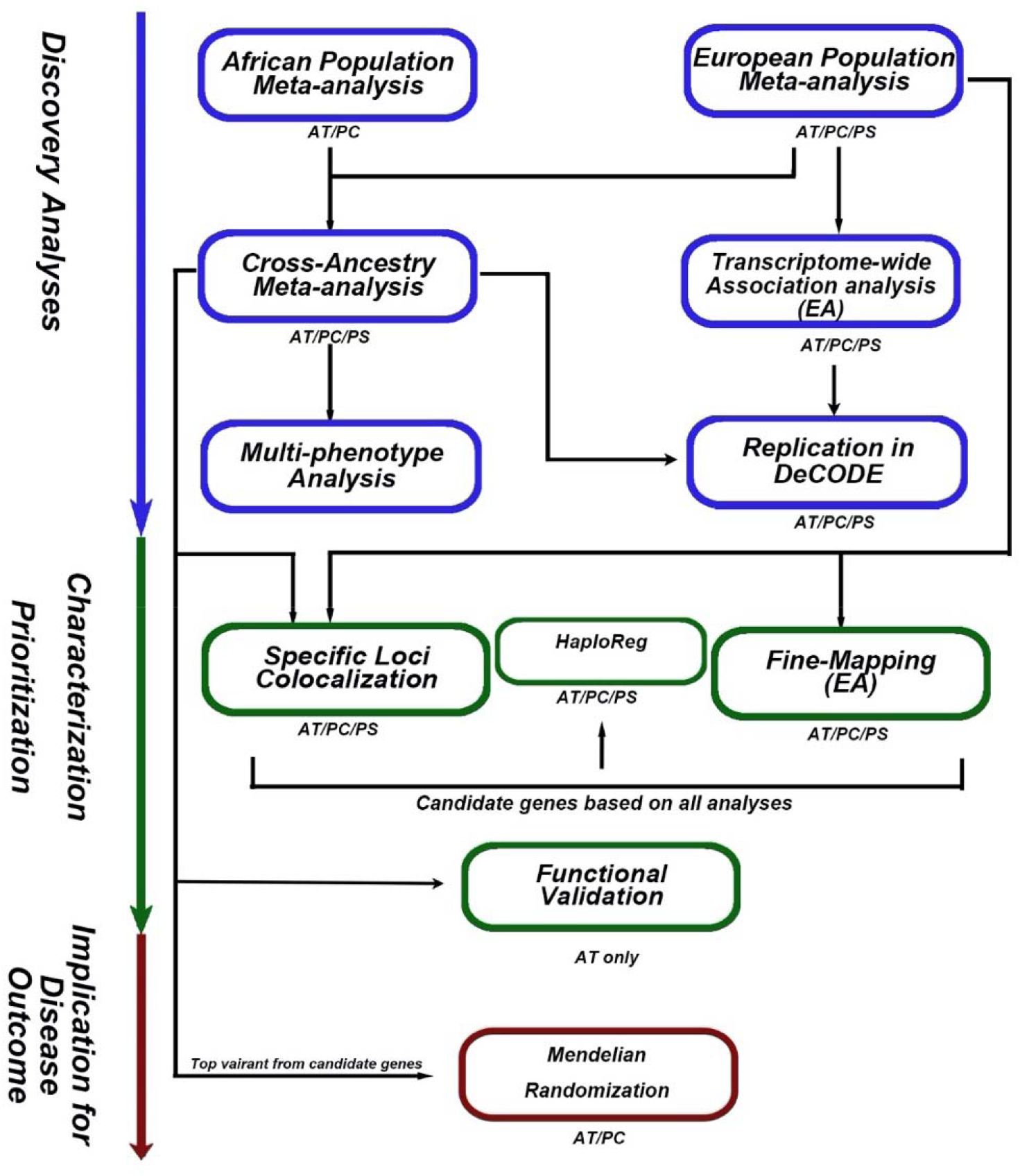
Schematic view of the analysis’s workflow

### Study Design and Participating Studies

The setting for the meta-analysis is the Cohorts for Heart and Aging Research in Genomic Epidemiology (CHARGE) Consortium Hemostasis Working Group^20^. We included data from 10 studies from the US and Europe that measured 1 or more of the 3 natural anticoagulants in plasma, by antigen or activity methods. Study details including genotype and phenotype measurement, study design, population, and baseline time are found in **Supplemental Tables S1, S2**, and **Supplementary Materials**^**14,16,21-31**^. In total, 27,606 EA and 2,688 AA participants were included. All studies were approved by appropriate research ethics committees and all participants provided informed consent.

### Discovery Analysis

#### Study-Specific Genome-Wide Association Analyses

Each study imputed measured genotypes to the Trans-Omic for Precision Medicine (TOPMed) reference panel before association analyses^32^. Study-specific quality control was implemented before the analysis. Details about genotyping platforms and specific quality control parameters can be found in **Supplementary Table S2**. Each study followed a common analysis plan that required performing linear regression within each ancestry group, adjusting for sex, age, principal components, and study-specific variables, which included a kinship matrix when necessary to account for family structure. Residuals from regression were inverse-normal transformed and were re-scaled by the standard deviation (SD) of the pre-transformed values for antithrombin and PS. Because different studies had different unit measures for PC, we did not re-scale by SD, and used the inverse-normal transformed levels for the PC analyses. Details of the measures of the 3 natural anticoagulants can be found in **Supplemental Table S2**. Associations with imputed genotypes were then tested using an additive genetic model between each imputed dosage and the residuals for each re-scaled (antithrombin, PS) or inverse-normal transformed (PC) phenotype using linear regression and adjusting for all the covariates used in 6 the phenotype regression. The X chromosome was additionally stratified by sex where women and men were coded as 0, 1, 2 and 0, 2, respectively.

#### Population-Specific and Cross-Ancestry Meta-Analysis

Quality control across studies was conducted using EasyQC^33^. Details of meta-analysis quality control can be found in **Supplementary Materials**. We meta-analyzed study-level summary results, first by phenotype measure (antigen or activity), then by ancestry. Only variants appearing in at least 2 cohorts were retained in the final meta-analyses. Cross-ancestry meta-analyses were conducted on those phenotypes that included EA and AA participants (AT and PC). Meta-analyses were performed by 2 analysts in parallel.

The significance threshold^34^ was set at 5 × 10^−9^. A locus was defined as 1 Mb upstream and downstream of the variant with the lowest p-value. Genome-wide significant variants with MAF < 1%, present in 2 cohorts or less, or with inconsistent beta directions between cohorts were not considered.

#### Conditional Analysis

We performed approximate conditional and joint analyses for all variants with MAF > 1% using summary statistics from ancestry-specific meta-analyses using COJO (Conditional & Joint; gcta—cojo--slct)^35^, implemented in the Genome-Wide Complex Trait Analysis (GCTA) software^36^, to identify additional independent signals at the associated loci.

#### Replication

We sought for replication of associations for the identified lead variants in an external dataset, using available summary data from DeCODE Genetics (available at https://www.decode.com/summarydata/)^19^. DeCODE Genetics used the SOMAscan multiplexed proteomics assay to obtain proteomic measurements on 35,559 individuals of Icelandic origin, for which antithrombin, PC and PS data is available. The significance threshold p-value of the replication cohort was set at 4.2 × 10^−3^, after correcting for the number of identified lead variants (n = 12; 0.05/12 = 4.2 × 10^−3^).

#### Transcriptome-Wide Association Analyses

We used GWAS results and S-PrediXcan and S-MultiXcan^37,38^ to perform transcriptome-wide analyses for each phenotype within the EA populations in order to infer significant associations between the *cis* component of gene expression and the phenotypes. See detailed methods in **Supplementary methods**. Only tissues with a potential role in the synthesis or regulation of anticoagulants proteins (artery aorta, artery coronary, artery tibial, liver and whole blood) were considered to reduce false positives from more distally related tissues. The significance threshold was established as a Bonferroni correction to the number of genes interrogated: up to 66,745 genes/0.05 = 7.5 × 10^−7^.

#### Multi-Phenotype Meta-Analysis

We jointly analyzed the 4 meta-analyses results (cross-ancestry meta-analyses for antithrombin and PC, and the 2 EA PS meta-analyses) using a multi-phenotype method implemented in the metaUSAT R package 1.17^39^. Significant multi-phenotype associations were defined as any genome-wide significant lead variants in the multivariate analysis (p-values_multivariate_ for the lead variant < 5 × 10^−9^), that were also nominally significant in a least 2 of the phenotypes individually (p-value_univariate_ < 0.005)^40^. Additionally, we considered novel variants to be those that were not genome-wide significant for any of the 4 phenotypes individually, or that had not been associated with antithrombin, PC, PS free or total in a previous GWAS for antithrombin, PC, PS free or total. Lead variants for each phenotype found in the discovery (**Table 1**) were queried using the HaploR R package v4.0.6 to extract functional annotations and biological information (**Table 1 and Supplementary Table S3**). Further details are reported in **Supplementary Methods**.

**Table 1.**
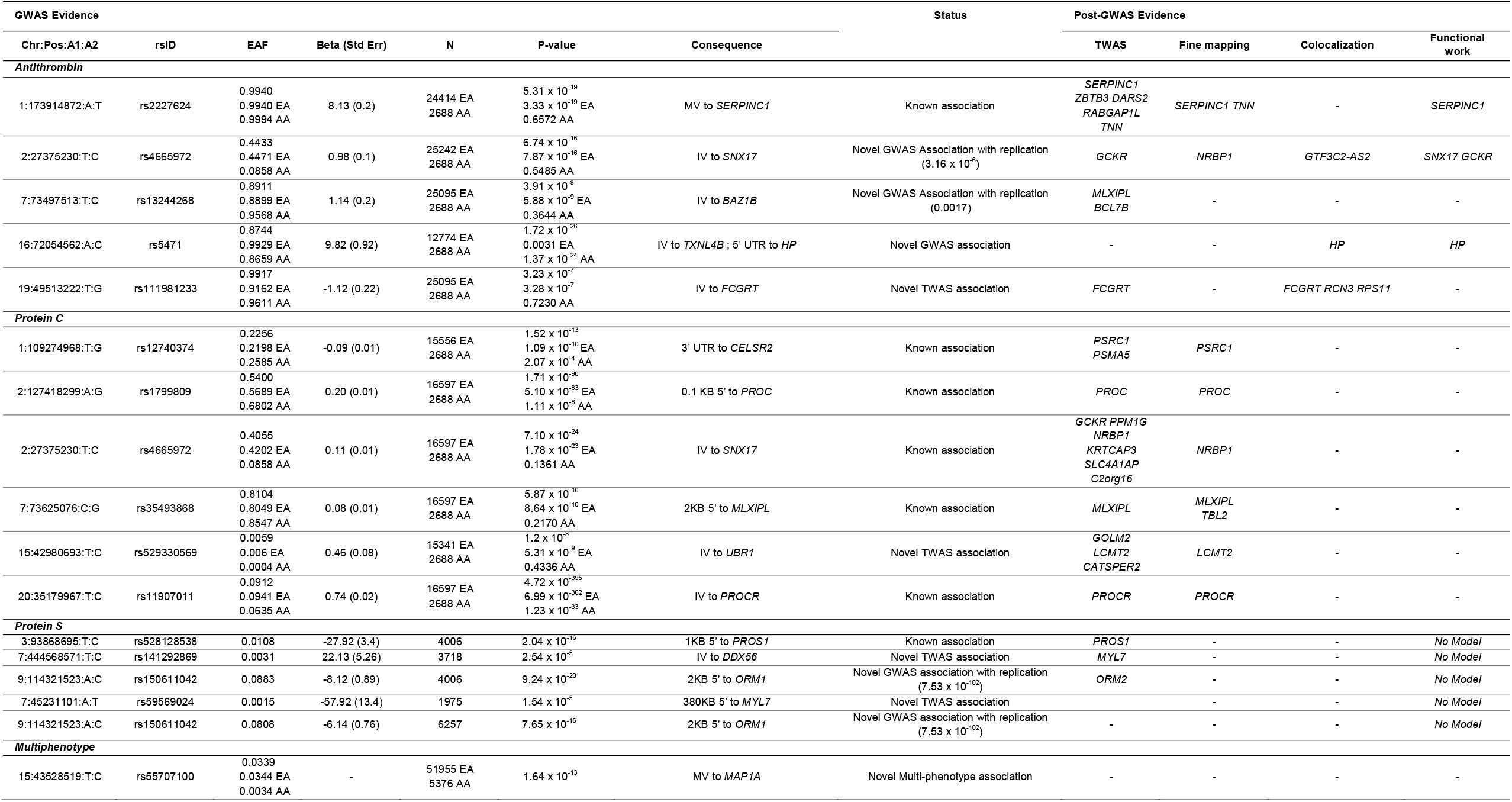
GWAS and post-GWAS evidence of candidate genes. A1: Effect Allele; A2: Other Allele; AA: African ancestry; EA: European ancestry; IV: Intronic Variant; MV: missense variant; 3’UTR: 3 Prime Untranslated Region; 5’UTR: 5 Prime Untranslated Region; PST: protein S total; PSF: protein S free.

### Characterization and Prioritization of Candidate Loci

#### Fine-Mapping and Colocalization

To prioritize causal genes among those residing at associated locus, we performed fine-mapping and colocalization. Detailed methods for fine-mapping and colocalization can be found in the **Supplementary Methods**.

#### In vitro Functional Validation

Functional validation of prioritized candidates was performed by *in vitro* silencing of candidate genes in a liver-derived hepatoblastoma cell (HepG2) expression system. Briefly, HepG2 cells were reverse transfected with small interfering RNA (siRNA) against candidate genes. Cells were counted, and target proteins and genes were characterized by immunoblot of cell supernatants and RT-qPCR, respectively. Normalized, log transformed data were compared using one-way ANOVA with Dunnett’s multiple comparisons test. Details on cell culture, transfection, RNA extraction, RT-qPCR, and immunoblotting methods can be found in **Supplementary Methods**.

### Mendelian Randomization

Two-sample summary statistics-based MR was used to assess the association of genetically determined levels of antithrombin and PC with the risk of thrombotic outcomes, VTE^41,42^, peripheral artery disease (PAD) (31,307 cases and 211,753 controls)^43^, coronary artery disease (CAD) (60,801 cases and 123,504 controls)^44^, and ischemic stroke (IS) (60,341 cases and 454,450 controls)^45^. Given the small proportion of variance explained by the identified PS variants we did not investigate PS (PS_free_ and PS_total_) in MR analyses because of insufficient number of genetic instruments.

All analyses were performed using the ‘TwoSampleMR’ v0.4.26 and ‘MRPRESSO’ v1.0 R packages. Further details are reported in the **Supplementary Methods**.

## RESULTS

Antithrombin activity (% or IU/mL*100, n = 26,999) or antigen (IU/mL*100; n = 932) was measured in 9 cohorts, PC activity (% or IU/mL*100; n = 6,734) or antigen (μg/mL; n = 12,551) was measured in 8 cohorts, PS Total (PS_total_) activity (% or IU/mL*100 or IU/mL; n = 5,045) or antigen (IU/mL or μg/dL; n = 1,363) was measured in 7 cohorts, and PS Free (PS_free_) activity (μg/dL or %; n = 1,998) or antigen (IU/mL*100; n = 2,115) was measured in 6 cohorts. See **Supplementary Table S4**.

### Antithrombin

#### GWAS

The antithrombin meta-analysis include 25,243 EA and 2,688 AA participants. After quality control and filtering, 80,168,840 variants remained in the meta-analysis. All λ_GC_ for individual GWAS were 1.04 or below for all chromosomes. Additional details about quality control are provided in **Supplementary Table S5 and Supplementary Material**. Manhattan plots for the overall cross-ancestry meta-analyses are shown in **Figure 2**. A quantile-to-quantile plot (QQ plot) of p-values for these variants is presented in **Supplementary Figure S1** and Manhattan plots for the EA and AA population specific analyses are available at **Supplementary Figure S2**.

**Figure 2.**
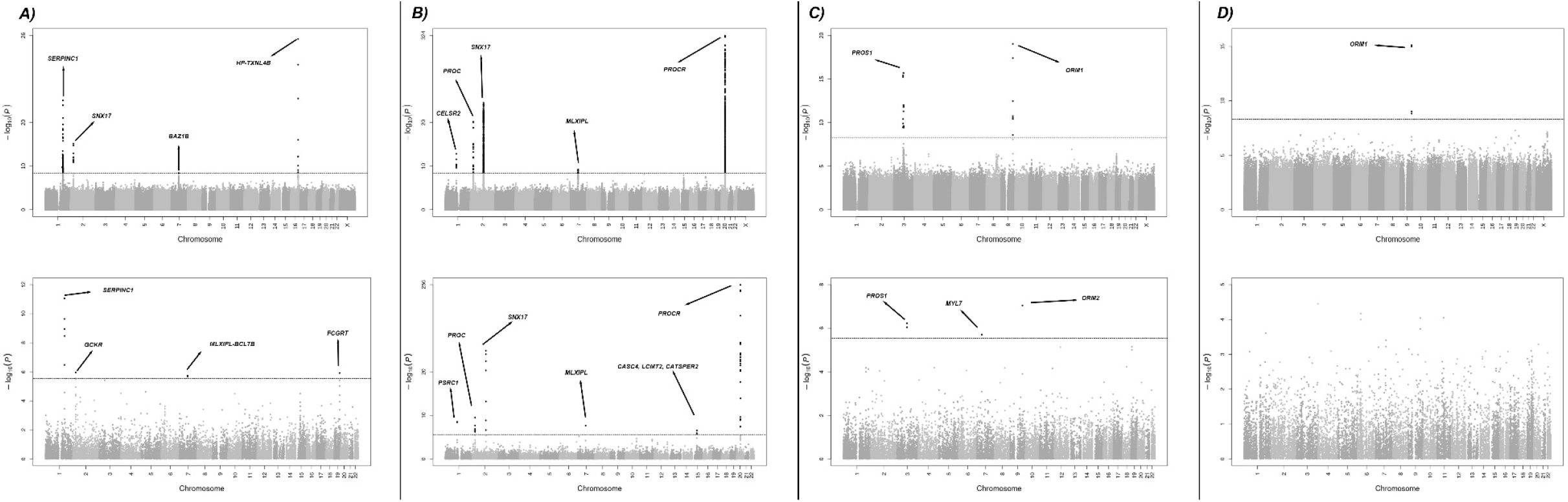
Manhattan plots for discovery meta-analyses of GWAS (up) and TWAS (down) results (A) Antithrombin (B) Protein C (C) Protein S Free (D) Protein S Total. Dots represent all allelic variants (GWAS) or genes (TWAS) sorted by chromosome and position throughout the X-axis. Y-axis report inverse log transformed p-value for the associations.

In total, 402 variants in 4 loci, exceeded the established genome-wide significance level in the cross-ancestry analysis, 394 (2 loci) in the EA-specific analysis and 57 (1 locus) in the AA-specific analysis. Forest plots for significant variants can be found in **Supplementary Figure S3**. Loci at *SNX17-GCKR-NRBP1* (2p23.3), *MLXIPL-BAZ1B-BCL7B* (7q11.23) and *HP-TXNL4B* (16q22.2) were new associations. The association at *HP-TXNL4B* (16q22.2) was only found in the AA population. Lead variants in the cross-ancestry meta-analysis in each region are listed in **Table 1** along with the meta-analysis p-value, ancestry specific p-value, effect allele frequency (EAF), beta estimates, and closest gene.

Conditional analyses using the population specific meta-analyses (**Supplementary Table S6**), identified no additional independent variants on *SNX17-GCKR-NRBP1* and *HP*-*TXNL4B* surrounding regions. On chromosome 1 locus (*SERPINC1*), we found 1 variant (rs182221508, MAF = 0.0017) intronic to *RABGAP1L* gene (600 kb upstream the lead variant), that was independent from the lead missense variant rs2227624 on *SERPINC1* gene.

**Supplementary Table S7** shows the lead variants with the strongest associations in the EA and AA meta-analyses. There was 1 significant locus in the AA population specific analysis at chromosomal position 16q22.2 (*HP-TXNL4B*), which also appeared in cross-ancestry analysis. In the EA-specific population analysis, the results reflected cross-ancestry findings at 1q25.1 (*SERPINC1*) and 2p23.3 (*SNX17-GCKR-NRBP1*), with a different lead variant on chromosome 2: rs4665972, located in an intronic region of *SNX17*, was the lead variant in the cross-ancestry analysis, while rs11127048,150 kb upstream rs4665972 and located in an intergenic region between *SNX17* and *GCKR* genes was the lead variant in the EA-specific analysis. We did not find significant signals at 7q11.23 in the EA-specific analysis. The proportion of variance explained by the independent lead variants was 1.4% in EA and 4.3% in AA, of the total antithrombin variance.

All lead variants from GWAS were replicated in the deCODE summary results derived from SOMAscan measures of these anticoagulants, except for the lead variant of the chromosome 16 locus, that was specific for the AA population and was not present in the DeCODE data (**Table 1 and Supplementary Table S8**).

#### TWAS

TWAS analyses identified associated genes in 4 different loci (**Figure 2A**). Associations on chromosomes 1 (*SERPINC1*), 2 (*GKCR*) and 7 (*MLXIPL*), identified by the strongest associated gene in the TWAS, matched associated loci found in the GWAS. Additionally, the *FCGRT* gene represented a new association on chromosome 19. The smallest GWAS p-value for this region approached significance and was for a rare intronic variant (rs111981233) in *FCGRT* gene (**Figure 2 and Supplementary Table S9**) that was replicated in the DeCODE cohort (**Table 1 and Supplementary Table S8**).

#### Fine Mapping

EA-specific fine-mapping results prioritized the *SERPINC1* gene on chromosome 1 and the *NRBP1* gene on chromosome 2. Given that FOCUS only prioritizes GWAS hits at TWAS risk loci, loci on chromosomes 16 (only GWAS) or 19 (only TWAS) could not be further explored for gene prioritization. In addition, after correcting for LD and pleiotropic effects, none of genes in chromosome 7 locus was included in the credible set, suggesting a regulation mechanism that does not involve gene expression **(Supplementary Table S10)**.

#### Colocalization

We obtained 2 significant colocalizations in lead variants located in the new antithrombin loci (CPC > 0.8) and gene expression of nearby genes. On chromosome 2, *GTF3C2*-*AS2 (at SNX17*-*GCKR-NRBP1 locus)* gene expression in artery tibial tissue colocalized with antithrombin plasma levels and on chromosome 16 locus, *HP* gene expression in liver and whole blood also colocalized with antithrombin plasma regulation (**Supplementary Table S11**).

#### Functional Validation

We selected 1-3 genes per locus for functional analysis (5 genes total): *SNX17, GCKR, NRBP1* (Chr 2), and *HP* (Chr 16). *LCMT2* (Chr 15) was also included for its association in the multi-phenotype analysis at *MAP1A* locus. We transfected HepG2 cells with siRNA against each candidate gene and confirmed that target genes were knocked down more than 60% using RT-qPCR (data not shown). We then characterized effects of the gene knockdowns on cell count. Finally, we quantified antithrombin expression by immunoblot of cell supernatants and *SERPINC1* expression by RT-qPCR. As expected, control experiments showed that treatment of HepG2 cells with lipofectamine (alone) or siRNA against *PROC* did not significantly alter antithrombin (protein) or *SERPINC1* (gene) expression, whereas silencing *SERPINC1* significantly suppressed antithrombin and *SERPINC1* expression (**Figure 4A-B**).

Quantification of immunoblots revealed that silencing *GCKR* enhanced, whereas silencing *SNX17* and *HP* suppressed, antithrombin protein production (**Figure 4A**). The *GCKR*-dependent increase in antithrombin was associated with a significant increase in *SERPINC1* expression, suggesting *GCKR* negatively regulates antithrombin gene expression (**Figure 4B**). The *SNX17*-dependent loss of antithrombin was associated with a significant decrease in *SERPINC1* expression, suggesting *SNX17* positively regulates antithrombin gene expression (**Figure 4B)**. Interestingly, *HP*-dependent loss of antithrombin was not accompanied by a significant decrease in *SERPINC1* expression (**Figure 4B**) suggesting that *HP* modifies antithrombin production in a post-transcriptional manner.

#### MR analysis

We used 4 genetic instruments (**Supplementary Table S12**) to investigate the association between antithrombin levels and VTE and PAD, and 3 to investigate its association with CAD and IS. We detected a significant deleterious effect of genetically determined low antithrombin levels and risk of VTE (IVW OR 0.84 [0.72-0.97], P-value: 0.015; **Figure 3A)**. Sensitivity analyses showed consistent effect in size and direction with MR Egger, MR weighted median, and MR weighted mode (**Supplementary Table S13** and **Supplementary Figure S4**). Leave-one-out sensitivity analyses showed homogeneity of effects among the instruments. No significant results were found for the association of genetically determined antithrombin levels with IS, CAD or PAD (**Figure 3A** and **Supplementary Figure S4**).

**Figure 3.**
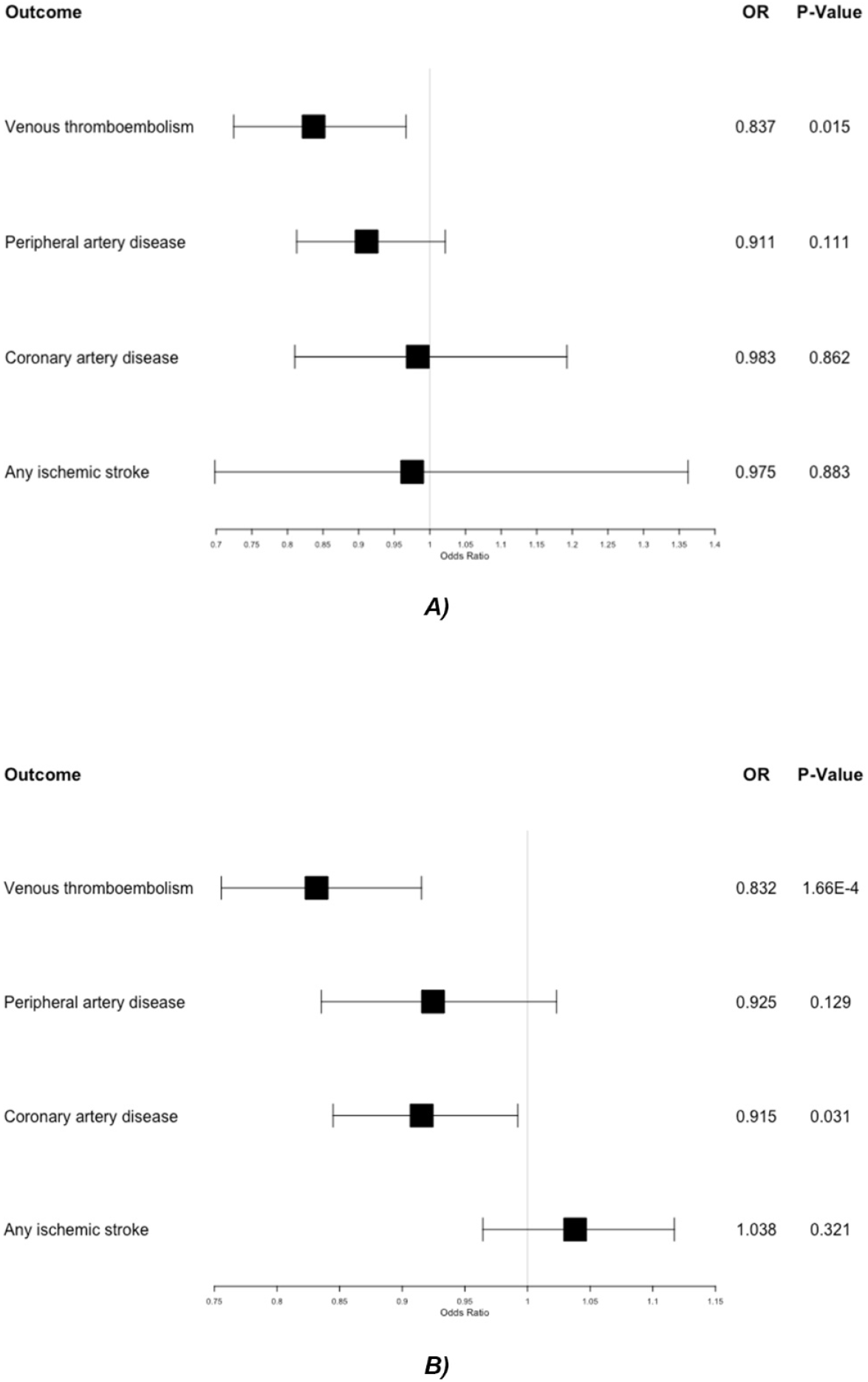
**Forest plot showing** inverse variance weighted mendelian randomization results for multiple outcomes using antithrombin (A) and protein C (B) as exposure. Squares indicate OR (95% CI).

**Figure 4.**
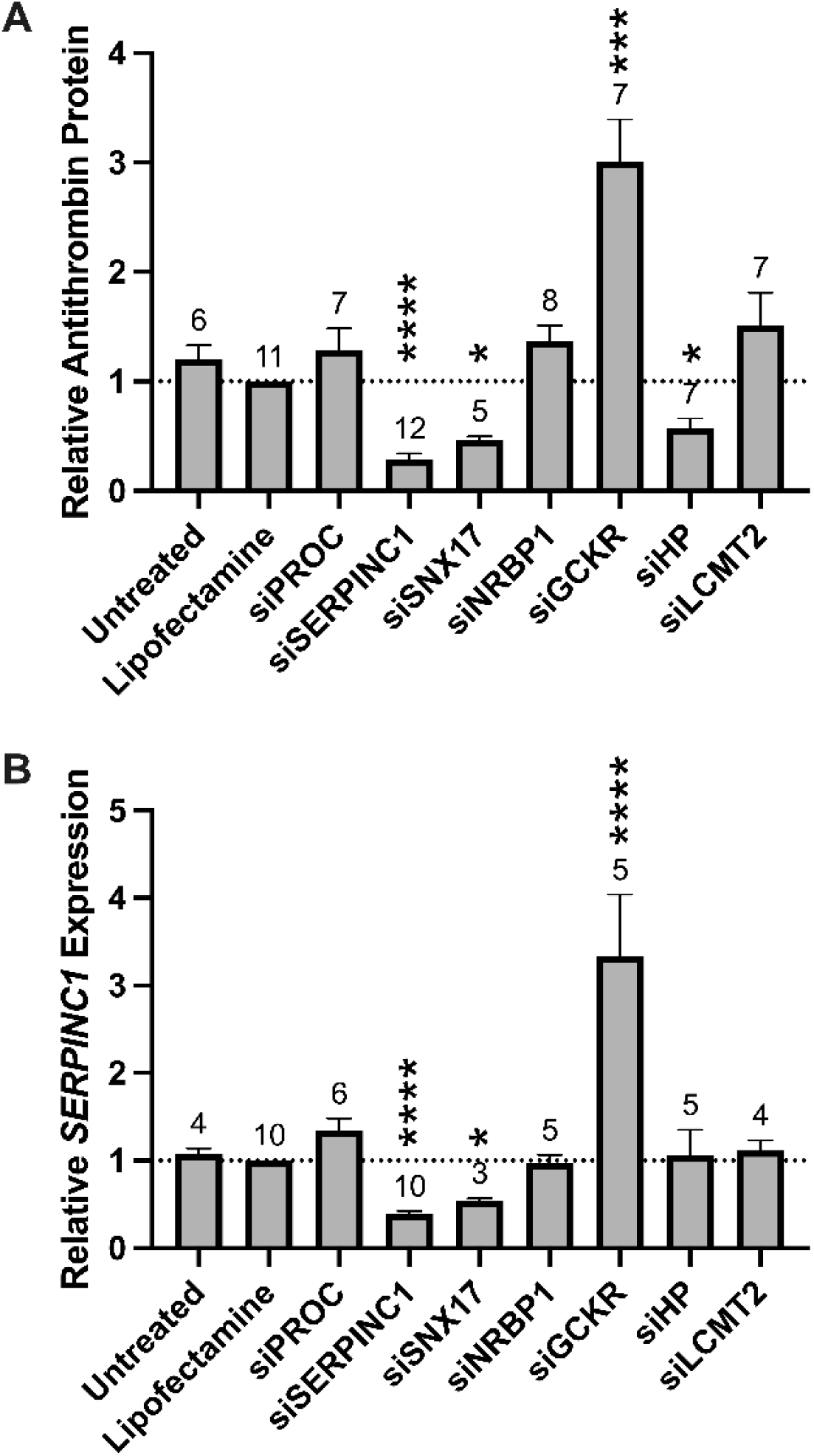
Knockdown of *GCKR, SNX17*, and *HP* alter antithrombin production in HepG2 cells. A) Antithrombin secreted into the culture supernatant was detected by immunoblot (upper) and quantified by densitometry (lower). B) *SERPINC1* expression was measured by RT-qPCR. Bars and error bars indicate mean and standard error of the mean; Numbers indicate biological replicates; *P-value < 0.05; ***P-value < 0.0005; ****P-value < 0.0001

### Protein C

#### GWAS

The PC meta-analysis included 16,597 EA and 2,688 AA participants. After quality control, 72,929,079 variants were included. All λ_GC_ for individual GWAS were 1.04 or below for autosomal chromosomes (1.18 for X chromosome). Additional details about quality control are provided in **Supplementary Table S5 and Supplementary Material**.

Manhattan and QQ-plots showing the cross-ancestry meta-analysis results are presented in **Figure 2 and Supplementary Figure S1**, respectively. Briefly, 2,198 variants exceeded the genome-wide significance level in the main analysis, identifying 5 regions associated with PC levels. All loci, located near *CELSR2-PRSC1* (1p13.3), *PROC* (2q14.3*), SNX17-GCKR-NRBP1* (2p23.3), *MLXIPL-TBL2* (7q11.23), and *PROCR* (20q11.22) genes, have been previously reported to be associated with PC. Coefficients, p-values, ancestry stratified EAF and p-values, and closest genes are listed in **Table 1**. Forest plots of significant signals found in the GWAS analysis can be found at **Supplementary Figure S5**.

In the conditional analysis at 1p13.3 (*CELSR2*-*PRSC1)*, 2p23.3 (*SNX17-GCKR-NRBP1)*, and 7q11.23 (*MLXIPL*-*TBL2)* loci in the EA population, no additional independent variants were identified (**Supplementary Table S6**). Within the *PROC* locus on chromosome 2, an additional independent variant (rs74392719, MAF = 0.01, 300 bases upstream of the lead variant) was identified in the EA population, located within the *PROC* gene. Finally, an additional independent variant (rs6060300, MAF = 0.2, 13 kb upstream of the lead variant) was found in the EA population, intronic to *PROCR*.

No significant heterogeneity was found in the direction or magnitude of beta coefficients for any of the lead variants associated with PC, within or between ancestries. AA and EA population-specific results are shown in **Supplementary Table S7** and **Supplementary Figure S2**. The AA population analysis had findings at 2q14.3 (*PROC*) and 20q11.22 (*PROCR*); the EA population analysis recapitulated all the candidate loci found in cross-ancestry analysis. The proportion of variance explained by the identified independent variants was 12.7 % in EA and 7.4% in AA. The lead variants at the *PROCR* locus (rs11907011 and rs867186) alone explain 9.5% and 9% of the total variance in the EA and AA meta-analyses, respectively. All lead variants from GWAS were replicated in the DeCODE data (**Table 1** and **Supplementary Table S8**).

#### TWAS

For PC levels, TWAS (**Figure 2B**) identified associated genes at 6 loci, matching all loci found in the cross-ancestry and EA GWAS, of which, the most significant based on TWAS z-score values were *PSRC1* (chromosome 1, *CELSR2*-*PRSC1 locus*), *GCKR* (chromosome 2, *SNX17-GCKR-NRBP1* locus), *PROC* (chromosome 2), *MLXIPL* (chromosome 7, *MLXIPL-TBL2* locus) and *PROCR* (chromosome 20). Additionally, 3 new associations with PC were found in 1 locus on chromosome 15 for *GOLM2, LCMT2* and *CATSPER2* genes (**Supplementary Table S9**) (**Table 1 and Supplementary Table S9**).

#### Fine Mapping

Fine-mapping results for PC prioritized the *PSRC1* gene on chromosome 1, *NRBP1* and *PROC* on chromosome 2 (*SNX17-GCKR-NRBP1 and CELSR2-PRSC1 locus*, respectively), *MLXIPL* and *TBL2* on chromosome 7 (*MLXIPL-TBL2* locus), and *PROCR* on chromosome 20. (**Supplementary Table S10**).

#### MR analysis

For PC, 4 variants were initially selected as genetic instruments. After examination of pleiotropic effects, the variant at the *PROCR* gene (rs1799809) was excluded to avoid violations of MR assumptions. Moreover, additional evidence indicates that this variant is strongly associated with several hemostasis and thrombosis phenotypes and has opposite effect directions for venous and arterial thrombosis reflecting distinct pleiotropic biological mechanisms^18,46,47^. Details of selected genetic instruments can be found in **Supplementary Table S12**. There was a significant deleterious effect of genetically determined lower PC levels on VTE and CAD risk (VTE IVW OR:0.83 (0.76-0.92), P-value: < 0.001; CAD IVW OR: 0.92 (0.84-0.99), P-value: 0.031; **Figure 3B**). Sensitivity analyses showed consistent significant associations (**Supplementary Table S13** and **Supplementary Figure S4**). No significant associations were found between genetically determined PC with PAD or IS (**Figure 3B and Supplementary Figure S4**).

### Protein S

#### GWAS

The PS meta-analysis included 4,113 EA individuals in PS_free_ analyses and 6,408 EA individuals in PS_total_ analyses. A total of 19,791,246 variants were investigated in the analysis of PS_free_ and 25,365,467 in the analysis of PS_total_. All λ_GC_ for individual GWAS were 1.04 or below for autosomal chromosomes (1.19 for X chromosome). Additional details about quality control are provided in **Supplementary Table S5 and Supplementary Material**. Manhattan and QQ-plots describing the main results are shown in **Figures 2C/D** and **Supplementary Figure S1** for PS_free_ and PS_total_, and main associated variants are listed in **Table 1**. Forest plots of significant signals for PS_free_ and PS_total_ can be found in **Supplementary Figure S8**.

We identified 1 novel genome-wide significant locus associated with PS_free_ and PS_total_ near *ORM1* and *ORM2* genes (9q32) and a known association located near *PROS1* gene (3q11.1) for PS_free._ The lead variant at *PROS1* locus (rs121918472, EA P-value = 2.04 × 10^−16^, PS_free_ EAF (G) = 0.0108) was a missense variant located in the protein S coding gene *PROS1*. In our analysis, this variant was associated with PS_free_ level, but genome-wide significance was not observed in PS_total_ (PS_total_ P-value = 2 × 10^−4^) although there was a consistent direction of effect.

Nominally significant heterogeneity p-values were detected in the *ORM1/ORM2* locus lead variant (PS_total_ Heterogeneity P-value = 0.03), indicating minor differences between the 2 measurement methods. No additional independent variants were found with conditional analyses (**Supplementary Table S6**). The variance explained by the identified variants in PS_free_ is 6% of the total variance of PS_free_ while the variance explained by the unique identified variant in PS_total_ is 1% of the phenotypic variance for PS_total_.

Variants at both loci replicated in the DeCODE data (**Table 1 and Supplementary Table S8**).

#### TWAS

PS_free_ TWAS results recapitulated the 2 significant GWAS associations at chromosomes 3 (*PROS1*) and 9 (*ORM2*) and additionally revealed a new association at *MYL7* gene on chromosome 7 **(Figure 2 and Supplementary Table S9)**.

#### Fine Mapping

Fine-mapping results did not prioritize any genes for PS_free_ or PS_total_.

#### Colocalization

There was a significant colocalization for both PS phenotypes and *ORM2* gene expression in liver (**Supplementary Table S11**).

#### MR analysis

Given the small proportion of variance explained by the limited number of genetic instruments (< 3), we did not investigate PS (PS_free_ and PS_total_) in MR analyses.

### Antithrombin, Protein C and S Multi-phenotype Analysis

Multi-phenotype analyses between antithrombin, PC, PS_free_ and PS_total_ revealed 1 additional novel GWAS association close to the *MAP1A* gene^48^, on chromosome 15 (**Table 1**), found in the PC TWAS (*GOLM2-LCMT2-CATSPER2* locus in PC). The lead variant is a missense variant on the *MAP1A* gene (rs55707100, P-value = 1.64 × 10^−13^, EAF EA [T] = 0.03, EAF AA [T] = 0.0042) that was nominally associated in the GWAS for antithrombin and PC individually (antithrombin P-value = 1.04 × 10^−6^, PC P-value = 4.76 × 10^−8^) and was not significantly associated to either of the PS phenotypes (PS_total_ P-value = 0.2717, PS_free_ P-value = 0.9937). The colocalization results were significant (CPC > 0.8) between antithrombin and PC, suggesting the existence of a common variant as regulator of both phenotypes.

## DISCUSSION

In this study, we performed GWAS for 4 natural anticoagulant hemostasis phenotypes (antithrombin, PC, PS_total_, PS_free_) using larger sample sizes and better imputation panels than previously reported and detected 4 novel associations: 3 loci for antithrombin (*SNX17-GCKR-NRBP1, MLXIPL*-*BAZ1B-BCL7B*, and *HP-TXNL4B*) and 1 locus for PS (*ORM1-ORM2*). For 3 genes within the newly associated loci with antithrombin (*SNX17, GCKR*, and *HP*), *in vitro* gene silencing in liver cell experiments provided functional evidence. Using TWAS methods, we detected 3 more novel associations that did not reach significance in individual GWAS: *FCGRT* for antithrombin; *GOLM2* for PC; and *MYL7* for PS. Using MR, we also identified a causal relationship of antithrombin and PC levels with VTE, and of PC levels with CAD. This investigation elucidated genetic regulation of the anticoagulant pathway and provides new information that could identify therapeutic targets in VTE prevention or treatment.

Additionally, we replicated 7 known loci. These loci are *SERPINC1* for antithrombin^6^; *CELSR2-PRSC1*^16^, *PROC*^14,16,17^, *SNX17-GCKR-NRBP1*^14,16^, *MLXIPL*-*TBL2*^14,16^ and *PROCR*^14,16-18^ for PC; and *PROS1* for PS^8^. Two of the PC loci, *SNX17-GCKR-NRBP1* and *MLXIPL*-*BAZ1B-BCL7B*-*TBL2*, also had novel associations with antithrombin, demonstrating some genetic overlap between different anticoagulant proteins. This was also reflected in the multi-phenotype analysis results where *MAP1A* was identified.

### Characterization of Novel Loci

#### Antithrombin-associated Loci

More than 45 rare variants within the *SERPINC1* gene have already been described using non-GWAS approaches^49^. Our lead variant, rs2227624, is a known missense variant causing a Val to Glu amino-acid substitution that leads to antithrombin deficiency^50-52^ and increases risk of VTE^53^. We identified a second independent variant at this locus at *RAPBGAP1L* and believe this rare variant is likely in LD with a *SERPINC1* variant but we were unable to demonstrate this due to the limitation of the CGTA conditional analyses.

On chromosome 2, lead variants in locus *SNX17*-*GCKR-NRBP1* differed by ancestry. In the cross-ancestry analysis, rs4665972 was in an intronic region of *SNX17* whereas, in the EA-specific analysis, the lead variant (rs11127048) was located in an intergenic region between the *SNX17* and *GCKR* genes. Neither rs4665972 nor rs11127048 were significant in AA population suggesting that these variants are tagging an association within a large LD block in EA population. Consistent with this observation, conditional results indicate that the lead variant (rs4665972) is the only independent variant on this locus. Given limited power in the AA-specific analysis, we could not refine the region with AA data (**Supplementary Figure S10**).

Functional validation in liver-derived cells suggest that *SNX17* positively, and *GCKR* negatively, alters plasma antithrombin levels via effects on *SERPINC1* expression. In contrast, *HP* appears to suppress antithrombin levels through an as-yet unidentified post-transcriptional mechanism.

*SNX17* is a regulator of low density lipoprotein (LDL) receptors^54^ and has not been previously associated to antithrombin levels but has been associated with CAD^55,56^. *GCKR* is a highly pleiotropic gene, that has been found significantly associated to PC^15,16^, Factor VII (FVII)^57^, Factor XI (FXI)^58^ and C-reactive protein (CRP)^59,60^ in previous GWAS meta-analyses. In previous candidate gene studies^61-63^, variant rs1260326 in *GCKR* was found to be related to multiple cardiometabolic traits, including total and LDL cholesterol, fasting plasma glucose, liver fat content and metabolic syndrome, suggesting that *GCKR* might act as a broad regulator of hepatocyte function.

On chromosome 7, the lead variant (rs13244268) was located in an intronic region of *BAZ1B* gene and was only significant in the EA population. This gene has been previously associated with PC^15,16^ and in our PC meta-analysis, but not with antithrombin. rs13244268 was also found significant in bivariate and univariate GWAS of CRP and high-density lipoprotein^64^. TWAS results confirmed an association between *BCL7B* and *MLXIPL* genes and antithrombin levels. Given the differences in LD blocks observed for this region in different populations, we sought to confirm the most plausible candidate genes in this locus with *in vitro* silencing studies in liver cells. Within the 3 closest genes in the region (*BAZ1B, MLXIPL* and *BCL7B*), *BAZ1B* and *BCL7B* are involved in chromatin remodeling and DNA repair and were not prioritized for functional validation. *MLXIPL* is a highly expressed transcription factor in liver that activates triglyceride synthesis in response to carbohydrates. However, this gene is substantially downregulated in HepG2 cells, and we therefore excluded it from further validation. As such, we did not have a good candidate for the functional work.

The lead variant on *HP*-*TXNL4B* locus (rs5471) was in an intronic region of the *TXNL4B* gene and 5’ UTR of the *HP* gene and was only significant in the AA population. Colocalization results performed using cross-ancestry data suggested the existence of a common regulatory variant between *HP* gene expression and antithrombin levels in liver and whole blood, and suggested that higher expression of *HP* in liver and blood were associated with higher levels of antithrombin in plasma. In the same direction, functional validation in HepG2 cells suggested a significant reduction of antithrombin levels upon *HP* silencing. *HP* codes for haptoglobin (Hp), which serves as a binding protein of hemoglobin, and affects the release of hemoglobin from red blood cells^65^. Its phenotype Hp2-2 was identified as a potential regulator of inflammation and reverse cholesterol transportation and has been suggested to have higher prevalence in VTE patients^66-68^. Binding of Hp to hemoglobin could prevent the generation of oxidized LDL^69^ from the activation pathway of free hemoglobin. Oxidized LDL could increase prothrombinase activity in vivo^70^, and affect the level of factors VII, IX and XI^71^. Overall, previous evidence suggests a potential role of Hp in the inflammation-induced thrombosis, and our results suggest *HP* is a potentially direct regulator of antithrombin production.

Finally, TWAS results suggested a novel locus associated to antithrombin levels on the *FCGRT* gene. Colocalization results suggested the existence of a common regulatory variant between antithrombin levels and the expression of *FCGRT, RPS11* and *RCN3* in the aorta, tibial artery, and whole blood. In GWAS analysis, rs111981233 (intronic to *FCGRT*) nearly reached genome-wide significance levels. *FCGRT* encodes a receptor that binds immunoglobulin G and transfers immunoglobulin G antibodies from mother to fetus across the placenta^72^ and previous studies demonstrate that *FCGRT* is also expressed in the liver^73,74^. Additional work is needed to further elucidate the role of this gene in antithrombin regulation.

#### Protein C-associated Loci

We found 5 loci associated with PC in the present GWAS meta-analysis, all of which had been previously described. In addition, 3 genes (*GOLM2, LCMT2, and CATSPER2)* were associated in a novel locus on chromosome 15 in the TWAS analysis. *GOLM2* encodes for a transmembrane protein predicted to colocalize in the Golgi apparatus with no known function; *LCMT2* acts as a checkpoint regulator in the cell cycle; and *CATSPER2* codes for a protein crucial for correct function of sperm cells. Interestingly, a variant near this locus was significant when in a multi-phenotype GWAS analysis, and colocalization results suggested the existence of a common variant between antithrombin and PC. The lead variant in the multi-phenotype analysis is a missense variant located on *MAP1A* gene that has been associated with lipid metabolism^75,76^ and platelet count^77^. *MAP1A* had low expression in hepatocytes and could not be included for further functional analyses^78^.

#### Protein S-associated Loci

For PS, genome-wide associations found at the *ORM1-ORM2* locus represented novel findings for both PS phenotypes, and colocalization analysis suggests the existence of a common regulatory variant between PS_free_ and PS_total_ levels and *ORM2* expression in liver. *ORM1* is responsible for encoding acute phase plasma protein orosomucoid (ORM, also known as α1-acid-glycoprotein, AGP), which is increased with acute inflammation^79^. Previous genetic results suggested that *ORM1* was associated with thrombin generation potential^80^ and the discovery was further confirmed with *in vitro* experiments. *ORM1* has also been associated with cell-free DNA levels in plasma, a surrogate marker of neutrophil extracellular traps that contribute to immunothrombosis^81^. Moreover, AGPs encoded by the *ORM1* and *ORM2* genes strongly bind to the vitamin K antagonist warfarin that reaches circulation, suggesting that these genes could be relevant in regulating the response to oral anticoagulation^82^. Supporting this hypothesis, *ORM1, ORM2* and *PROC* were nominally associated with warfarin dose requirement in a study of candidate gene analysis with 201 patients^83^. This is interesting, since it is widely known that one of the challenges in oral anticoagulation is the wide variation in response among patients^84^. Confirming novel genomic regulators of anticoagulant response could help explain the mechanisms of action of these drugs and move towards a personalized treatment based on genomic background. Our results suggest an involvement of *ORM* genes in PS regulation.

*MYL7*, associated with PS levels in TWAS analyses, is the coding gene for myosin light chain 7, and is related to calcium ion binding activity^85,86^. Variants in this gene have been associated with fasting glucose levels and type II diabetes^87,88^ probably for their proximity to the *glucokinase (GCK)* gene, which lies 1.9 kb upstream of *MYL7*, and is essential for producing glucose-6-phosphate. Variants at *GCK* have been associated with multiple types of diabetes and hemoglobin A1c measurement.

##### Implication with for disease outcomes

The present MR results confirm a causal relationship between genetically determined plasma levels of antithrombin and PC with VTE events, and for PC with CAD outcomes. Specifically, we observed a 19% VTE risk increase per 1 SD decrease in antithrombin plasma levels, a 20% VTE risk increase per 1 SD decrease of PC plasma levels, and a 9% CAD risk increase per 1 SD decrease in PC plasma levels. Our findings of a causal relationship of antithrombin and PC with VTE agree with previous epidemiological studies that report an increased VTE risk in individuals with deficiencies of these anticoagulants^5,89,90^. The causal relationship between PC and CAD was also reported in previous epidemiological and MR studies.^91,92^ Overall, these results support previous data suggesting that AT and PC are relevant proteins that regulate the risk of VTE, confirmed the causal association between PC levels and CAD, and corroborated that intervention in the anticoagulant system could be considered for VTE or CAD prevention^93,94^.

##### Strengths and limitations

A major strength of this study is in the modestly large sample size, including around 30,000 individuals, compared with more limited studies in the previous discovery efforts. Additionally, the TOPMed imputation panel, provides better imputation quality for low-frequency variants compared with previous panels, which increases our power to detect rare variation. However, the present study was not designed to provide a detailed evaluation of rare variation within coding genes, and some rare variants within these genes were excluded from the analyses if they were presented in less than 2 cohorts. Larger studies combined with whole genome sequencing data will help identify novel rare (familial) associations for these phenotypes and may provide better instruments that will improve the power for MR studies.

Inclusion of AA ancestry individuals has allowed the identification of novel associated loci for antithrombin in this population. There is a recent debate ^95-97^ on transferability of results from GWAS studies to non-European populations, given the overwhelming majority of GWAS results in EA populations for most phenotypes. Although our sample was predominantly of EA, we were able to observe differences in LD blocks between EA and AA ancestry groups, which allowed us to detect novel associations in variants with lower frequency in the EA population, and to refine loci where the linkage blocks differed between ancestries. However, some of the follow-up methods (TWAS, approximate conditional analyses) depend on population reference panels and were limited to the EA population.

Finally, to reduce the risk of false positives, we used a stringent significance threshold (5 × 10^−9^), sought replication of the main findings in an external proteomics cohort, and provided additional post-GWAS evidence for our novel findings. We included functional validation using *in vitro* silencing to provide evidence for causality of candidate genes and help understand the biological mechanism. We believe this strengthens the credibility of our results. However, liver cell-derived expression system is only able to assess effects of candidate genes on synthetic mechanisms (e.g., transcription, translation), and is not able to assess potential effects on protein stability and/or clearance. Thus, genes that did not demonstrate an effect, as well as genes that were not selected for testing in this system, could regulate circulating anticoagulant protein expression via synthesis-independent mechanisms.

##### Summary

Using cross-ancestry GWAS and TWAS methods, we report 7 novel associations for antithrombin, PC, and PS plasma levels: 4 novel loci regulating antithrombin plasma levels, 2 novel loci regulating PS plasma levels, and 1 novel locus regulating PC plasma levels. Post-GWAS analyses and functional work suggest both *SNX17* and *GCKR* are regulators of antithrombin on the chromosome 2 locus and validate an AA-specific *HP* gene locus. MR analyses provided evidence implicating low antithrombin levels in VTE risk and low PC levels in VTE and CAD risk. Overall, our findings identified novel pathways regulating the main anticoagulant proteins in hemostasis and strengthen their implication on disease outcomes.

## Data Availability

All data produced in the present study are available upon reasonable request to the authors and will be available through dbGAPs after publication.

## ABBREVIATIONS

TOPMed: Trans-Omic for Precision Medicine
PC: protein C
PS: protein S
VTE: venous thromboembolism
CAD: coronary artery disease
PAD: peripheral artery disease
IS: ischemic stroke
GWAS: genome-wide association study
TWAS: transcription-wide association study
EA: European ancestry
AA: African ancestry
eQTL: expression quantitative trait locus

## ACKNOWLEDGMENTS

We would also like to thank MEGASTROKE, DeCODE genetics, INVENT Consortium, CARDIoGRAMplusC4D and Million Veterans Program for making their data publicly available. The MEGASTROKE project received funding from sources specified at http://www.megastroke.org/acknowledgments.html. Appendix 1 contains a list with all authors that contributed to MEGASTROKE project (https://www.megastroke.org/authors.html). Data on coronary artery disease has been contributed by CARDIoGRAMplusC4D investigators and have been downloaded from www.CARDIOGRAMPLUSC4D.ORG. Data from DeCODE genetics was accessed through DeCode genetics web page at decode.com. VTE and PAD data from de Million Veteran Program were accessed through dbGAPs (phs001672 v7.p1).

## SOURCES OF FUNDING

This study is supported in part by the National Heart, Lung, and Blood Institute grant HL134894 and HL139553; infrastructure for the CHARGE Consortium is supported in part by HL105756. The views expressed in this manuscript are those of the authors and do not necessarily represent the views of the National Heart, Lung, and Blood Institute; the National Institutes of Health; or the US Department of Health and Human Services. G. Temprano-Sagrera is supported by the *Pla Estratègic de Recerca i Innovació en Salut (PERIS)* grant from the Catalan Department of Health for junior research personnel (SLT017/20/000100). P de Vries is supported by American Heart Association grant 17POST33350042. M. Sabater-Lleal is supported by a *Miguel Servet* contract from the ISCIII Spanish Health Institute (CP17/00142) and co-financed by the European Social Fund, and acknowledges funding from the CERCA Programme/Generalitat de Catalunya. Sources of funding for the specific cohorts can be found in the online-only Data Supplement.

## DISCLOSURES

None

## Notes

### Competing Interest Statement

The authors have declared no competing interest.

### Author Declarations

The ARIC study was approved by the UNC Office of Human Research Ethics/Institutional Review Board (OHRE/IRB); University of Mississippi Medical Center IRB, Wake Forest University Health Sciences IRB, University of Minnesota IRB, and John Hopkins University IRB. CHRIS study was approved by the Ethical Committee of the Healthcare System of the Autonomous Province of Bolzano CHS was approved by the Wake Forest University Health Sciences IRB, University of California, Davis IRB, John Hopkins University IRB, and University of Pittsburgh IRB, and University of Washington IRB GAIT was approved by the Institutional Review Board of the Hospital de la Santa Creu i Sant Pau, Barcelona, Spain The HVH study is part of data repository which is under the oversight of the University of Washington Human Subjects Division. LURIC was approved by the Ethics Committee at the Arztekammer Rheinland-Pfalz MARTHA was approved by the Health Department of the General Directorate for the French Ministry of Research and Innovation (Projects DC: 2008-880 and 09.576) RETROVE was approved by Institutional Review Board of the Hospital de la Santa Creu i Sant Pau, Barcelona, Spain The Genes and Blood Clotting study GABC was approved by the University of Michigan IRB. #HUM00043243. For the Trinity Student Study (TSS), ethical approval was obtained from the Dublin Federated Hospitals Research Ethics Committee, which is affiliated with the Trinity College, and reviewed by the Office of Human Subjects Research at the National Institutes of Health.

## REFERENCES

1. Broekmans AW, Veltkamp JJ, Bertina RM. Congenital protein C deficiency and venous thromboembolism. A study of three Dutch families. N Engl J Med. 1983;309:340–344. doi: 10.1056/nejm198308113090604

2. Griffin JH, Evatt B, Zimmerman TS, Kleiss AJ, Wideman C. Deficiency of protein C in congenital thrombotic disease. J Clin Invest. 1981;68:1370–1373. doi: 10.1172/jci110385

3. Schwarz HP, Fischer M, Hopmeier P, Batard MA, Griffin JH. Plasma protein S deficiency in familial thrombotic disease. Blood. 1984;64:1297–1300.

4. Egeberg O. INHERITED ANTITHROMBIN DEFICIENCY CAUSING THROMBOPHILIA. Thromb Diath Haemorrh. 1965;13:516–530.

5. Folsom AR, Aleksic N, Wang L, Cushman M, Wu KK, White RH. Protein C, antithrombin, and venous thromboembolism incidence: a prospective population-based study. Arterioscler Thromb Vasc Biol. 2002;22:1018–1022. doi: 10.1161/01.atv.0000017470.08363.ab

6. Antón AI, Teruel R, Corral J, Miñano A, Martínez-Martínez I, Ordóñez A, Vicente V, Sánchez-Vega B. Functional consequences of the prothrombotic SERPINC1 rs2227589 polymorphism on antithrombin levels. Haematologica. 2009;94:589–592. doi: 10.3324/haematol.2008.000604

7. Shamsher MK, Chuzhanova NA, Friedman B, Scopes DA, Alhaq A, Millar DS, Cooper DN, Berg LP. Identification of an intronic regulatory element in the human protein C (PROC) gene. Hum Genet. 2000;107:458–465. doi: 10.1007/s004390000391

8. Leroy-Matheron C, Duchemin J, Levent M, Gouault-Heilmann M. Genetic modulation of plasma protein S levels by two frequent dimorphisms in the PROS1 gene. Thromb Haemost. 1999;82:1088–1092.

9. El-Galaly TC, Severinsen MT, Overvad K, Steffensen R, Vistisen AK, Tjønneland A, Kristensen SR. Single nucleotide polymorphisms and the risk of venous thrombosis: results from a Danish case-cohort study. Br J Haematol. 2013;160:838–841. doi: 10.1111/bjh.12132

10. Grundy CB, Chisholm M, Kakkar VV, Cooper DN. A novel homozygous missense mutation in the protein C (PROC) gene causing recurrent venous thrombosis. Hum Genet. 1992;89:683–684. doi: 10.1007/bf00221963

11. Millar DS, Grundy CB, Bignell P, Mitchell DC, Corden D, Woods P, Kakkar VV, Cooper DN. A novel nonsense mutation in the protein C (PROC) gene (Trp-29-->term) causing recurrent venous thrombosis. Hum Genet. 1993;91:196. doi: 10.1007/bf00222726

12. Wu D, Zhong Z, Chen Y, Ding H, Yang M, Lian N, Huang Z, Zhang Q, Zhao J, Deng C. Analysis of PROC and PROS1 single nucleotide polymorphisms in a thrombophilia family. Clin Respir J. 2019;13:530–537. doi: 10.1111/crj.13055

13. de la Morena-Barrio ME, Buil A, Antón AI, Martínez-Martínez I, Miñano A, Gutiérrez-Gallego R, Navarro-Fernández J, Aguila S, Souto JC, Vicente V, et al. Identification of antithrombin-modulating genes. Role of LARGE, a gene encoding a bifunctional glycosyltransferase, in the secretion of proteins? PLoS One. 2013;8:e64998. doi: 10.1371/journal.pone.0064998

14. Oudot-Mellakh T, Cohen W, Germain M, Saut N, Kallel C, Zelenika D, Lathrop M, Trégouët DA, Morange PE. Genome wide association study for plasma levels of natural anticoagulant inhibitors and protein C anticoagulant pathway: the MARTHA project. Br J Haematol. 2012;157:230–239. doi: 10.1111/j.1365-2141.2011.09025.x

15. Tang W, Basu S, Kong X, Pankow JS, Aleksic N, Tan A, Cushman M, Boerwinkle E, Folsom AR. Genome-wide association study identifies novel loci for plasma levels of protein C: the ARIC study. Blood. 2010;116:5032–5036. doi: 10.1182/blood-2010-05-283739

16. Pankow JS, Tang W, Pankratz N, Guan W, Weng LC, Cushman M, Boerwinkle E, Folsom AR. Identification of Genetic Variants Linking Protein C and Lipoprotein Metabolism: The ARIC Study (Atherosclerosis Risk in Communities). Arterioscler Thromb Vasc Biol. 2017;37:589–597. doi: 10.1161/atvbaha.116.308109

17. Munir MS, Weng LC, Tang W, Basu S, Pankow JS, Matijevic N, Cushman M, Boerwinkle E, Folsom AR. Genetic markers associated with plasma protein C level in African Americans: the atherosclerosis risk in communities (ARIC) study. Genet Epidemiol. 2014;38:709–713. doi: 10.1002/gepi.21868

18. Athanasiadis G, Buil A, Souto JC, Borrell M, López S, Martinez-Perez A, Lathrop M, Fontcuberta J, Almasy L, Soria JM. A genome-wide association study of the Protein C anticoagulant pathway. PLoS One. 2011;6:e29168. doi: 10.1371/journal.pone.0029168

19. Ferkingstad E, Sulem P, Atlason BA, Sveinbjornsson G, Magnusson MI, Styrmisdottir EL, Gunnarsdottir K, Helgason A, Oddsson A, Halldorsson BV, et al. Large-scale integration of the plasma proteome with genetics and disease. Nat Genet. 2021;53:1712–1721. doi: 10.1038/s41588-021-00978-w

20. Psaty BM, O’Donnell CJ, Gudnason V, Lunetta KL, Folsom AR, Rotter JI, Uitterlinden AG, Harris TB, Witteman JC, Boerwinkle E. Cohorts for Heart and Aging Research in Genomic Epidemiology (CHARGE) Consortium: Design of prospective meta-analyses of genome-wide association studies from 5 cohorts. Circ Cardiovasc Genet. 2009;2:73–80. doi: 10.1161/circgenetics.108.829747

21. Investigators A. The Atherosclerosis risk in COMMUNIT (ARIC) study: design and objectives. American journal of epidemiology. 1989;129:687–702.

22. Pattaro C, Gögele M, Mascalzoni D, Melotti R, Schwienbacher C, De Grandi A, Foco L, D’Elia Y, Linder B, Fuchsberger C, et al. The Cooperative Health Research in South Tyrol (CHRIS) study: rationale, objectives, and preliminary results. J Transl Med. 2015;13:348. doi: 10.1186/s12967-015-0704-9

23. Fried LP, Borhani NO, Enright P, Furberg CD, Gardin JM, Kronmal RA, Kuller LH, Manolio TA, Mittelmark MB, Newman A, et al. The Cardiovascular Health Study: design and rationale. Ann Epidemiol. 1991;1:263–276. doi: 10.1016/1047-2797(91)90005-w

24. Desch K, Li J, Kim S, Laventhal N, Metzger K, Siemieniak D, Ginsburg D. Analysis of informed consent document utilization in a minimal-risk genetic study. Annals of internal medicine. 2011;155:316–322.

25. Desch KC, Ozel AB, Siemieniak D, Kalish Y, Shavit JA, Thornburg CD, Sharathkumar AA, McHugh CP, Laurie CC, Crenshaw A, et al. Linkage analysis identifies a locus for plasma von Willebrand factor undetected by genome-wide association. Proc Natl Acad Sci U S A. 2013;110:588–593. doi: 10.1073/pnas.1219885110

26. Souto JC, Almasy L, Borrell M, Blanco-Vaca F, Mateo J, Soria JM, Coll I, Felices R, Stone W, Fontcuberta J, et al. Genetic susceptibility to thrombosis and its relationship to physiological risk factors: the GAIT study. Genetic Analysis of Idiopathic Thrombophilia. Am J Hum Genet. 2000;67:1452–1459. doi: 10.1086/316903

27. Psaty BM, Heckbert SR, Koepsell TD, Siscovick DS, Raghunathan TE, Weiss NS, Rosendaal FR, Lemaitre RN, Smith NL, Wahl PW, et al. The risk of myocardial infarction associated with antihypertensive drug therapies. Jama. 1995;274:620–625.

28. Tomaschitz A, Pilz S, Ritz E, Meinitzer A, Boehm BO, März W. Plasma aldosterone levels are associated with increased cardiovascular mortality: the Ludwigshafen Risk and Cardiovascular Health (LURIC) study. European heart journal. 2010;31:1237–1247.

29. Antoni G, Oudot-Mellakh T, Dimitromanolakis A, Germain M, Cohen W, Wells P, Lathrop M, Gagnon F, Morange PE, Tregouet DA. Combined analysis of three genome-wide association studies on vWF and FVIII plasma levels. BMC Med Genet. 2011;12:102. doi: 10.1186/1471-2350-12-102

30. Vázquez-Santiago M, Vilalta N, Cuevas B, Murillo J, Llobet D, Macho R, Pujol-Moix N, Carrasco M, Mateo J, Fontcuberta J, et al. Short closure time values in PFA-100® are related to venous thrombotic risk. Results from the RETROVE Study. Thromb Res. 2018;169:57–63. doi: 10.1016/j.thromres.2018.07.012

31. Mills JL, Carter TC, Scott JM, Troendle JF, Gibney ER, Shane B, Kirke PN, Ueland PM, Brody LC, Molloy AM. Do high blood folate concentrations exacerbate metabolic abnormalities in people with low vitamin B-12 status? The American journal of clinical nutrition. 2011;94:495–500.

32. Kowalski MH, Qian H, Hou Z, Rosen JD, Tapia AL, Shan Y, Jain D, Argos M, Arnett DK, Avery C, et al. Use of >100,000 NHLBI Trans-Omics for Precision Medicine (TOPMed) Consortium whole genome sequences improves imputation quality and detection of rare variant associations in admixed African and Hispanic/Latino populations. PLoS Genet. 2019;15:e1008500. doi: 10.1371/journal.pgen.1008500

33. Winkler TW, Day FR, Croteau-Chonka DC, Wood AR, Locke AE, Mägi R, Ferreira T, Fall T, Graff M, Justice AE, et al. Quality control and conduct of genome-wide association meta-analyses. Nat Protoc. 2014;9:1192–1212. doi: 10.1038/nprot.2014.071

34. Pulit SL, de With SA, de Bakker PI. Resetting the bar: Statistical significance in whole-genome sequencing-based association studies of global populations. Genet Epidemiol. 2017;41:145–151. doi: 10.1002/gepi.22032

35. Yang J, Ferreira T, Morris AP, Medland SE, Madden PA, Heath AC, Martin NG, Montgomery GW, Weedon MN, Loos RJ, et al. Conditional and joint multiple-SNP analysis of GWAS summary statistics identifies additional variants influencing complex traits. Nat Genet. 2012;44:369-375, s361-363. doi: 10.1038/ng.2213

36. Yang J, Lee SH, Goddard ME, Visscher PM. GCTA: a tool for genome-wide complex trait analysis. Am J Hum Genet. 2011;88:76–82. doi: 10.1016/j.ajhg.2010.11.011

37. Barbeira AN, Dickinson SP, Bonazzola R, Zheng J, Wheeler HE, Torres JM, Torstenson ES, Shah KP, Garcia T, Edwards TL, et al. Exploring the phenotypic consequences of tissue specific gene expression variation inferred from GWAS summary statistics. Nat Commun. 2018;9:1825. doi: 10.1038/s41467-018-03621-1

38. Barbeira AN, Pividori M, Zheng J, Wheeler HE, Nicolae DL, Im HK. Integrating predicted transcriptome from multiple tissues improves association detection. PLoS Genet. 2019;15:e1007889. doi: 10.1371/journal.pgen.1007889

39. Ray D, Boehnke M. Methods for meta-analysis of multiple traits using GWAS summary statistics. Genet Epidemiol. 2018;42:134–145. doi: 10.1002/gepi.22105

40. Benjamin DJ, Berger JO, Johannesson M, Nosek BA, Wagenmakers EJ, Berk R, Bollen KA, Brembs B, Brown L, Camerer C, et al. Redefine statistical significance. Nat Hum Behav. 2018;2:6–10. doi: 10.1038/s41562-017-0189-z

41. Lindström S, Wang L, Smith EN, Gordon W, van Hylckama Vlieg A, de Andrade M, Brody JA, Pattee JW, Haessler J, Brumpton BM, et al. Genomic and transcriptomic association studies identify 16 novel susceptibility loci for venous thromboembolism. Blood. 2019;134:1645–1657. doi: 10.1182/blood.2019000435

42. Klarin D, Busenkell E, Judy R, Lynch J, Levin M, Haessler J, Aragam K, Chaffin M, Haas M, Lindström S, et al. Genome-wide association analysis of venous thromboembolism identifies new risk loci and genetic overlap with arterial vascular disease. Nat Genet. 2019;51:1574–1579. doi: 10.1038/s41588-019-0519-3

43. Klarin D, Lynch J, Aragam K, Chaffin M, Assimes TL, Huang J, Lee KM, Shao Q, Huffman JE, Natarajan P, et al. Genome-wide association study of peripheral artery disease in the Million Veteran Program. Nat Med. 2019;25:1274–1279. doi: 10.1038/s41591-019-0492-5

44. Nikpay M, Goel A, Won HH, Hall LM, Willenborg C, Kanoni S, Saleheen D, Kyriakou T, Nelson CP, Hopewell JC, et al. A comprehensive 1,000 Genomes-based genome-wide association meta-analysis of coronary artery disease. Nat Genet. 2015;47:1121–1130. doi: 10.1038/ng.3396

45. Malik R, Chauhan G, Traylor M, Sargurupremraj M, Okada Y, Mishra A, Rutten-Jacobs L, Giese AK, van der Laan SW, Gretarsdottir S, et al. Multiancestry genome-wide association study of 520,000 subjects identifies 32 loci associated with stroke and stroke subtypes. Nat Genet. 2018;50:524–537. doi: 10.1038/s41588-018-0058-3

46. Stacey D, Chen L, Howson JM, Mason AM, Burgess S, MacDonald S, Langdown J, McKinney H, Downes K, Farahi N. Elucidating mechanisms of genetic cross-disease associations: an integrative approach implicates protein C as a causal pathway in arterial and venous diseases. medRxiv. 2020.

47. Stacey D, Chen L, Stanczyk PJ, Howson JM, Mason AM, Burgess S, MacDonald S, Langdown J, McKinney H, Downes K. Elucidating mechanisms of genetic cross-disease associations at the PROCR vascular disease locus. Nature communications. 2022;13:1–15.

48. Temprano-Sagrera G, Sitlani CM, Bone WP, Martin-Bornez M, Voight BF, Morrison AC, Damrauer SM, de Vries PS, Smith NL, Sabater-Lleal M. Multi-phenotype analyses of hemostatic traits with cardiovascular events reveal novel genetic associations. J Thromb Haemost. 2022;20:1331–1349. doi: 10.1111/jth.15698

49. Manderstedt E, Lind-Halldén C, Halldén C, Elf J, Svensson PJ, Dahlbäck B, Engström G, Melander O, Baras A, Lotta LA, et al. Classic Thrombophilias and Thrombotic Risk Among Middle-Aged and Older Adults: A Population-Based Cohort Study. J Am Heart Assoc. 2022;11:e023018. doi: 10.1161/jaha.121.023018

50. Daly M, O’Meara A, Hallinan FM. Identification and characterization of a new antithrombin III familial variant (AT Dublin) with possible increased frequency in children with cancer. Br J Haematol. 1987;65:457–462. doi: 10.1111/j.1365-2141.1987.tb04150.x

51. Daly M, Bruce D, Perry DJ, Price J, Harper PL, O’Meara A, Carrell RW. Antithrombin Dublin (−3 Val----Glu): an N-terminal variant which has an aberrant signal peptidase cleavage site. FEBS Lett. 1990;273:87–90. doi: 10.1016/0014-5793(90)81057-u

52. Dürr C, Hinney A, Luckenbach C, Kömpf J, Ritter H. Genetic studies of antithrombin III with IEF and ASO hybridization. Hum Genet. 1992;90:457–459. doi: 10.1007/bf00220477

53. Downes K, Megy K, Duarte D, Vries M, Gebhart J, Hofer S, Shamardina O, Deevi SVV, Stephens J, Mapeta R, et al. Diagnostic high-throughput sequencing of 2396 patients with bleeding, thrombotic, and platelet disorders. Blood. 2019;134:2082–2091. doi: 10.1182/blood.2018891192

54. Stockinger W, Sailler B, Strasser V, Recheis B, Fasching D, Kahr L, Schneider WJ, Nimpf J. The PX-domain protein SNX17 interacts with members of the LDL receptor family and modulates endocytosis of the LDL receptor. Embo j. 2002;21:4259–4267. doi: 10.1093/emboj/cdf435

55. Zhao D, Li X, Liang H, Zheng N, Pan Z, Zhou Y, Liu X, Qian M, Xu B, Zhang Y, et al. SNX17 produces anti-arrhythmic effects by preserving functional SERCA2a protein in myocardial infarction. Int J Cardiol. 2018;272:298–305. doi: 10.1016/j.ijcard.2018.07.025

56. Yang J, Villar VAM, Rozyyev S, Jose PA, Zeng C. The emerging role of sorting nexins in cardiovascular diseases. Clin Sci (Lond). 2019;133:723–737. doi: 10.1042/cs20190034

57. Smith NL, Chen MH, Dehghan A, Strachan DP, Basu S, Soranzo N, Hayward C, Rudan I, Sabater-Lleal M, Bis JC, et al. Novel associations of multiple genetic loci with plasma levels of factor VII, factor VIII, and von Willebrand factor: The CHARGE (Cohorts for Heart and Aging Research in Genome Epidemiology) Consortium. Circulation. 2010;121:1382–1392. doi: 10.1161/circulationaha.109.869156

58. Sennblad B, Basu S, Mazur J, Suchon P, Martinez-Perez A, van Hylckama Vlieg A, Truong V, Li Y, Gådin JR, Tang W, et al. Genome-wide association study with additional genetic and post-transcriptional analyses reveals novel regulators of plasma factor XI levels. Hum Mol Genet. 2017;26:637–649. doi: 10.1093/hmg/ddw401

59. Ellis J, Lange EM, Li J, Dupuis J, Baumert J, Walston JD, Keating BJ, Durda P, Fox ER, Palmer CD, et al. Large multiethnic Candidate Gene Study for C-reactive protein levels: identification of a novel association at CD36 in African Americans. Hum Genet. 2014;133:985–995. doi: 10.1007/s00439-014-1439-z

60. Folsom AR, Lutsey PL, Astor BC, Cushman M. C-reactive protein and venous thromboembolism. A prospective investigation in the ARIC cohort. Thromb Haemost. 2009;102:615–619. doi: 10.1160/th09-04-0274

61. Santoro N, Zhang CK, Zhao H, Pakstis AJ, Kim G, Kursawe R, Dykas DJ, Bale AE, Giannini C, Pierpont B, et al. Variant in the glucokinase regulatory protein (GCKR) gene is associated with fatty liver in obese children and adolescents. Hepatology. 2012;55:781–789. doi: 10.1002/hep.24806

62. Petit JM, Masson D, Guiu B, Rollot F, Duvillard L, Bouillet B, Brindisi MC, Buffier P, Hillon P, Cercueil JP, et al. GCKR polymorphism influences liver fat content in patients with type 2 diabetes. Acta Diabetol. 2016;53:237–242. doi: 10.1007/s00592-015-0766-4

63. Yeh KH, Hsu LA, Teng MS, Wu S, Chou HH, Ko YL. Pleiotropic Effects of Common and Rare GCKR Exonic Mutations on Cardiometabolic Traits. Genes (Basel). 2022;13. doi: 10.3390/genes13030491

64. Ligthart S, Vaez A, Võsa U, Stathopoulou MG, de Vries PS, Prins BP, Van der Most PJ, Tanaka T, Naderi E, Rose LM, et al. Genome Analyses of >200,000 Individuals Identify 58 Loci for Chronic Inflammation and Highlight Pathways that Link Inflammation and Complex Disorders. Am J Hum Genet. 2018;103:691–706. doi: 10.1016/j.ajhg.2018.09.009

65. Wejman JC, Hovsepian D, Wall JS, Hainfeld JF, Greer J. Structure of haptoglobin and the haptoglobin-hemoglobin complex by electron microscopy. J Mol Biol. 1984;174:319–341. doi: 10.1016/0022-2836(84)90341-3

66. Landis RC, Philippidis P, Domin J, Boyle JJ, Haskard DO. Haptoglobin Genotype-Dependent Anti-Inflammatory Signaling in CD163(+) Macrophages. Int J Inflam. 2013;2013:980327. doi: 10.1155/2013/980327

67. Asleh R, Miller-Lotan R, Aviram M, Hayek T, Yulish M, Levy JE, Miller B, Blum S, Milman U, Shapira C, et al. Haptoglobin genotype is a regulator of reverse cholesterol transport in diabetes in vitro and in vivo. Circ Res. 2006;99:1419–1425. doi: 10.1161/01.Res.0000251741.65179.56

68. Vormittag R, Vukovich T, Mannhalter C, Minar E, Schönauer V, Bialonczyk C, Hirschl M, Pabinger I. Haptoglobin phenotype 2-2 as a potentially new risk factor for spontaneous venous thromboembolism. Haematologica. 2005;90:1557–1561.

69. Miller YI, Altamentova SM, Shaklai N. Oxidation of low-density lipoprotein by hemoglobin stems from a heme-initiated globin radical: antioxidant role of haptoglobin. Biochemistry. 1997;36:12189–12198. doi: 10.1021/bi970258a

70. Rota S, McWilliam NA, Baglin TP, Byrne CD. Atherogenic lipoproteins support assembly of the prothrombinase complex and thrombin generation: modulation by oxidation and vitamin E. Blood. 1998;91:508–515.

71. Kumagai T, Hoshi Y, Tsutsumi H, Ebina K, Yokota K. Inhibition of plasma coagulation through interaction between oxidized low-density lipoprotein and blood coagulation factor VIII. Biol Pharm Bull. 2005;28:952–956. doi: 10.1248/bpb.28.952

72. Mikulska JE, Pablo L, Canel J, Simister NE. Cloning and analysis of the gene encoding the human neonatal Fc receptor. Eur J Immunogenet. 2000;27:231–240. doi: 10.1046/j.1365-2370.2000.00225.x

73. Cejas RB, Ferguson DC, Quiñones-Lombraña A, Bard JE, Blanco JG. Contribution of DNA methylation to the expression of FCGRT in human liver and myocardium. Sci Rep. 2019;9:8674. doi: 10.1038/s41598-019-45203-1

74. Pyzik M, Rath T, Kuo TT, Win S, Baker K, Hubbard JJ, Grenha R, Gandhi A, Krämer TD, Mezo AR, et al. Hepatic FcRn regulates albumin homeostasis and susceptibility to liver injury. Proc Natl Acad Sci U S A. 2017;114:E2862–e2871. doi: 10.1073/pnas.1618291114

75. Hoffmann TJ, Theusch E, Haldar T, Ranatunga DK, Jorgenson E, Medina MW, Kvale MN, Kwok PY, Schaefer C, Krauss RM, et al. A large electronic-health-record-based genome-wide study of serum lipids. Nat Genet. 2018;50:401–413. doi: 10.1038/s41588-018-0064-5

76. Klarin D, Damrauer SM, Cho K, Sun YV, Teslovich TM, Honerlaw J, Gagnon DR, DuVall SL, Li J, Peloso GM, et al. Genetics of blood lipids among ∼300,000 multi-ethnic participants of the Million Veteran Program. Nat Genet. 2018;50:1514–1523. doi: 10.1038/s41588-018-0222-9

77. Astle WJ, Elding H, Jiang T, Allen D, Ruklisa D, Mann AL, Mead D, Bouman H, Riveros-Mckay F, Kostadima MA, et al. The Allelic Landscape of Human Blood Cell Trait Variation and Links to Common Complex Disease. Cell. 2016;167:1415-1429.e1419. doi: 10.1016/j.cell.2016.10.042

78. Halpain S, Dehmelt L. The MAP1 family of microtubule-associated proteins. Genome Biol. 2006;7:224. doi: 10.1186/gb-2006-7-6-224

79. Dente L, Pizza MG, Metspalu A, Cortese R. Structure and expression of the genes coding for human alpha 1-acid glycoprotein. Embo j. 1987;6:2289–2296. doi: 10.1002/j.1460-2075.1987.tb02503.x

80. Rocanin-Arjo A, Cohen W, Carcaillon L, Frère C, Saut N, Letenneur L, Alhenc-Gelas M, Dupuy AM, Bertrand M, Alessi MC, et al. A meta-analysis of genome-wide association studies identifies ORM1 as a novel gene controlling thrombin generation potential. Blood. 2014;123:777–785. doi: 10.1182/blood-2013-10-529628

81. Lopez S, Martinez-Perez A, Rodriguez-Rius A, Viñuela A, Brown AA, Martin-Fernandez L, Vilalta N, Arús M, Panousis NI, Buil A, et al. Integrated GWAS and Gene Expression Suggest ORM1 as a Potential Regulator of Plasma Levels of Cell-Free DNA and Thrombosis Risk. Thromb Haemost. 2022;122:1027–1039. doi: 10.1055/s-0041-1742169

82. Otagiri M, Maruyama T, Imai T, Suenaga A, Imamura Y. A comparative study of the interaction of warfarin with human alpha 1-acid glycoprotein and human albumin. J Pharm Pharmacol. 1987;39:416–420. doi: 10.1111/j.2042-7158.1987.tb03412.x

83. Wadelius M, Chen LY, Eriksson N, Bumpstead S, Ghori J, Wadelius C, Bentley D, McGinnis R, Deloukas P. Association of warfarin dose with genes involved in its action and metabolism. Hum Genet. 2007;121:23–34. doi: 10.1007/s00439-006-0260-8

84. Bourgeois S, Jorgensen A, Zhang EJ, Hanson A, Gillman MS, Bumpstead S, Toh CH, Williamson P, Daly AK, Kamali F, et al. A multi-factorial analysis of response to warfarin in a UK prospective cohort. Genome Med. 2016;8:2. doi: 10.1186/s13073-015-0255-y

85. Morano I, Hofmann F, Zimmer M, Rüegg JC. The influence of P-light chain phosphorylation by myosin light chain kinase on the calcium sensitivity of chemically skinned heart fibres. FEBS Lett. 1985;189:221–224. doi: 10.1016/0014-5793(85)81027-9

86. Himpens B, Matthijs G, Somlyo AV, Butler TM, Somlyo AP. Cytoplasmic free calcium, myosin light chain phosphorylation, and force in phasic and tonic smooth muscle. J Gen Physiol. 1988;92:713–729. doi: 10.1085/jgp.92.6.713

87. Vujkovic M, Keaton JM, Lynch JA, Miller DR, Zhou J, Tcheandjieu C, Huffman JE, Assimes TL, Lorenz K, Zhu X, et al. Discovery of 318 new risk loci for type 2 diabetes and related vascular outcomes among 1.4 million participants in a multi-ancestry meta-analysis. Nat Genet. 2020;52:680–691. doi: 10.1038/s41588-020-0637-y

88. Chung RH, Chiu YF, Wang WC, Hwu CM, Hung YJ, Lee IT, Chuang LM, Quertermous T, Rotter JI, Chen YI, et al. Multi-omics analysis identifies CpGs near G6PC2 mediating the effects of genetic variants on fasting glucose. Diabetologia. 2021;64:1613–1625. doi: 10.1007/s00125-021-05449-9

89. Mahmoodi BK, Brouwer JL, Ten Kate MK, Lijfering WM, Veeger NJ, Mulder AB, Kluin-Nelemans HC, Van Der Meer J. A prospective cohort study on the absolute risks of venous thromboembolism and predictive value of screening asymptomatic relatives of patients with hereditary deficiencies of protein S, protein C or antithrombin. J Thromb Haemost. 2010;8:1193–1200. doi: 10.1111/j.1538-7836.2010.03840.x

90. Pabinger I, Kyrle PA, Heistinger M, Eichinger S, Wittmann E, Lechner K. The risk of thromboembolism in asymptomatic patients with protein C and protein S deficiency: a prospective cohort study. Thromb Haemost. 1994;71:441–445.

91. Schooling CM, Zhong Y. Plasma levels of the anti-coagulation protein C and the risk of ischaemic heart disease. A Mendelian randomisation study. Thromb Haemost. 2017;117:262–268. doi: 10.1160/th16-07-0518

92. O’Connor NT, Broekmans AW, Bertina RM. Protein C values in coronary artery disease. Br Med J (Clin Res Ed). 1984;289:1192. doi: 10.1136/bmj.289.6453.1192

93. Wessler S, Gaston LW. Anticoagulant therapy in coronary artery disease. Circulation. 1966;34:856–864. doi: 10.1161/01.cir.34.5.856

94. Wilbur J, Shian B. Deep Venous Thrombosis and Pulmonary Embolism: Current Therapy. Am Fam Physician. 2017;95:295–302.

95. Kuchenbaecker K, Telkar N, Reiker T, Walters RG, Lin K, Eriksson A, Gurdasani D, Gilly A, Southam L, Tsafantakis E, et al. The transferability of lipid loci across African, Asian and European cohorts. Nat Commun. 2019;10:4330. doi: 10.1038/s41467-019-12026-7

96. Evans DS, Avery CL, Nalls MA, Li G, Barnard J, Smith EN, Tanaka T, Butler AM, Buxbaum SG, Alonso A, et al. Fine-mapping, novel loci identification, and SNP association transferability in a genome-wide association study of QRS duration in African Americans. Hum Mol Genet. 2016;25:4350–4368. doi: 10.1093/hmg/ddw284

97. Adeyemo A, Bentley AR, Meilleur KG, Doumatey AP, Chen G, Zhou J, Shriner D, Huang H, Herbert A, Gerry NP, et al. Transferability and fine mapping of genome-wide associated loci for lipids in African Americans. BMC Med Genet. 2012;13:88. doi: 10.1186/1471-2350-13-88

