## Supplementary for "Antithrombin, protein C and protein S: Genome and transcriptome wide association studies identify 7 novel loci regulating plasma levels"

**Supplemental Materials**

**Section	Page**

Supplemental Methods: Genome-wide Association Studies	2

Supplemental Methods: Functional Studies	4

Supplemental Methods: Mendelian Randomization Analyses	5

Cohort-Specific Information	6

Supplemental Table S1	10

Supplemental Table S2	12

Supplemental Table S3	15

Supplemental Table S4	16

Supplemental Table S5	17

Supplemental Table S6	21

Supplemental Table S7	22

Supplemental Table S8	23

Supplemental Table S9	24

Supplemental Table S10	26

Supplemental Table S11	27

Supplemental Table S12	28

Supplemental Table S13	29

Supplemental Figure S1	30

Supplemental Figure S2	31

Supplemental Figure S3	32

Supplemental Figure S4	33

Supplemental Figure S5	34

Supplemental Figure S6	35

Funding and Acknowledgements from Specific Cohorts	36

References for Supplement	38

**Supplementary Methods:**

**Genome-wide Association Studies (GWAS)**

**Meta-level quality control and meta-analysis**

We used the EasyQC software to perform basic quality control and homogenization of the data. SE-N (standard error against sample size) plots were used to reveal inconsistencies of phenotype transformations or units between studies. Allele frequencies were plotted against ancestry-specific allele reference panels from TOPMed to uncover issues of strand and allele coding. Lambda was calculated for all studies individually to detect inflation of p-values caused by problems of stratification. All variants with inconsistencies, extreme beta, or standard error values and imputation values < 0.3 were removed prior to meta-analysis. We also excluded variants that had missing values of coefficients, frequency, and P-value.

**Meta-analysis**

We meta-analyzed the 22 autosomal chromosomes and the X chromosome separately, using fixed-effects inverse variance or sample size methods implemented in the METAL software^1^. Meta-analyses within each phenotyping method were adjusted for the genomic control coefficients (λ_GC_) of each individual cohort. Summarized coefficients and p-values for each phenotype were presented as the result. Genome-wide association significance level was set as 5x10^-9^.

**Calculation of the Proportion of variance explained (r^2^)**

The proportion of variance explained by the associated variants was calculated for each ethnicity using the variants identified in the cross-ancestry meta-analysis with the following formula: r^2^ = 2*EAF(1 - EAF)*𝛽2 / var(y); where EAF and 𝛽 represent the frequency and the per-allele increment in phenotype of the effect allele, respectively, and var(y) represents the phenotypic variance. We defined var(y) as the square of the weighted mean of standard deviations across all the cohorts that contributed to each ethnic specific meta-analysis.

**Transcription-wide association studies (TWAS)**

We used the EA population from the 1000 Genomes project^2^ for the imputation of the missing variants in our GWAS input and for linkage disequilibrium (LD) blocks definition. Data from the Genotype-Tissue Expression (GTEx)^3^ V8 database was used as reference to build the prediction models for cis-eQTL sites using the mash-R method (v0.2.5)^4-7^. Weights computed with mash-R were then applied to the final antithrombin, PC, and PS GWAS meta-analyses summary statistics to identify gene-trait associations.

S-PrediXcan was run to identify gene-trait associations for each tissue, and then the significantly shared eQTL across the 5 chosen tissues were leveraged using S-MultiXcan analyses following the pipelines described in previous studies^5,8^.

**Multi-phenotype analyses**

MetaUSAT meta-analyzes summary statistics of common variants between multiple phenotypes, returning a p-value for each variant (p-value multivariate). The alleles of all datasets were previously harmonized before running the analysis and variants with minor allele count (MAC) < 30 were removed prior to the analysis. The significance threshold used was set at 5 x 10-9 and loci were defined as +/- 1 Mb around the variants with the lowest p-value. Previous GWAS results for antithrombin, PC, PS free and total were queried using the GWAS catalog database (available at <https://www.ebi.ac.uk/gwas/docs/file-downloads>).

**Fine Mapping**

We used FOCUS (fine mapping of causal gene sets)^9^ to prioritize TWAS genes in each genome-wide significant locus by estimating credible gene sets, avoiding confounding caused by LD or horizontal pleiotropy between variants that normally affect TWAS. Summary data from the EA meta-analyses for antithrombin, PC and PS, and expression prediction weights from PrediXcan GTEx v8 MASH-R models were analyzed following software recommendations. Reference LD was estimated using the PROCARDIS database, including 5,820 unrelated EA individuals with genome-wide genotyping data imputed to the TOPMed v5 panel. For fine-mapping purposes, only genes within approximately 1Mb regions (obtained from LDetect software^10^ using EA 1000 Genomes - b38) that contained at least 1 variant in the GWAS summary statistics with p-value < 5 x 10−8 were considered as recommended in the software pipe-line. This strategy was used to prioritize putative causal genes by providing a posterior inclusion probability (PIP) for each gene per locus. The PIP was interpreted as the probability of a gene to be causal given the TWAS statistics.

**Colocalization Analyses**

Colocalization analyses were performed for all the new genome-wide significant associations detected in the GWAS (cross-ancestry and EA) analyses using the COLOC R package V5.1.0.1.^11^ GWAS summary statistics in each associated locus were subjected to a colocalization analysis with RNAseq data from the Genotype-Tissue Expression (GTEx) project^5^ to seek for evidence of shared variants affecting both gene expression levels of closest genes (eQTLs) and the levels of each anticoagulant protein. We used the complete GTEx V8 files (available at https://console.cloud.google.com/storage/browser/gtex-resources) and considered windows of +/- 1 Mb around the lead variants to run the colocalization. We required that the conditional probability of colocalization (CPC) for a variant, given that the 2 phenotypes showed evidence of association (PPH4 / (PPH3 + PPH4)) was higher than 0.8.

We also performed colocalization analyses in all novel loci identified in the multi-phenotype analysis. First, we performed colocalization across multiple traits using COLOC (for 2 phenotypes)^11^, or HyPrColoc^12^ (for more than 2 phenotypes) R-packages, to identify loci regulating more than one anticoagulant phenotype. We also required a CPC higher than 0.8 for COLOC, or a probability of colocalization higher than 0.7 for HyPrColoc. Second, to help prioritize the causal gene in each locus, we tested colocalization with expression data at the novel loci derived from the multi-phenotype analyses, using HyPrColoc and the complete GTEx V8 files. A probability of colocalization > 0.7 was also required. As in TWAS analysis, we only considered functional related tissues to reduce false positives triggered by non-related tissues.

**Functional Studies**

**Cell Culture and Gene Silencing**

HepG2 cells were obtained from the UNC Lineberger Comprehensive Cancer Center Tissue Culture Facility and cultured in Minimum Essential Media (MEM, catalog #11095-080, Gibco, Waltham, MA) supplemented with 10% FBS (VWR, Radnor, PA), 1 mM sodium pyruvate, and 0.1 mM non-essential amino acids (Gibco). Cells were passaged using 0.25% Trypsin-EDTA (catalog #25200-056, Gibco). Cells from passage 135-145 were used for transfection experiments. Small interfering RNA (siRNAs, Silencer Select siRNAs, Thermo Fisher Scientific, Waltham, MA) were complexed with Lipofectamine RNAi MAX (Thermo Fisher Scientific) in OptiMEM Reduced Serum Media (RSM, catalog #31985-070, Gibco) for 15 minutes before adding to a 24-well plate. Cells were passaged, resuspended in MEM supplemented with 10% FBS, 1 mM sodium pyruvate, 0.1 mM non-essential amino acids, and added to the transfection complex (100,000-150,000 cells/well and 32 nM siRNA, final). After 48 hours, media was replaced with fresh OptiMEM RSM and antithrombin-enriched supernatants were collected 24 hours later.

**RNA Extraction and Quantitative RT-PCR**

Total RNA was extracted from transfected HepG2 cells using the RNeasy Mini Kit (Qiagen, German Town, MD). RNA concentration was determined using a NanoDrop 2000 (Thermo Fisher Scientific). cDNA was prepared from 1 µg RNA using the QuantiTect Reverse Transcription Kit (Qiagen). Real Time quantitative PCR (RT-qPCR) was performed using the QuantiFast SYBR Green RT-PCR kit (Qiagen) and Quant Studio 3 (Thermofisher), with activation (95°C, 5 minutes), denaturation (95°C, 10 seconds), and annealing/extension (60°C, 30 seconds) for 40 cycles. RNA18S was used as the housekeeping gene for relative expression calculations.

**Immunoblotting**

Proteins were separated on 10% gels by reducing SDS-PAGE and transferred to PVDF membranes. Membranes were blocked for 1 hour in LICOR Odyssey Blocking Buffer (Lincoln, NE) and incubated overnight with 25 µg/mL sheep anti-human antithrombin (Haematologic Technologies, Essex Junction, VT) diluted in Tris-buffered saline, 0.1% Tween 20 (TBST). Membranes were incubated in LICOR IRDye 800 CW donkey anti-goat secondary antibody (1:10,000 in TBST) for 1 hour at RT. All washes were done using TBST. Antithrombin bands were quantified by fluorescence (Biorad Chemi-Doc MP Imager).

**Cell counts**

HepG2 cells were washed with PBS followed by trypsinization using 0.25% trypsin EDTA and resuspended in culture media. Cells were counted on the Countess II FL cells counter using cell counting chamber slides (Thermo Fisher Scientific).

**Table. Primers used for qPCR.**

| **Primer** | **Forward Sequence** | **Reverse Sequence** |
| --- | --- | --- |
| **PROC** | **CTTGTCAGGCTTGGAGAGTATG** | **GTGGTGCTCTTGCTGTAGTT** |
| **SERPINC1** | **GATGAGGGCTCAGAACAGAAG** | **TGCCAGGTGCTGATAGAAAG** |
| **SNX17** | **TGAGGTAGAACAGAGGAGAGAG** | **GACGCAGGAAACTGTTGAAAG** |
| **NRBP1** | **CACAAATCCTCTCTGCCCTAAG** | **GTCCGTTGTGCTGGATGAA** |
| **GCKR** | **GTGGAGCAGGTGAAAGAGAA** | **TGCTGATGATGGAGGGAAAG** |
| **HP** | **CTGTGCTGGCATGTCTAAGT** | **CTTAAGATCCCAGTCGCATACC** |
| **LCMT2** | **GGCTTCGACTCGCTCTATTT** | **GCGTCTCTCCAATCCTTTCT** |

**Mendelian Randomization Analyses**

Summary statistics for the outcomes were obtained from previously published, large-scale GWAS^13-18^ .To maximize power and ensure African ancestry specific variants were included in our MR analyses for VTE, we performed an inverse variance weighted (IVW) meta-analysis on the results of the latest INVENT (30,234 cases and 172,122 controls) and Million Veteran Program (MVP) (11,844, and 251,951 controls) GWAS of VTE ^16,17^.

IVW was used as the primary analysis and MR-Egger^19-21^, Weighted Median, and Weighted Mode served as secondary analyses. MR Presso was used to test for horizontal pleiotropy and variants showing evidence of pleiotropy were excluded. IVW excluding the ABO locus was also performed for PC despite the locus passing MR Presso because it has been previously associated with several hemostatic phenotypes and therefore fails the horizontal pleiotropy assumption of MR^22^. Genetic instruments for antithrombin and PC were chosen separately by first restricting the exposure and outcome summary statistics to variants they had in common, and then, filtering the genetic variants in the exposure GWAS for genome-wide significance (P-value < 5 x 10^-9^). Finally, the remaining genetic variants for each exposure were distance pruned to select the most significant genetic variants at each locus (+/- 10 Mb for antithrombin to account for the longer disequilibrium pattern of rare variants, and +/- 1 Mb for PC). Known pleiotropic loci would be removed during selection of genetic instruments.

**Cohort-Specific Information**

The **Atherosclerosis Risk In Communities (ARIC)** study is a population-based prospective cohort study of cardiovascular disease sponsored by the National Heart, Lung, and Blood Institute (NHLBI). ARIC included 15,792 individuals, predominantly European American and African American, aged 45-64 years at baseline (1987-89), chosen by probability sampling from four US communities. Cohort members completed three additional triennial follow-up examinations, a fifth exam in 2011-2013, a sixth exam in 2016-2017, and a seventh exam in 2018-2019. The ARIC study has been described in detail previously^23^.

Antithrombin was measured at baseline using an amidolytic assay using a synthetic chromogenic substrate for thrombin (CBS 34.47, Diagnostica Stago) and bovine thrombin (Kabi). Protein C was measured at baseline with a commercial ELISA kit (Asserachrom Protein C, Diagnostica Stago).

The **Cooperative Health Research In South Tyrol** **(CHRIS)** study^24^ is a population-based study with a longitudinal lookout established in 2011 in the Val Venosta/Vinschgau district, South Tyrol, Italy, to investigate the genetic basis of common chronic conditions associated with human ageing. Recruited into the study at baseline between 2011 and 2018 were 13,393 participants from 13 municipalities, each one characterized by a central town, small villages, and scattered mountain farms. Settlements are located at an altitude of 600 to 2,000 m above sea level.

Participants were all adults aged over 18 and cover more than one-third of the target region population. More detailed information about the study design could be found in elsewhere^24^.

After overnight fasting, blood samples were collected between 08:00 AM and 10:00 AM at the study center. After pre-analytical sample processing, samples were shipped to the laboratory of the Merano hospital, Italy. There, two different systems were used to assess hemostatic factors in an aliquot of citrated plasma: 1) ROCHE CA1500 system (between August 2011 and January 2014) and 2) Stago STA Compact max system (between February 2014 and December 2018). Antithrombin was measured with the use of chromogenic methods (Siemens Innovance Antithrombin in ROCHE system and STA-Stachrom AT III in Stago system).

The **Cardiovascular Health Study(CHS)^25^** is a population-based cohort study of risk factors for CHD and stroke in adults ≥65 years conducted across 4 field centers. The original predominantly Caucasian cohort of 5,201 persons was recruited in 1989-1990 from random samples of the Medicare eligibility lists; subsequently, an additional predominantly African American cohort of 687 persons was enrolled for a total sample of 5,888. DNA was extracted from blood samples drawn on all participants at their baseline examination in 1989-90 and all participants provided informed consent for the use of their genetic data in analyses. After an 8-12-h fast, CHS participants underwent phlebotomy by atraumatic venipuncture with a 21-gauge butterfly needle connected to a Vacutainer (Becton Dickinson, Rutherford, NJ) outlet via a Luer adaptor^26^.Antithrombin was measured using a chromogenic assay with coefficient of variation as 7.0%. Protein C was measured by immunoassay with a coefficient of variation as 14.4%. These measures were not abundantly available in African-American participants so analyses were restricted to those of European ancestry.

The **Genes and Blood-clotting Study** **(GABC)** is a cohort study of 1,150 siblings from the University of Michigan, Ann Arbor, during 2006-2009. All participants enrolled in the study were aged between 14 years to 35 years and had at least one eligible healthy sibling, and have signed an online agreement form^27^. Whole blood was obtained at a one-time sampling and rapidly processed into citrated plasma while buffy coats were preserved for DNA extraction. Multiple biochemical phenotypes have been assayed with preserved plasma from GABC participants, primarily focused on measurement of hemostasis and thrombosis related proteins. Participants were also phenotype for bleeding tendency and multiple other common human traits.

Antithrombin was measured by a commercial AlphaLISA kit. When available, plasma concentrations were determined by standard curve fitting to plasma standards provided by George King FACT plasma^28^. https://kingbiomed.com/products/fact-factor-assay-control/

The **Genetic Analysis of Idiopathic Thrombophilia (GAIT)** project is a family-based study where 935 subjects in 35 extended pedigrees were collected^29,30^. To be included in the study, a family was required to have at least 10 living individuals in 3 or more generations. Families were selected through a proband with idiopathic thrombophilia, which was defined as recurrent thrombotic events (at least one of which was spontaneous), a single spontaneous thrombotic episode plus a first-degree relative also affected, or onset of thrombosis before age 45. Thrombosis in these probands was considered idiopathic when biological causes as antithrombin deficiency, protein S and C deficiencies, activated protein C resistance, plasminogen deficiency, heparin cofactor II deficiency, Factor V Leiden, dysfibrinogenemia, lupus anticoagulant and antiphospholipid antibodies, were excluded. Subjects were interviewed by a physician to determine their health and reproductive history, current medications, alcohol consumption, use of sex hormones (oral contraceptives or hormonal replacement therapy) and their smoking history. The study was performed according to the Declaration of Helsinki. All procedures of the study were reviewed by the Institutional Review Board of the Hospital de la Santa Creu i Sant Pau, Barcelona, Spain. Adult subjects gave informed consent for themselves and for their minor children. Antithrombin and Protein C were measured in a biochemical analyzer (Metrolab;RAL,Barcelona, Spain) with the use of chromogenic methods from Chromogenix, an. Functional protein S was assayed with deficient plasma from Diagnostica Stago (Ansières, France) in the STA automated coagulometer (Boehringer Mannheim). Total free Protein S was measured by ELISA from Diagnostica Stago.

The **Heart and Vascular Health** **(HVH)** The Heart and Vascular Health (HVH) Study is a set of population-based, case-control studies evaluating risk factors for cardiovascular disease in the member of Group Health Cooperative, Washington^31-36^.Eligible participants for this study were postmenopausal female HVH study controls, who were randomly-selected GHC enrollees between 2003 and 2010. Among the 1499 women with available blood samples, not history of VTE, and not using anticoagulation, we measured hemostatic measures on users and non-user of hormone therapy. Two hemostatic measurements were performed: [1] antithrombin activity (ATc); and [2] total protein S antigen. For ATc, 100 μl AT was added to 100 μl plasma in a 1/20 dilution with Diluent buffer. After incubation for 60 seconds, 100 μl AT substrate was added and measurement started. All steps were automatically performed by the STA analyser; total protein S antigen levels (CV using commercial quality control: 4.2%) were measured by an enzyme-linked immunosorbent assay (Diagnostica Stago, Asnières, France). (Diagnostica Stago, Asnières, France)^37^.

The **Ludwigshafen Risk and Cardiovascular Health (LURIC)** study^38^ is an ongoing prospective study of more than 3,300 individuals of German ancestry in whom cardiovascular and metabolic phenotypes (coronary artery disease, myocardial infarction, dyslipidemia, hypertension, metabolic syndrome and diabetes mellitus) have been defined or ruled out using standardized methodologies in all study participants. Inclusion criteria for LURIC were: German ancestry (limitation of genetic heterogeneity), clinical stability (except for acute coronary syndromes) and availability of a coronary angiogram. Exclusion criteria were: any acute illness other than acute coronary syndromes, any chronic disease where non-cardiac disease predominated and a history of malignancy within the last five years. Genome-wide analyses using the Affymetrix 6.0 have been completed in all participants. A 10-year clinical follow-up for total and cause specific mortality has been completed.

Antithrombin was determined using STA ANTITHROMBIN III assay (Stago Diagnostica/Roche, Mannheim, Germany), Protein C was determined using the COAMATIC assay on a STA Stago analyser (Chromogenix Instrumentation, Laboratory SpA, Milan, Italy).

The **MARseille THrombosis Association (MARTHA)** project has been previously described^39^. Briefly, MARTHA consists on two independent samples of VT patients, named MARTHA08 (N=1,006) and MARTHA10 (N=586). MARTHA patients are unrelated subjects of European origin, with the majority being of French ancestry, consecutively recruited at the Thrombophilia center of La Timone hospital (Marseille, France) between January 1994 and October 2005. All patients had a documented history of VT and free of well characterized genetic risk factors including AT, PC, or PS deficiency, homozygosity for FV Leiden or FII 20210A, and lupus anticoagulant. They were interviewed by a physician on their medical history, which emphasized manifestations of deep vein thrombosis and pulmonary embolism using a standardized questionnaire. The thrombotic events were confirmed by venography, Doppler ultrasound, spiral computed tomographic scanning angiography, and/or ventilation/perfusion lung scan. Blood samples were collected by antecubital venipuncture into Vacutainer® tubes 0.105 M trisodium citrate (ratio 9:1, Becton Dickinson) for the coagulation tests. Platelet-poor plasma (PPP) was obtained after double centrifugation of citrated blood (3000 g for 10 min at 25°C) and kept frozen at -80°C until analysis.

Antithrombin activity levels were performed using an amidolytic assay (STA-Stachrom ATIII kit; Diagnostica Stago, Asnieres, France). Protein C activity was assayed using a chronometric assay (Instrumentation Laboratory, Paris, France). Quantitative determination of free PS levels was performed by enzyme-linked immunosorbent assay using the Asserachrom f-PS assay (Diagnostica Stago).

The **Riesgo de Enfermedad TROmboembolica VEnosa Study (RETROVE)** is a prospective case–control study that includes 400 consecutive patients with VTE (cancer associated thrombosis was excluded) and 400 healthy control volunteers^40^.All individuals were ≥ 18 years. The diagnosis was confirmed with Doppler ultrasonography, tomography, magnetic resonance, arteriography, phlebography or pulmonary gammagraphy.

Blood samples from the patients were taken at least 6 months after thrombosis to minimize the influence of the acute phase. None of the participants was using oral anticoagulants, heparin, or antiplatelet therapy at the time of blood collection. Controls were selected according to the age and sex distribution of the Spanish population (2001 census). A total of 5 ml of blood was obtained in a Vacutainer tube (BD Vacutainer Becton Dickinson and Company, New Jersey, USA) containing EDTA as anticoagulant. All individuals were genotyped using Infinium Global Screening Array-24 v3.0 kit from Illumina. Written informed consent was obtained for all participants and all procedures were approved by the Institutional Review Board of the Hospital de la Santa Creu i Sant Pau (Barcelona). Antithrombin and Protein C functional levels were were measured in a biochemical analyzer (Metrolab;RAL,Barcelona, Spain) with the use of chromogenic methods from Chromogenix. Functional PS was determined with a kit from Diagnostica Stago. Total PS and free PS were assayed with the use of ELISA methods from Diagnostica Stago (Ansières, France).

The **Trinity Student Study (TSS)** is a cohort study of 2,524 healthy European individuals attending the University of Dublin, Trinity College, with ages between 18 and 28 years, recruited over one academic year in 2003-2004^41,42^. Plasma and DNA was collected from participants as well as demographic, diet and nutrition-based information.  Hemostasis and thrombosis related traits were assayed by collaborators at the University of Michigan.

Antithrombin was measured by a commercial AlphaLISA assay. When available, plasma concentrations were determined by standard curve fitting to plasma standards provided by George King FACT plasma^28^(https://kingbiomed.com/products/fact-factor-assay-control/).

**Supplementary Table S1. Phenotype measurements and sample size in selected cohorts**

| **Study** | **Measure** | **AN** | **Antithrombin** | | | **Protein C** | | | **Protein S Free** | | | **Protein S total** | | |
| --- | --- | --- | --- | --- | --- | --- | --- | --- | --- | --- | --- | --- | --- | --- |
|  |  |  | N | Mean (SD) | Unit | N | Mean (SD) | Unit | N | Mean (SD) | Unit | N | Mean (SD) | Unit |
| **ARIC** | Activity | EA | 9,179 | 110.2(20.9) | % | / | / | / | / | / | / | / | / | / |
|  | Antigen | EA | / | / | / | 9,180 | 3.18(0.6) | Ug/mL | / | / | / | / | / | / |
|  | Activity | AA | 2,688 | 115.3(23.1) | % | / | / | / | / | / | / | / | / | / |
|  | Antigen | AA | / | / | / | 2,688 | 3.14(0.6) | Ug/mL | / | / | / | / | / | / |
| **CHS** | Activity | EA | 681 | 109.4 (14.1) | % | / | / | / | / | / | / | / | / | / |
|  | Antigen | EA | / | / | / | 317/366 | 3.5 (0.7)/4.9(1.9) | ug/mL | 288 | 5.02 (0.94) | Ug/dL | 290 | 22.43 (3.09) | Ug/dL |
| **CHRIS** | Activity | EA | 9,012 | 102.5 (11.5) | % | / | / | / | / | / | / | / | / | / |
| **GABC** | / | / | / | / | / | 919 | 118.7(20.21) | IU/dL | / | / | / | 934 | 83.49(26.5) | IU/mL*100 |
|  | Antigen | EA | 932 | 36.32 (6.5) | IU/mL*100 | / | / | / | / | / | / | / | / | / |
| **GAIT** | Activity | EA | 890 | 112.1(13.5) | IU/mL*100 | 890 | 115.8(26.6) | IU/mL*100 | / | / | / | 911 | 102.7(24) | IU/mL*100 |
|  | Antigen | EA | / | / | / | / | / | / | 922 | 94.7 (23.0) | IU/mL*100 | 922 | 96.5(20) | IU/mL*100 |
| **HVH1** | Activity | EA | 148 | 109(13.5) | % | / | / | / | / | / | / | / | / | / |
|  | Antigen | EA | / | / | / | / | / | / | 107 | 95(21) | IU/mL*100 | 151 | 115(18) | IU/mL*10) |
| **LURIC** | Activity | EA | 2,705 | 97.1 (13.4) | % | 1,041 | 110.5 (22.6) | % | 1049 | 101 (27.4) | % | 1,040 | 118.9 (31.6) | % |
| **MARTHA** | Activity | EA | 897 | 102.9 (11.6) | % | 949 | 111.6 (24.7) | % | 949 | 90.4(21.5) | % | / | / | / |
| **RETROVE cases** | Activity | EA | 399 | 106.9(13.4) | IU/mL*100 | 399 | 127.498 (24.67) | IU/mL*100 | / | / | / | 398 | 109.944 (20.3) | IU/mL*100 |
|  | Antigen | EA | / | / | / | / | / | / | 398 | 100.6 (21.3) | IU/mL*100 | / | / | / |
| **RETROVE controls** | Activity | EA | 400 | 107.81(10.8) | IU/mL*100 | 400 | 120.49 (23.855) | IU/mL*100 | / | / | / | 400 | 108.3(21.3) | IU/mL*100 |
|  | Antigen | EA | / | / | / | / | / | / | 400 | 99.6(21.3) | IU/mL*100 | / | / | / |
| **TSS** | Activity | EA | / | / | / | 2136 | 128.33 (19.55) | IU/dL | / | / | / | 2272 | 84.44 (27.0) | IU/mL |

AN: Ancestry, EA: European; AA: African.

**Supplementary Table S2. Detailed Description of cohort characteristics, and methods for phenotyping, genotyping and imputation**

| **Study** | **Ancestry** | **Phenotype** | **Study design** | **Population** | **Cite(s)** | **Time** | **Genotype measurement** | **Phenotype measurement** | **Imputation** |
| --- | --- | --- | --- | --- | --- | --- | --- | --- | --- |
| **Atherosclerosis Risk in Communities Study (ARIC)** | European/African | AT act, PC act, PC ant | Population-based cohort study | Men and Women aged 45-65 years in the selected US communities | North Carolina, Mississippi, Minnesota, Maryland, US | 1987- | Affymetrix SNP array 6.0 | ELISA | TOPMed |
| **Cardiovascular Health Study (CHS)** | European/African | AT act, PST act, PSF act | Population-based cohort study | Adults aged over 65 years in the selected US communities | North Carolina, California, Maryland, Pennsylvania, US | 1989-2010 | Illumina 370CNV BeadChip/HumanOmnil-Quad_v1 Bead Chip | Coag-A-mate X2 | TOPMed |
| **The Cooperative Health Research in South Tyrol Study (CHRIS)** | European | AT act | Population-based cohort study | Adults aged in middle and Upper Vinschgau/Val Venosta area of South Tyrol, Italy | Middle and upper Vinschgau/Val Venosta area of South Tyrol, Italy | 2011- | Illumina Human OmniExpress Exome Bead Chip. |  | TOPMed |
| **Genes and Blood-Clotting Study (GABC)** | European | AT ant, PC act, PST act | Population-based cohort study | Healthy sibilings aged between 14 and 35 from University of Michigan | Michigan, US | 2006-2009 | Illumina HumanOmni1-Quad_v1-0_B array | AlphaLISA assay | TOPMed |
| **Genetic Analysis of Idiopathic Thrombophilia project (GAIT)** | European | AT act, PC act, PSF ant, PST act. PST ant | Family-based cohort  study | 397 individuals from 21 families selected from a proband with VTE diagnosis and pedigree size | Barcelona, Spain | 1999-2000 | HumanOmniExpressExome-8v1.2 (324 individuals and coverage 964,193 variants) and HumanCoreExome-12v1.1 (610 individuals and coverage 542,585 variants) | CPA/ ELISA | TOPMed |
| **HVH** | European | PSF ant, PST ant | Population-based case control study | Perimenopausal and postmenopausal women aged 30 to 89 in Washington State, US | Washington State | 1995-2002 | GoldenGate custom panel using BeadArray tech | Medical record | TOPMed |
| **Ludwigshafen Risk and Cardiovascular Health study (LURIC)** | European | AT act, PC act, PSF act, PST act | Hospital-based cohort study | Patients diagnosed with coronary angiography | Southwestern Germany | 1997-2000 | Affymetrix SNP array 6.0 |  | TOPMed |
| **MARseille THrombosis Association Project (MARTHA)** | European | AT act, PC act | Hospital-based cohort study | People with documented history VT and free of AT, PS, PC deficiency or other coagulation related status | La Timone hospital (Marseille, France) | 1994-2012 | Illumina Human610-Quad Beadchip/Illumina Human660W-Quad Beadchip | amidolytic assay (STA-Stachrom ATIII kit)/chronometci assay/Asserachrom f-PS assay | TOPMed |
| **Riesgo de Enfermedad TROmboembolica VEnosa (RETROVE)** | European | AT act, PC act, PC ant, PSF ant, PST act | Hospital-based case-control study | VTE patients and controls from the hospital | Barcelona**,** Spain |  | Illumina Infinium Global Screening Array-24 v2.0 |  | TOPMed |
| **Trinity Student Study (TSS)** | European | PC act, PST act | Population-based cohort study | Irish individuals aged 18 to 28 years attending University of Dulbin | Trinity, Ireland | 2003-2004 | he Illumina HumanOmni1-Quad_v1-0_B array |  | TOPMed |
| AT: antithrombin, PC: protein C, PSF: protein S free, PST: protein S total, act: activity, ant: antigen. | | | | | | | | | |

**Supplementary Table S3. Functional annotations for candidate variants, derived from HaploReg v4.1**

| **Phenotype** | **Chr:Pos:A1:A2** | **RSID** | **Gene** | **Functional annotation** |
| --- | --- | --- | --- | --- |
| AT | 1:173914872:A:T | rs2227624 | *SERPINC1* | MV to *SERPINC1*.Active transcription marks in liver. Estrogen receptor binding motif. |
| AT | 2:27375230:T:C | rs4665972 | *SNX17* | IV to *SNX17*; GWAS: Triglycerides |
| AT | 7:7349751:T:C | rs13244268 | *BAZ1B* | IV to *BAZ1B*; Active transcription marks in liver |
| AT | 16:72054562:A:C | rs5471 | *HP/TXNL4B* | IV to *HP*; 5’ UTR to *TXNL4B* |
| AT | 19:49513222:T:G | rs111981233 | *FCGRT* | IV to *FCGRT* |
| PC | 1:1092794968:T:G | rs12740374 | *CELSR2* | 3’ UTR to *CELSR2*; GWAS: LDL cholesterol, phospholipase A2 |
| PC | 2:127418299:A:G | rs1799809 | *PROC* | 0.1 KB 5’ to *PROC* |
| PC | 2:27375230:T:C | rs4665972 | *SNX17* | IV to *SNX17*; GWAS: Triglycerides |
| PC | 7:73625076:C:G | rs35493868 | *MLXIPL* | 2KB 5’ to *MLXIPL* |
| PC | 15:42980693:T:C | rs529330569 | *UBR1* | IV to *UBR1* |
| PC | 20:35179967:T:C | rs11907011 | *PROCR* | IV to *PROCR* |
| PSF | 3:93868695:T:C | rs528128538 | *PROS1* | 1KB 5’ to *PROS1* |
| PSF | 7:444568571:T:C | rs141292869 | *DDX56* | IV to *DDX56* |
| PSF | 9:114321523:A:C | rs150611042 | *ORM1* | 2KB 5’ to *ORM1*; GWAS: Thrombin generation potentials |
| PST | 7:45231101:A:T | rs59569024 | *MYL7* | 380KB 5’ to *MYL7** |
| PST | 9:114321523:A:C | rs150611042 | *ORM1* | 2KB 5’ to *ORM1*; GWAS: Thrombin generation potentials |
| Multi-phenotype | 15:43528519:T:C | rs55707100 | *MAP1A* | MV to *MAP1A* |

AT: antithrombin, PC: protein C, PSF: protein S free, PST: protein S total, Chr: chromosome, Pos: position, A1: effect allele, A2: other allele, IV: intronic variant, MV: missense variant, 3’UTR: 3 prime untranslated regions; 5’UTR: 5 prime untranslated regions.

**Supplementary Table S4. Participants and cohorts count of all specific meta-analyses**

| **Phenotype** | **Cross-ethnic** | | **African** | | **European** | |
| --- | --- | --- | --- | --- | --- | --- |
|  | **Cohorts** | **Participants** | **Cohorts** | **Participants** | **Cohorts** | **Participants** |
| AT/Activity | - | - | 1 | 2,688 | 8 | 24,311 |
| AT/Antigen | - | - | - | - | 1 | 932 |
| AT/All | 9 | 27,931 | 1 | 2,688 | 9 | 25,243 |
| PC/Activity | - | - | - | - | 6 | 6,734 |
| PC/Antigen | - | - | 1 | 2,688 | 2 | 9,863 |
| PC/All | 8 | 19,285 | 1 | 2,688 | 8 | 16,597 |
| PST/Activity | - | - | - | - | 4 | 5,045 |
| PST/Antigen | - | - | - | - | 3 | 1,363 |
| PST/All | - | - | - | - | 7 | 6,408 |
| PSF/Activity | - | - | - | - | 2 | 1,998 |
| PSF/Antigen | - | - | - | - | 4 | 2,115 |
| PSF/All | - | - | - | - | 6 | 4,113 |

AT: Antithrombin, PC: Protein C, PST: Protein S Total, PSF: Protein S Free

**Supplementary Table S5. Number of variants considered for each specific cohort (stratified by ancestry) before and after quality control**

| **Study Name** | **Variants In** | **Variants Out** | **N** | **Lambda** |
| --- | --- | --- | --- | --- |
| Antithrombin Autosome | | | | |
| ARIC EA | 70,219,668 | 17,889,491 | 9,179 | 1.005 |
| CHRIS EA | 23,250,229 | 15,754,873 | 9,012 | 1.017 |
| CHS EA | 14,442,735 | 9,046,889 | 681 | 1.001 |
| GABC EA | 22,307,894 | 9,767,792 | 932 | 1.044 |
| GAIT2 EA | 18,771,000 | 9,564,407 | 890 | 1.021 |
| LURIC EA | 35,830,264 | 11,151,214 | 2,705 | 1.005 |
| MARTHA EA | 13,548,418 | 9,674,035 | 897 | 0.997 |
| RETROVECASES EA | 24,924,998 | 8,307,177 | 399 | 0.994 |
| RETROVECONTROLS EA | 24,963,152 | 8,311,223 | 400 | 0.996 |
| ARIC AA | 67,480,830 | 25,070,269 | 2,688 | 1.008 |
| Antithrombin X chromosome | | | | |
| ARIC XF | 2,634,440 | 524,469 | 4,866 | 0.997 |
| ARIC XM | 2,526,220 | 509,663 | 4,313 | 1.008 |
| CHS XF | 520,226 | 267,751 | 514 | 0.994 |
| CHS XM | 520,226 | 205,063 | 167 | 0.975 |
| GAIT2 XF | 568,088 | 238,551 | 449 | 0.988 |
| GAIT2 XM | 555,598 | 237,844 | 441 | 1.003 |
| MARTHA XF | 387,325 | 257,744 | 643 | 0.988 |
| MARTHA XM | 286,764 | 210,053 | 254 | 0.959 |
| retroveCASES XF | 829,108 | 215,542 | 203 | 0.951 |
| retroveCASES XM | 742,620 | 214,540 | 196 | 0.955 |
| retroveCONTROLS XF | 828,855 | 216,582 | 206 | 0.960 |
| retroveCONTROLS XM | 739,571 | 211,793 | 194 | 1.034 |
| Protein C Autosome | | | | |
| ARIC EA | 70,219,809 | 17,890,753 | 9,180 | 1.029 |
| CHS 1 EA | 14,442,735 | 8,183,875 | 366 | 1.008 |
| CHS 2 EA | 14,442,735 | 7,952,747 | 317 | 1.011 |
| GABC EA | 34,112,072 | 9,737,717 | 919 | 1.035 |
| GAIT2 EA | 19,894,686 | 9,813,030 | 890 | 1.028 |
| LURIC EA | 35,830,264 | 9,059,673 | 1,041 | 0.997 |
| MARTHA EA | 13,728,590 | 9,773,535 | 949 | 0.999 |
| retroveCASES EA | 26,496,726 | 8,535,945 | 399 | 1.015 |
| retroveCONTROLS EA | 26,531,578 | 8,539,692 | 400 | 1.010 |
| TSS EA | 28,227,516 | 11,107,799 | 2,136 | 1.007 |
| ARIC AA | 67,480,830 | 25,070,269 | 2,688 | 1.014 |
| Protein C X Chromosome | | | | |
| ARIC XF | 2,634,454 | 524,485 | 4,866 | 1.038 |
| ARIC XM | 2,526,284 | 509,725 | 4,314 | 1.010 |
| CHS 1 XM | 520,226 | 245,952 | 366 | 1.002 |
| CHS 2 XF | 520,226 | 199,852 | 150 | 0.982 |
| CHS 2 XM | 520,226 | 205,063 | 167 | 0.980 |
| GABC XF | 1,116,585 | 291,773 | 562 | 1.179 |
| GABC XM | 1,059,867 | 247,835 | 357 | 0.767 |
| GAIT2 XF | 568,088 | 238,551 | 449 | 0.963 |
| GAIT2 XM | 555,598 | 237,844 | 441 | 1.141 |
| MARTHA XF | 390,592 | 260,079 | 677 | 0.988 |
| MARTHA XM | 291,911 | 213,324 | 272 | 1.050 |
| retroveCASES XF | 829,108 | 215,542 | 203 | 0.966 |
| retroveCASES XM | 742,620 | 214,540 | 196 | 0.950 |
| retroveCONTROLS XF | 828,855 | 216,582 | 206 | 0.886 |
| retroveCONTROLS XM | 739,571 | 211,793 | 194 | 0.992 |
| Protein S free Autosome | | | | |
| CHS | 14,442,735 | 7,809,143 | 288 | 1.022 |
| GAIT2 | 19,910,472 | 9,896,490 | 922 | 1.036 |
| HVH1 | 15,455,952 | 1,902,147 | 107 | 1.013 |
| LURIC | 35,830,264 | 9,074,110 | 1,049 | 1.016 |
| MARTHA | 13,759,440 | 9,780,346 | 949 | 0.993 |
| retroveCASES | 26,492,308 | 8,531,798 | 398 | 1.005 |
| retroveCONTROLS | 26,531,578 | 8,539,692 | 400 | 1.012 |
| Protein S free X Chromosome | | | | |
| CHS XF | 520,226 | 193,777 | 132 | 0.921 |
| CHS XM | 520,226 | 202,047 | 156 | 0.987 |
| GAIT2 XF | 568,380 | 240,782 | 465 | 0.954 |
| GAIT2 XM | 556,577 | 239,807 | 457 | 1.193 |
| MARTHA XF | 392,378 | 260,309 | 673 | 1.024 |
| MARTHA XM | 292,793 | 213,940 | 276 | 1.013 |
| retroveCASES XF | 828,693 | 215,349 | 202 | 1.006 |
| retroveCASES XM | 742,620 | 214,540 | 196 | 1.038 |
| retroveCONTROLS XF | 828,855 | 216,582 | 206 | 1.039 |
| retroveCONTROLS XM | 739,571 | 211,793 | 194 | 1.087 |
| Protein S total | | | | |
| CHS | 14,442,735 | 7,818,959 | 290 | 0.997 |
| GABC | 34,112,072 | 9,771,502 | 934 | 1.043 |
| GAIT2 activity | 19,908,400 | 9,870,882 | 911 | 1.040 |
| GAIT2 antigen | 19,910,472 | 9,896,490 | 922 | 1.032 |
| HVH1 | 15,455,952 | 1,902,147 | 151 | 1.002 |
| LURIC | 35,830,264 | 9,059,870 | 1,041 | 1.013 |
| retroveCASES | 26,492,308 | 8,531,798 | 398 | 1.007 |
| retroveCONTROLS | 26,531,578 | 8,539,692 | 400 | 1.011 |
| TSS | 28,227,516 | 1,121,0226 | 2,272 | 1.009 |
| Protein S total X Chromosome | | | | |
| CHS XF | 520,226 | 193,777 | 132 | 1.043 |
| CHS XM | 520,226 | 202,657 | 158 | 0.984 |
| GABC XF | 1,117,388 | 292,736 | 571 | 0.885 |
| GABC XM | 1,062,748 | 251,085 | 363 | 0.890 |
| GAIT2 activity XF | 568,274 | 239,811 | 458 | 1.077 |
| GAIT2 activity XM | 556,390 | 239,427 | 453 | 1.198 |
| GAIT2 antigen XF | 568,380 | 240,782 | 465 | 1.012 |
| GAIT2 antigen XM | 556,577 | 239,807 | 457 | 1.062 |
| LURIC XF | 35,830,264 | 9,059,870 | 1,041 | 1.013 |
| retroveCASES XF | 828,693 | 215,349 | 202 | 1.050 |
| retroveCASES XM | 742,620 | 214,540 | 196 | 0.913 |
| retroveCONTROLS XF | 828,855 | 216,582 | 206 | 0.921 |
| retroveCONTROLS XM | 739,571 | 211,793 | 194 | 1.059 |

XF: X chromosome in females, XM: X chromosome in males, Lambda: genomic control coefficients. Analyses were stratified and by sex for X-chromosome

**Supplementary Table S6. Additional independent variants from conditional analyses**

| **Candidate gene in the loci** | **Independent Gene/Variant location** | **RSID** | **Beta** | **SE** | **Joint Effect** | **Joint SE** | **Original P-value** | **Joint P-value** | **LD R^2^** | **Phenotype/Ancestry** |
| --- | --- | --- | --- | --- | --- | --- | --- | --- | --- | --- |
| *SERPINC1* | *RABGAP1L*/IV | rs182221508 | 13.40 | 2.14 | 13.51 | 2.14 | 3.91E-10 | 2.97E-10 | 0.0046 | AT EA |
| *PROC* | *PROC*/IV | rs73492719 | -0.25 | 0.05 | -0.33 | 0.05 | 1.23E-06 | 1.29E-10 | -0.0086 | PC EA |
| *PROCR* | *PROCR*/IV, *MMP24-AS1-EDEM2*/IV | rs6060300 | 0.23 | 0.01 | 0.16 | 0.01 | 1.14E-65 | 5.92E-30 | 0.0000 | PC EA |

IV: Intronic Variant, AT: antithrombin, PC: protein C, EA: European, SE: standard error, LD R^2^: linkage disequilibrium r^2^.

**Supplementary Table S7. Ancestry specific results of antithrombin and protein C meta-analyses**

| **Ancestry** | **Chr:Pos:A1:A2** | **RSID** | **Closest Genes** | **N** | **EAF** | **Beta**  **(SE)** | **P-value** |
| --- | --- | --- | --- | --- | --- | --- | --- |
| Antithrombin | | | | | | | |
| AA | 16:72054562:A:C | rs5471 | *HP\|TXNL4B* | 2688 | 0.866 | 9.77 (0.95) | 1.37E-24 |
| EA | 1:173914872:A:T | rs2227624 | *SERPINC1* | 24414 | 0.994 | 8.11 (0.20) | 3.33E-19 |
| EA | 2:27375230:T:C | rs4665972 | *SNX17* | 25242 | 0.447 | 1.00 (0.12) | 7.87E-16 |
| Protein C | | | | | | | |
| AA | 2:127423170:T:C | rs200045749 | *PROC\|MIR4783* | 2688 | 0.014 | -1.41 (0.12) | 4.83E-34 |
| AA | 20:35176751:A:G | rs867186 | *PROCR\|MMP24-AS1-EDEM2* | 2688 | 0.909 | -0.73 (0.05) | 4.96E-54 |
| EA | 1:109279544:A:G | rs599839 | *PSRC1* | 15556 | 0.773 | 0.09 (0.01) | 4.30E-11 |
| EA | 2:127418299:A:G | rs1799809 | *PROC* | 16597 | 0.569 | 0.21 (0.01) | 5.10E-83 |
| EA | 2:27375230:T:C | rs4665972 | *SNX17* | 16597 | 0.420 | 0.11 (0.01) | 1.92E-23 |
| EA | 7:73562919:T:C | rs34594435 | *BAZ1B \| BCL7B \| TBL2* | 16597 | 0.187 | -0.09 (0.01) | 2.16E-10 |
| EA | 20:35179967:T:C | rs11907011 | *PROCR\|MMP24-AS1-EDEM2* | 16597 | 0.094 | 0.75 (0.02) | 5.07E-363 |
| EA | 20:34176377:T:C | rs17332951 | *RPS2P1\|ASIP* | 16597 | 0.952 | -0.51 (0.03) | 1.05E-88 |
| EA | 20:36270102:A:G | rs117473488 | *AAR2\|DLGAP4* | 16597 | 0.018 | 0.50 (0.04) | 3.58E-33 |

AA: African ancestry, EA: European ancestry, Chr: chromosome, Pos: position, A1: effect allele, A2: other allele, SE: standard error, EAF: effect allele frequency.

**Supplementary Table S8. Summary statistics results for the selected lead variants in DeCODE genetics dataset**

| **Chr:Pos:A1:A2** | **RSID** | **Beta** | **P-value** | **SE** | **N** | **EAF** |
| --- | --- | --- | --- | --- | --- | --- |
| Antithrombin | | | | | | |
| 1:173914872:T:A | rs2227624 | -0.48 | 3.10E-21 | 0.05 | 35,351 | 0.005 |
| 2:27375230:C:T | rs4665972 | -0.04 | 3.16E-06 | 0.01 | 35,374 | 0.339 |
| 7:73497513:C:T | rs13244268 | -0.04 | 0.003 | 0.01 | 35,354 | 0.111 |
| 19:49513222:G:T | rs111981233 | 0.05 | 0.002 | 0.02 | 35,375 | 0.060 |
| Protein C | | | | | | |
| 1:109279544:A:G | rs599839 | 0.08 | 1.71E-17 | 0.01 | 35,344 | 0.208 |
| 2:127418299:A:G | rs1799809 | 0.14 | 4.90E-69 | 0.01 | 35,366 | 0.419 |
| 2:27375230:C:T | rs4665972 | -0.06 | 6.93E-14 | 0.01 | 35,374 | 0.339 |
| 7:73625076:G:C | rs35493868 | -0.03 | 0.004 | 0.01 | 35,354 | 0.199 |
| 20:35179967:T:C | rs11907011 | 0.55 | <5E-307 | 0.01 | 35,367 | 0.093 |
| Protein S | | | | | | |
| 3:93868695:T:C | rs528128538 | -0.93 | 1.44E-10 | 0.15 | 35,327 | 0.001 |
| 9:114321523:A:C | rs150611042 | -0.30 | 7.53E-102 | 0.01 | 35,361 | 0.099 |

Chr: chromosome, Pos: position, A1: effect allele, A2: other allele, SE: standard error; EAF: effect allele frequency. Analysis population was based on Iceland (European) population.

**Supplementary Table S9. Significant results from TWAS S-MultiXcan analyses**

| **Region** | **Gene** | **Best predicted tissue** | **P-value** |
| --- | --- | --- | --- |
| Antithrombin | | | |
| chr1:173924653-173858808 | *DARS2* | Artery Coronary | 3.36E-07 |
| chr1:173868082-173903549 | *ZBTB37* | Artery Aorta | 2.28E-10 |
| chr1:173903800-173917327 | *SERPINC1* | Whole Blood | 8.78E-12 |
| chr1:174159410-174995308 | *RABGAP1L* | Whole Blood | 1.13E-09 |
| chr1:175067833-175148075 | *TNN* | Liver | 3.40E-09 |
| chrchr2:27496839-27523684 | *GCKR* | Liver | 1.12E-06 |
| chrchr7:73536356-73557960 | *BCL7B* | Whole Blood | 2.08E-06 |
| chrchr7:73593194-73624543 | *MLXIPL* | Artery Coronary | 1.87E-06 |
| chr19:49506816-49526428 | *FCGRT* | Artery Coronary | 1.12E-06 |
| Protein C | | | |
| chr1:109279556-109283186 | *PSRC1* | Liver | 3.82E-09 |
| chr1:109399042-109426448 | *PSMA5* | Liver | 3.17E-09 |
| chr2:27381195-27409591 | *PPM1G* | Artery Aorta | 6.28E-07 |
| chr2:27427790-27442259 | *NRBP1* | Artery Aorta | 2.19E-08 |
| chr2:27442366-27446481 | *KRTCAP3* | Artery Tibial | 1.34E-07 |
| chr2:27496839-27523684 | *GCKR* | Liver | 3.26E-10 |
| chr2:27537386-27582720 | *C2orf16* | Liver | 3.66E-07 |
| chr2:27663471-27694976 | *SLC4A1AP* | Artery Aorta | 2.01E-07 |
| chr2:127183832-127220313 | *CYP27C1* | Artery Tibial | 1.46E-09 |
| chr2:127257290-127294166 | *ERCC3* | Artery Tibial | 1.67E-49 |
| chr2:127298668-127388465 | *MAP3K2* | Artery Coronary | 2.53E-68 |
| chr2:127388178-127406228 | *AC068282.3* | Whole Blood | 2.46E-07 |
| chr2:127418427-127429242 | *PROC* | Liver | 6.78E-78 |
| chr2:127436207-127526886 | *IWS1* | Artery Aorta | 2.02E-24 |
| chr2:127638381-127681786 | *LIMS2* | Whole Blood | 6.74E-14 |
| chr7:73593194-73624543 | *MLXIPL* | Artery Coronary | 2.35E-08 |
| chr15:43323649-43330582 | *LCMT2* | Artery Aorta | 1.62E-06 |
| chr15:43628503-43668118 | *CATSPER2* | Whole Blood | 1.27E-06 |
| chr15:44288719-44415758 | *GOLM2* | Whole Blood | 3.19E-07 |
| chr20:33490075-33650036 | *CBFA2T2* | Artery Coronary | 1.31E-09 |
| chr20:33655701-33656423 | *RP1-63M2.7* | Liver | 4.32E-08 |
| chr20:34088309-34112243 | *EIF2S2* | Whole Blood | 1.44E-53 |
| chr20:34280268-34311802 | *AHCY* | Liver | 1.20E-67 |
| chr20:34363241-34540748 | *ITCH* | Artery Aorta | 2.47E-10 |
| chr20:34560542-34698790 | *PIGU* | Artery Tibial | 7.33E-100 |
| chr20:34704339-34713439 | *TP53INP2* | Artery Coronary | 2.17E-18 |
| chr20:34844720-34872856 | *GGT7* | Artery Tibial | 2.36E-49 |
| chr20:34872146-34927962 | *ACSS2* | Liver | 3.79E-08 |
| chr20:34928432-34956027 | *GSS* | Artery Tibial | 1.50E-241 |
| chr20:34955810-35002437 | *MYH7B* | Liver | 9.83E-38 |
| chr20:35002404-35092807 | *TRPC4AP* | Whole Blood | 3.82E-256 |
| chr20:35115364-35147336 | *EDEM2* | Artery Tibial | 2.25E-98 |
| chr20:35172072-35216240 | *PROCR* | Liver | 2.71E-239 |
| chr20:35201745-35278131 | *MMP24-AS1* | Artery Aorta | 9.29E-96 |
| chr20:35226690-35276998 | *MMP24* | Artery Coronary | 2.32E-43 |
| chr20:35278907-35284985 | *EIF6* | Liver | 8.42E-173 |
| chr20:35542038-35557634 | *ERGIC3* | Artery Tibial | 3.08E-25 |
| chr20:35615829-35621094 | *SPAG4* | Liver | 8.39E-60 |
| chr20:35648925-35664956 | *RBM12* | Liver | 4.60E-61 |
| chr20:35668052-35699355 | *NFS1* | Artery Tibial | 3.68E-67 |
| chr20:35701347-35742312 | *RBM39* | Artery Aorta | 1.63E-22 |
| chr20:35953617-35959472 | *SCAND1* | Artery Coronary | 9.19E-71 |
| chr20:35954564-36030700 | *CNBD2* | Artery Aorta | 1.12E-25 |
| chr20:36045618-36051018 | *NORAD* | Liver | 7.27E-10 |
| chr20:36651766-36746090 | *NDRG3* | Artery Coronary | 1.29E-14 |
| chr20:33983052-33993124 | *RALY-AS1* | Artery Tibial | 1.08E-50 |
| chr20:33666498-33668525 | *ACTL10* | Whole Blood | 2.11E-07 |
| Protein S | | | |
| chr3:93873051-93980003 | *PROS1* | Artery Aorta | 5.86E-07 |
| chr3:93980139-94055678 | *ARL13B* | Artery Aorta | 8.96E-07 |
| chr7:44138864-44141332 | *MYL7* | Liver | 1.98E-06 |
| chr9:114329869-114332521 | *ORM2* | Whole Blood | 8.97E-08 |

**Supplementary Table 10. Prioritized genes in fine-mapping analyses.**

| **Region** | **Prioritized genes** | | | | |
| --- | --- | --- | --- | --- | --- |
|  | **Artery**  **Aorta** | **Artery Coronary** | **Artery**  **Tibial** | **Liver** | **Whole**  **Blood** |
| Antithrombin | | | | | |
| chr1:173099832-175089390 | *SERPINC1* | *SERPINC1* | *SERPINC1* | NA | *SERPINC1* |
| chr2:26896379-28598746 | NA | NA | NA | *TNN/NRBP1* | NA |
| Protein C | | | | | |
| chr1:108468105-1:110303740 | NA | NA | NA | *PSRC1* | *PSRC1* |
| chr2:26896379-2:28598746 | NRPB1 | NA | NA | *NRBP1* | *NRBP1* |
| chr2:125841643-127372950 | *ERCC3* | *ERCC3* | *ERCC3* | *ERCC3* | NA |
| chr2:127373815-128033703 | *IWS1* | *MAP3K2* | *AC068282.3* | *MAP3K2* | *MAP3K2/PROC* |
| chr7:73335940-76455022 | *MLXIPL* | NA | NA | NA | *TBL2* |
| chr15:42776816-44197582 | *LCMT2* | NA | NA | *LCMT2* | NA |
| chr20:32813520-34959830 | *AHCY* | *AHCY* | *AHCY* | *GSS* | *EIF2S2* |
| chr20:34961516-36909151 | *MMP24-AS1/TRPC4AP/PROCR* | *MMP24-AS1/MYH7B* | *TRPC4AP*  *EDEM2* | *MMP24-AS1/EDEM2/EIF6* | *MMP24-AS1*  */TRPC4Ap/EDEM2* |

Significance was based on posterior inclusion probability calculated with the FOCUS package.

**Supplementary Table S11. Colocalization results in candidate novel loci**

| **Phenotype** | **Chr:Pos:A1:A2** | **Tissue** | **Gene ID** | **Gene Name** | **PP.H3** | **PP.H4** | **CPC** |
| --- | --- | --- | --- | --- | --- | --- | --- |
| AT | 2:27375230:C:T | Artery Tibial | ENSG00000234072 | *GTF3C2-AS2* | 0.16 | 0.73 | 0.82 |
| AT | 2:27375230:C:T | Artery Tibial | ENSG00000115234 | *SNX17* | 1.00 | 6.09E-04 | 6.09E-04 |
| AT | 2:27375230:C:T | Artery Tibial | ENSG00000115216 | *NRBP1* | 0.99 | 0.01 | 0.01 |
| AT | 2:27375230:C:T | Whole Blood | ENSG00000138085 | *ATRAID* | 1.00 | 2.05E-11 | 2.05E-11 |
| AT | 2:27375230:C:T | Whole Blood | ENSG00000115216 | *NRBP1* | 1.00 | 6.30E-06 | 6.30E-06 |
| AT | 2:27375230:C:T | Whole Blood | ENSG00000157992 | *KRTCAP3* | 1.00 | 4.91E-05 | 4.91E-05 |
| AT | 16:72054562:A:C | Artery Tibial | ENSG00000257017 | *HP* | 0.83 | 0.17 | 0.17 |
| AT | 16:72054562:A:C | Artery Tibial | ENSG00000261701 | *HPR* | 1.00 | 6.94E-18 | 6.94E-18 |
| AT | 16:72054562:A:C | Liver | ENSG00000257017 | *HP* | 0.01 | 0.97 | 0.98 |
| AT | 16:72054562:A:C | Whole Blood | ENSG00000257017 | *HP* | 0.11 | 0.89 | 0.89 |
| AT | 16:72054562:A:C | Whole Blood | ENSG00000261701 | *HPR* | 0.72 | 0.28 | 0.28 |
| PSF | 9:114321523:A:C | Liver | ENSG00000228278 | *ORM2* | 0.01 | 0.98 | 0.99 |
| PSF | 9:114321523:A:C | Whole Blood | ENSG00000229314 | *ORM1* | 1.00 | 9.77E-14 | 9.77E-14 |
| PSF | 9:114321523:A:C | Whole Blood | ENSG00000228278 | *ORM2* | 1.00 | 1.25E-05 | 1.25E-05 |
| PST | 9:114321523:A:C | Liver | ENSG00000228278 | *ORM2* | 0.02 | 0.96 | 0.98 |
| PST | 9:114321523:A:C | Whole Blood | ENSG00000229314 | *ORM1* | 1.00 | 1.31E-10 | 1.31E-10 |
| PST | 9:114321523:A:C | Whole Blood | ENSG00000228278 | *ORM2* | 1.00 | 5.90E-06 | 5.90E-06 |

AT: antithrombin, PSF: protein S free, PST: protein S total, Chr: chromosome, Pos: position, A1: effect allele, A2: other allele, PP.H3: posterior probability under hypothesis 3, PP.H4: posterior probability under hypothesis 4, CPC: conditional probability of colocalization.

**Supplementary Table S12. Selected genetic instruments for Mendelian randomization analyses**

| **Outcome** | **RSID** | **Chr:Pos:A1:A2** | **Beta, Exposure** | **SE,**  **Exposure** | **P-value,**  **Exposure** | **EAF,**  **Exposure** | **Beta,**  **Outcome** | **SE,**  **Outcome** | **P-value,**  **Outcome** | **EAF,**  **Outcome** |
| --- | --- | --- | --- | --- | --- | --- | --- | --- | --- | --- |
| Antithrombin | | | | | | | | | | |
| VTE | rs142967416 | 1:175159627:T:C | -7.23 | 0.86 | 3.84E-17 | 0.007 | 0.26 | 0.09 | 0.004 | 0.004 |
|  | rs5471 | 16:72054562:A:C | 9.82 | 0.92 | 1.72E-26 | 0.874 | -0.08 | 0.05 | 0.082 | 0.897 |
|  | rs4665972 | 2:27375230:T:C | 0.98 | 0.12 | 6.63E-16 | 0.443 | -0.01 | 0.01 | 0.414 | 0.403 |
|  | rs13244268 | 7:73497513:T:C | 1.13 | 0.19 | 3.92E-09 | 0.891 | -0.02 | 0.01 | 0.242 | 0.883 |
| PAD | rs142967416 | 1:175159627:T:C | -7.23 | 0.86 | 3.84E-17 | 0.007 | 0.21 | 0.11 | 0.050 | 0.004 |
|  | rs5471 | 16:72054562:A:C | 9.82 | 0.92 | 1.72E-26 | 0.874 | -0.06 | 0.04 | 0.128 | 0.902 |
|  | rs4665972 | 2:27375230:T:C | 0.98 | 0.12 | 6.63E-16 | 0.443 | 0.01 | 0.01 | 0.372 | 0.418 |
|  | rs13244268 | 7:73497513:T:C | 1.13 | 0.19 | 3.92E-09 | 0.891 | -0.01 | 0.02 | 0.554 | 0.889 |
| CAD | rs2227590 | 1:173916899:A:G | 1.28 | 0.18 | 2.65E-13 | 0.149 | 0.01 | 0.01 | 0.498 | 0.145 |
|  | rs4665972 | 2:27375230:T:C | 0.98 | 0.12 | 6.63E-16 | 0.443 | -0.01 | 0.01 | 0.481 | 0.401 |
|  | rs13244268 | 7:73497513:T:C | 1.13 | 0.19 | 3.92E-09 | 0.891 | -0.01 | 0.02 | 0.737 | 0.885 |
| IS | rs2227590 | 1:173916899:A:G | 1.28 | 0.18 | 2.65E-13 | 0.149 | -0.03 | 0.01 | 0.027 | 0.188 |
|  | rs4665972 | 2:27375230:T:C | 0.98 | 0.12 | 6.63E-16 | 0.443 | 0.01 | 0.01 | 0.312 | 0.419 |
|  | rs13244268 | 7:73497513:T:C | 1.13 | 0.19 | 3.92E-09 | 0.891 | 0.02 | 0.01 | 0.259 | 0.888 |
| Protein C | | | | | | | | | | |
| VTE | rs1799809 | 2:127418299:A:G | 0.21 | 0.01 | 9.13E-90 | 0.540 | -0.05 | 0.01 | 1.86E-06 | 0.558 |
|  | rs4665972 | 2:27375230:T:C | 0.11 | 0.01 | 7.09E-24 | 0.406 | -0.01 | 0.01 | 0.414 | 0.403 |
|  | rs35493868 | 7:73625076:C:G | 0.08 | 0.01 | 5.87E-10 | 0.810 | -0.01 | 0.01 | 0.223 | 0.807 |
| PAD | rs1799809 | 2:127418299:A:G | 0.21 | 0.01 | 9.13E-90 | 0.540 | -0.02 | 0.01 | 0.010 | 0.539 |
|  | rs4665972 | 2:27375230:T:C | 0.11 | 0.01 | 7.09E-24 | 0.406 | 0.01 | 0.01 | 0.372 | 0.418 |
|  | rs35493868 | 7:73625076:C:G | 0.08 | 0.01 | 5.87E-10 | 0.810 | -0.01 | 0.01 | 0.594 | 0.818 |
| CAD | rs1799809 | 2:127418299:A:G | 0.21 | 0.01 | 9.13E-90 | 0.540 | -0.02 | 0.01 | 0.020 | 0.562 |
|  | rs4665972 | 2:27375230:T:C | 0.11 | 0.01 | 7.09E-24 | 0.406 | -0.01 | 0.01 | 0.481 | 0.401 |
|  | rs35493868 | 7:73625076:C:G | 0.08 | 0.01 | 5.87E-10 | 0.810 | 0.01 | 0.01 | 0.512 | 0.823 |
| IS | rs1799809 | 2:127418299:A:G | 0.21 | 0.01 | 9.13E-90 | 0.540 | 0.003 | 0.01 | 0.731 | 0.583 |
|  | rs4665972 | 2:27375230:T:C | 0.11 | 0.01 | 7.09E-24 | 0.406 | 0.01 | 0.01 | 0.312 | 0.419 |
|  | rs35493868 | 7:73625076:C:G | 0.08 | 0.01 | 5.87E-10 | 0.810 | 0.01 | 0.01 | 0.328 | 0.822 |

VTE: venous thrombosis; PAD: Peripheral Artery disease; CAD: Coronary artery disease; IS: Ischemic Stroke, Chr: chromosome, Pos: position, A1: effect allele, A2: other allele, SE: standard error, EAF: effect allele frequency.

**Supplementary Table S13. Significant results from MR analyses: AT with VTE and PC with VTE and CAD**

| **Method** | **Number of variants** | **Beta** | **SE** | **P-value** |
| --- | --- | --- | --- | --- |
| Antithrombin to VTE | | | | |
| MR Egger | 4 | -0.19 | 0.12 | 0.251 |
| Weighted median | 4 | -0.13 | 0.07 | 0.056 |
| Inverse variance weighted | 4 | -0.18 | 0.07 | 0.015 |
| Weighted mode | 4 | -0.13 | 0.07 | 0.171 |
| Protein C to VTE | | | | |
| MR Egger | 3 | -0.32 | 0.12 | 0.224 |
| Weighted median | 3 | -0.20 | 0.04 | 9.60E-06 |
| Inverse variance weighted | 3 | -0.18 | 0.05 | 0.000 |
| Weighted mode | 3 | -0.22 | 0.05 | 0.040 |
| Protein C to CAD | | | | |
| MR Egger | 3 | -0.22 | 0.12 | 0.312 |
| Weighted median | 3 | -0.10 | 0.04 | 0.017 |
| Inverse variance weighted | 3 | -0.09 | 0.04 | 0.031 |
| Weighted mode | 3 | -0.10 | 0.05 | 0.149 |

SE: standard error. Significance was based on inverse variance weighted method, with P-value < 0.05.

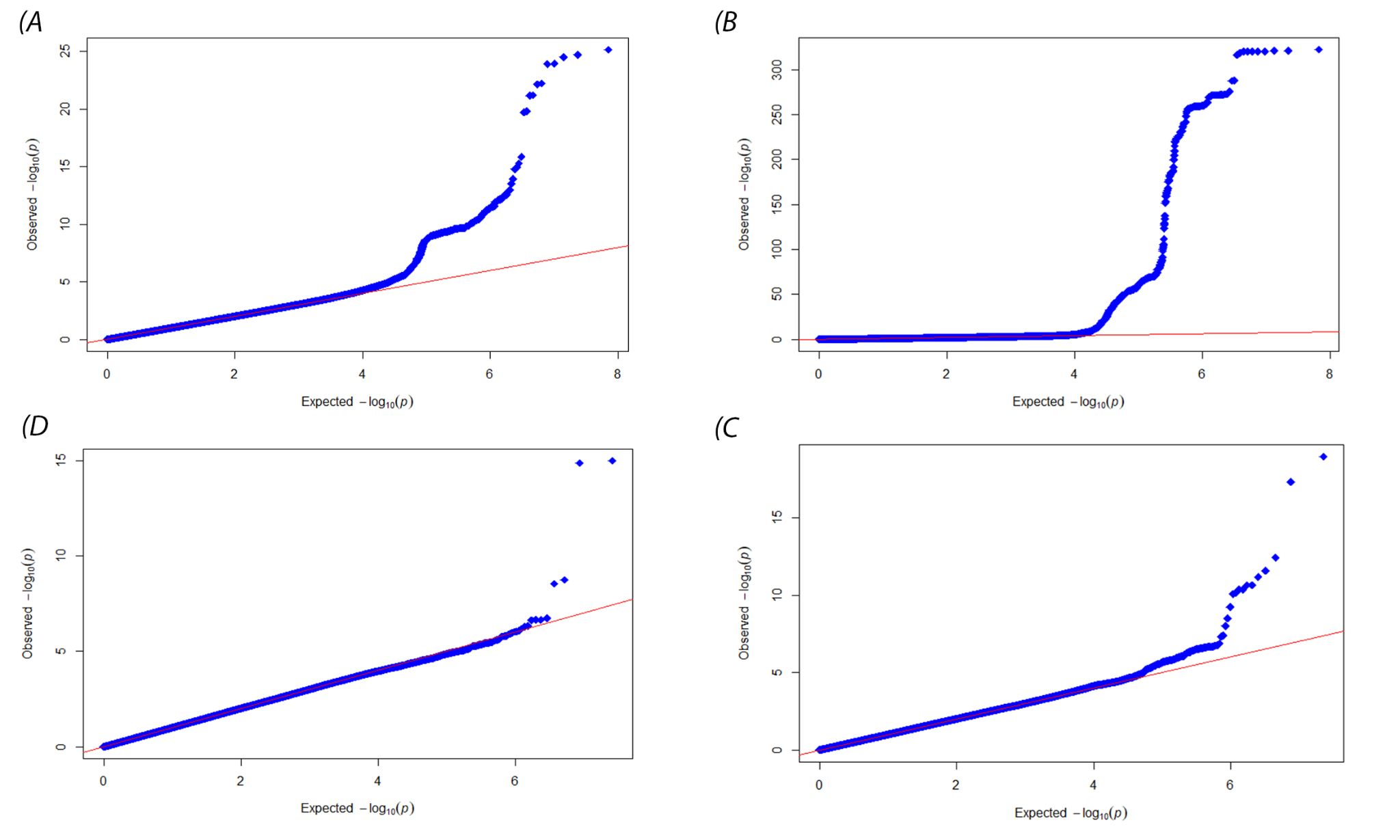

**Supplementary Figure S1. Q-Q Plots of meta-analyses (A: antithrombin cross-ethnic, B: protein C cross ethnic, C: protein S free, D: protein S total).**

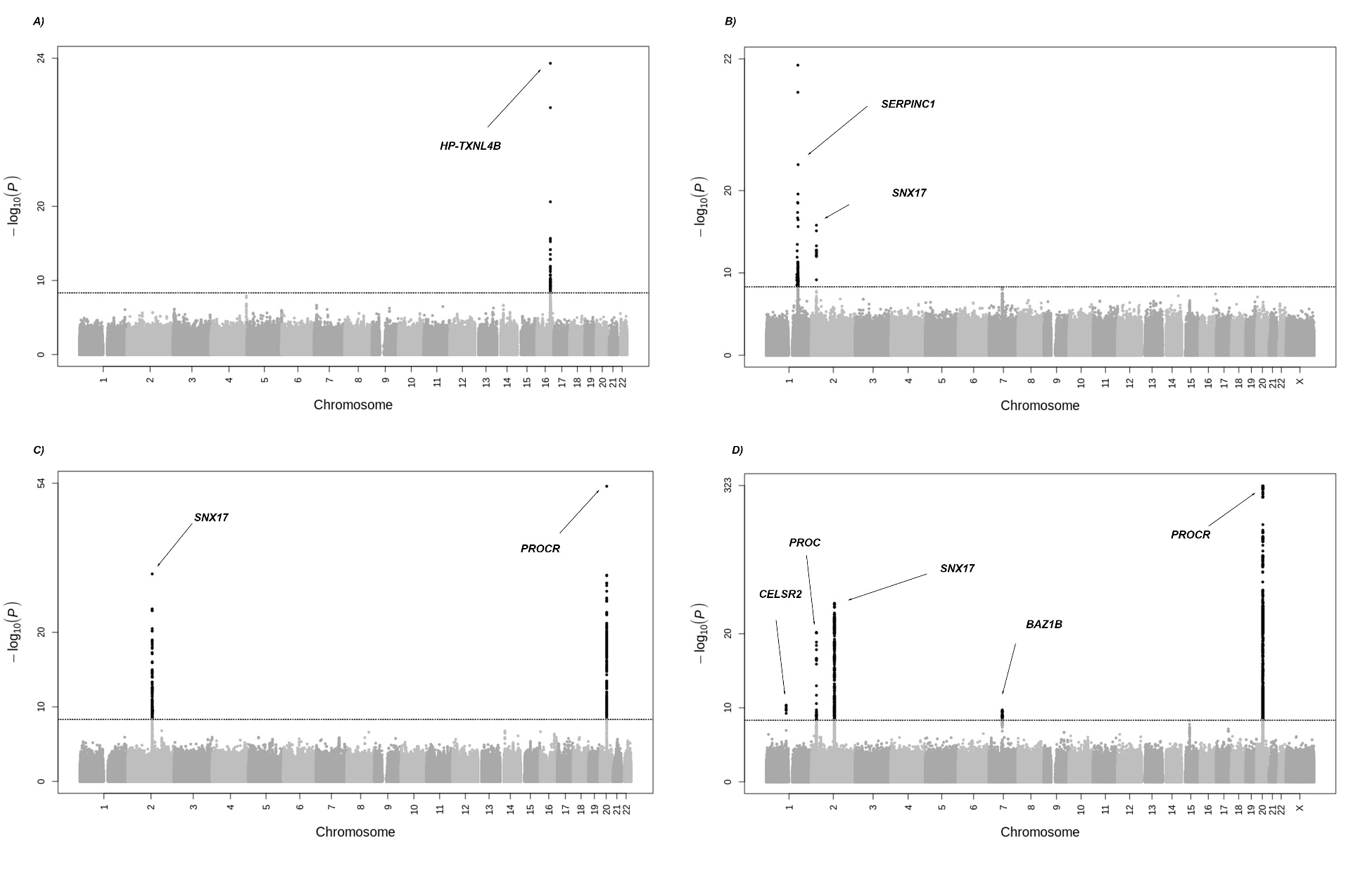

**Supplementary Figure S2. Manhattan plots of population specific meta-analysis of antithrombin and protein C (A: antithrombin European, B: antithrombin African, C: protein C European, D: protein C African)**

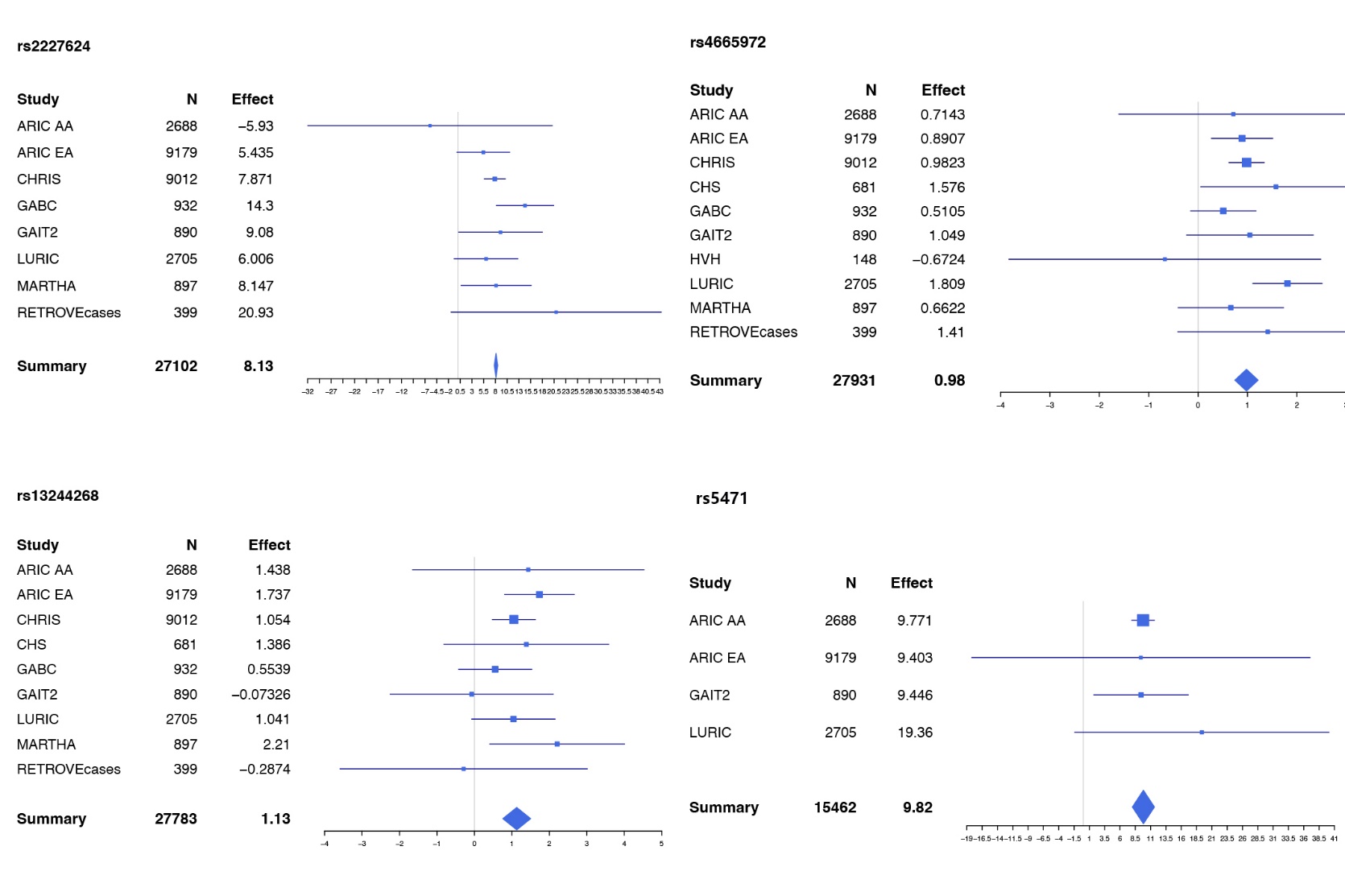

**Supplementary Figure S3. Forest plots of lead variants in antithrombin cross-population meta-analysis**

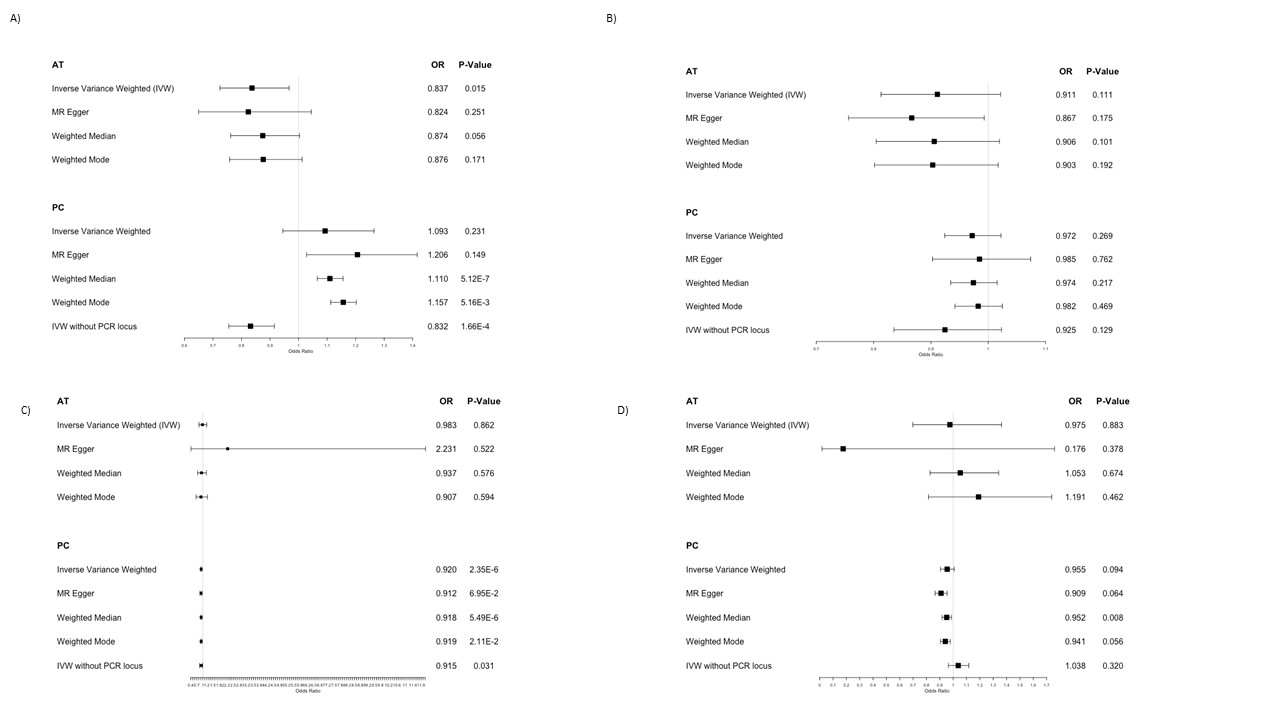

**Supplementary Figure S4. Forest plots of MR analysis results on selected outcomes (A: Venous thrombosis; B: Peripheral Artery disease; C: Coronary artery disease; D: Ischemic Stroke)**

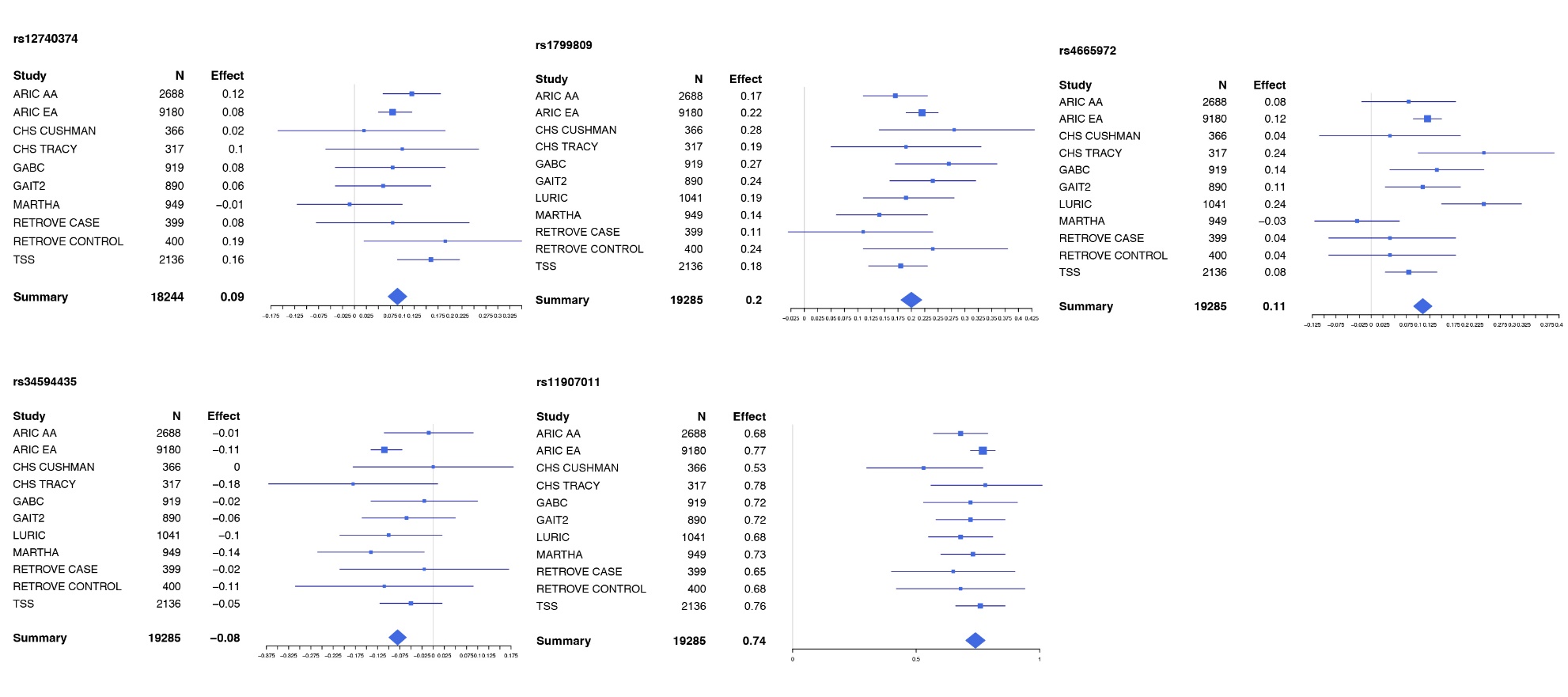

**Supplementary Figure S5. Forest plots of significant loci in protein C cross-ancestry meta-analysis**

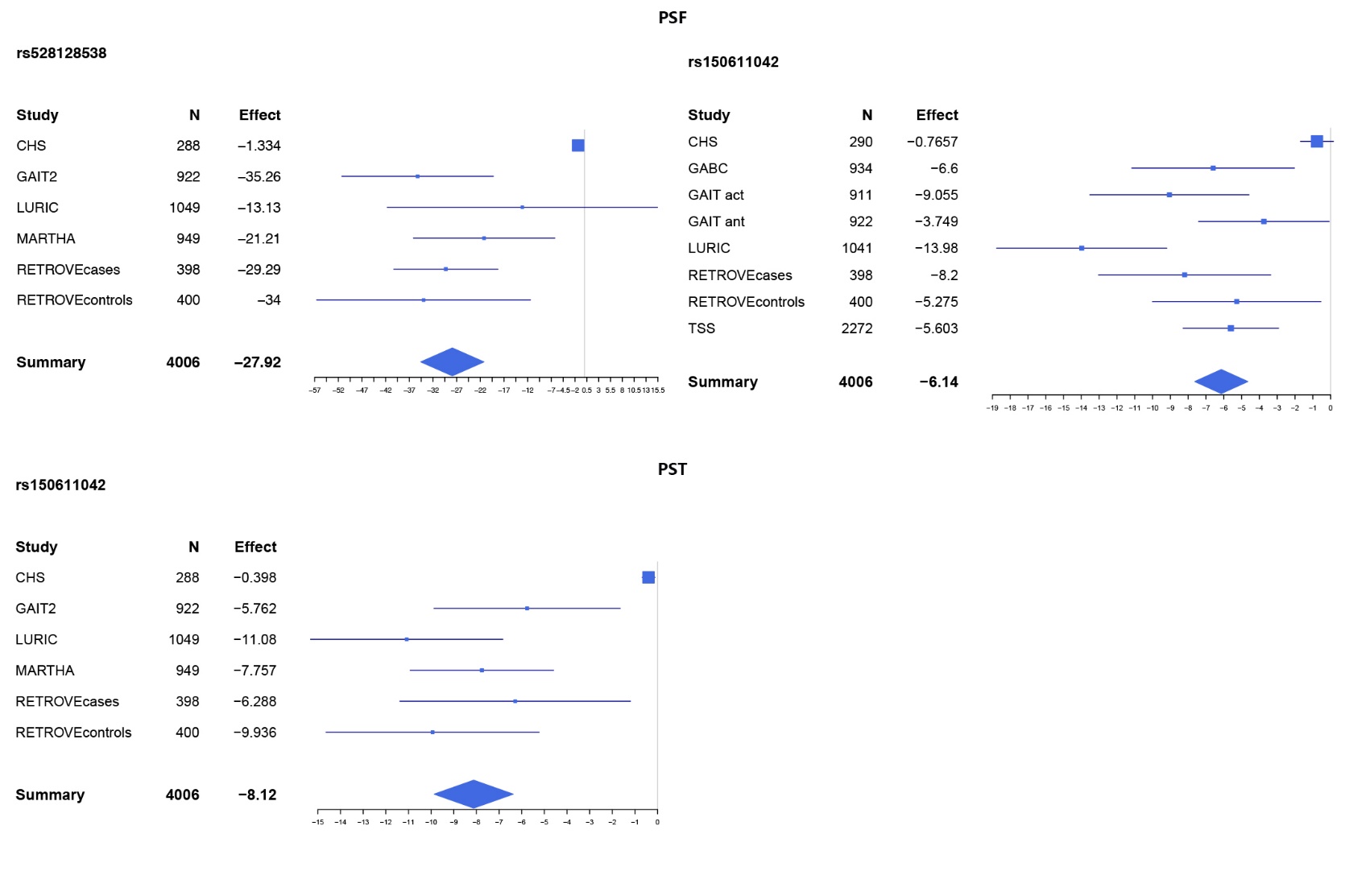

**Supplementary Figure S6. Forest plots of significant loci in Protein S free and total meta-analyses (Up: Protein S Free, down: Protein S total)**

**Funding and Acknowledgments for specific cohorts**:

**ARIC**: The ARIC study has been funded in whole or in part with Federal funds from the National Heart, Lung, and Blood Institute, National Institutes of Health, Department of Health and Human Services (contract numbers HHSN268201700001I, HHSN268201700002I, HHSN268201700003I, HHSN268201700004I and HHSN268201700005I), R01HL087641, R01HL059367 and R01HL086694; National Human Genome Research Institute contract U01HG004402; and National Institutes of Health contract HHSN268200625226C. The authors thank the staff and participants of the ARIC study for their important contributions. Infrastructure was partly supported by Grant Number UL1RR025005, a component of the National Institutes of Health and NIH Roadmap for Medical Research.

**CHRIS**: The CHRIS study was funded by the Department of Innovation, Research, and University of the Autonomous Province of Bolzano/Bozen. CHRIS study investigators thank all study participants, all general practitioners, the personnel of the Hospital of Silandro/Schlanders, and the personnel of the South Tyrolean Healthcare System for their participation and collaboration in the project. Full acknowledgements for the CHRIS study are reported elsewhere^24^.

**CHS**: This CHS research was supported by NHLBI contracts HHSN268201200036C, HHSN268200800007C, HHSN268201800001C, N01HC55222, N01HC85079, N01HC85080, N01HC85081, N01HC85082, N01HC85083, N01HC85086, 75N92021D00006; and NHLBI grants U01HL080295, R01HL087652, R01HL105756, R01HL103612, R01HL120393, and U01HL130114 with additional contribution from the National Institute of Neurological Disorders and Stroke (NINDS). Additional support was provided through R01AG023629 from the National Institute on Aging (NIA). A full list of principal CHS investigators and institutions can be found at CHS-NHLBI.org. The provision of genotyping data was supported in part by the National Center for Advancing Translational Sciences, CTSI grant UL1TR001881, and the National Institute of Diabetes and Digestive and Kidney Disease Diabetes Research Center (DRC) grant DK063491 to the Southern California Diabetes Endocrinology Research Center. The content is solely the responsibility of the authors and does not necessarily represent the official views of the National Institutes of Health.

**GABC**: The Genes and Blood Clotting Study was supported by NIH/NHLBI grants R35 HL135793 and R01 HL141399.

**GAIT**: Genetic Analysis for Idiopathic Thrombophilia (GAIT) project was supported partially by grants PI-11/0184, PI-14/0582 and Red Investigación Cardiovascular RD12/0042/0032 from the Spanish Health Institute (ISCIII), and acknowledges current support as a Consolidated Research Group from the Government of Catalonia and CERCA Program (2017 SGR 1736), and from the non-profit organization ”Associació Activa´TT per la Salut”.

**HVH**: The Health and Vascular Health Study was supported by National Heart, Lung, and Blood Institute grants HL43201, HL60739, HL68986, HL73410, HL74745, HL85251, HL95080, and HL121414.

**LURIC**: LURIC was supported by the 7th Framework Program RiskyCAD (grant agreement number 305739) of the European Union and the H2020 Program TO_AITION (grant agreement number 848146) of the European Union. The work of G.E.D. is supported by the European Union’s Horizon 2020 research and innovation programme under the ERA-Net Cofund action N° 727565 (OCTOPUS project) and the German Ministry of Education and Research (grant number 01EA1801A). The work of APM, MEK and WM was supported by the German Ministry for Education and Research as part of the Competence Cluster for Nutrition and Cardiovascular Health (nutriCARD) Halle-Jena-Leipzig (grant numbers 01EA1411A and 01EA1808A)

The **MARTHA** project was supported by a grant from the Program Hospitalier de la Recherche Clinique and by the GENMED Laboratory of Excellence on Medical Genomics [ANR-10-LABX-0013], a research program managed by the National Research Agency (ANR) as part of the French Investment for the Future. D-AT was supported by the  EPIDEMIOM-VT Senior Chair from the University of Bordeaux initiative of excellence IdEX.

**RETROVE** acknowledges support from the Government of Catalonia and CERCA Program (2017 SGR 1736), from the Spanish Health Institute (ISCIII grants PI12/000612, PI15/00269, PI18/00434 and PI 21/01772), and from the non-profit organization ”Associació Activa´TT per la Salut”. The genotyping service was carried out at CEGEN-PRB3-ISCIII; and it is supported by grant PT17/0019, of the PE I+D+i 2013-2016, funded by ISCIII and ERDF.

**TSS**: This study was supported by the Intramural Research Programs of the National Institutes of Health, the Eunice Kennedy Shriver National Institute of Child Health and Human Development, and the National Human Genome Research Institute.

**REFERENCES**

1.	Willer CJ, Li Y, Abecasis GR. METAL: fast and efficient meta-analysis of genomewide association scans. *Bioinformatics*. 2010;26:2190-2191. doi: 10.1093/bioinformatics/btq340

2.	Auton A, Brooks LD, Durbin RM, Garrison EP, Kang HM, Korbel JO, Marchini JL, McCarthy S, McVean GA, Abecasis GR. A global reference for human genetic variation. *Nature*. 2015;526:68-74. doi: 10.1038/nature15393

3.	The Genotype-Tissue Expression (GTEx) project. *Nat Genet*. 2013;45:580-585. doi: 10.1038/ng.2653

4.	Barbeira AN, Dickinson SP, Bonazzola R, Zheng J, Wheeler HE, Torres JM, Torstenson ES, Shah KP, Garcia T, Edwards TL, et al. Exploring the phenotypic consequences of tissue specific gene expression variation inferred from GWAS summary statistics. *Nat Commun*. 2018;9:1825. doi: 10.1038/s41467-018-03621-1

5.	Barbeira AN, Bonazzola R, Gamazon ER, Liang Y, Park Y, Kim-Hellmuth S, Wang G, Jiang Z, Zhou D, Hormozdiari F, et al. Exploiting the GTEx resources to decipher the mechanisms at GWAS loci. *Genome Biol*. 2021;22:49. doi: 10.1186/s13059-020-02252-4

6.	Gamazon ER, Wheeler HE, Shah KP, Mozaffari SV, Aquino-Michaels K, Carroll RJ, Eyler AE, Denny JC, Nicolae DL, Cox NJ, et al. A gene-based association method for mapping traits using reference transcriptome data. *Nat Genet*. 2015;47:1091-1098. doi: 10.1038/ng.3367

7.	Urbut SM, Wang G, Carbonetto P, Stephens M. Flexible statistical methods for estimating and testing effects in genomic studies with multiple conditions. *Nat Genet*. 2019;51:187-195. doi: 10.1038/s41588-018-0268-8

8.	Barbeira AN, Pividori M, Zheng J, Wheeler HE, Nicolae DL, Im HK. Integrating predicted transcriptome from multiple tissues improves association detection. *PLoS Genet*. 2019;15:e1007889. doi: 10.1371/journal.pgen.1007889

9.	Mancuso N, Freund MK, Johnson R, Shi H, Kichaev G, Gusev A, Pasaniuc B. Probabilistic fine-mapping of transcriptome-wide association studies. *Nat Genet*. 2019;51:675-682. doi: 10.1038/s41588-019-0367-1

10.	nygcesearch/ IDetect. <https://bitbucket.org/nygcresearch/ldetect>. Accessed April 06.

11.	Giambartolomei C, Vukcevic D, Schadt EE, Franke L, Hingorani AD, Wallace C, Plagnol V. Bayesian test for colocalisation between pairs of genetic association studies using summary statistics. *PLoS genetics*. 2014;10:e1004383.

12.	Foley CN, Staley JR, Breen PG, Sun BB, Kirk PD, Burgess S, Howson JM. A fast and efficient colocalization algorithm for identifying shared genetic risk factors across multiple traits. *Nature communications*. 2021;12:1-18.

13.	Klarin D, Lynch J, Aragam K, Chaffin M, Assimes TL, Huang J, Lee KM, Shao Q, Huffman JE, Natarajan P, et al. Genome-wide association study of peripheral artery disease in the Million Veteran Program. *Nat Med*. 2019;25:1274-1279. doi: 10.1038/s41591-019-0492-5

14.	Nikpay M, Goel A, Won HH, Hall LM, Willenborg C, Kanoni S, Saleheen D, Kyriakou T, Nelson CP, Hopewell JC, et al. A comprehensive 1,000 Genomes-based genome-wide association meta-analysis of coronary artery disease. *Nat Genet*. 2015;47:1121-1130. doi: 10.1038/ng.3396

15.	Malik R, Chauhan G, Traylor M, Sargurupremraj M, Okada Y, Mishra A, Rutten-Jacobs L, Giese AK, van der Laan SW, Gretarsdottir S, et al. Multiancestry genome-wide association study of 520,000 subjects identifies 32 loci associated with stroke and stroke subtypes. *Nat Genet*. 2018;50:524-537. doi: 10.1038/s41588-018-0058-3

16.	Lindström S, Wang L, Smith EN, Gordon W, van Hylckama Vlieg A, de Andrade M, Brody JA, Pattee JW, Haessler J, Brumpton BM, et al. Genomic and transcriptomic association studies identify 16 novel susceptibility loci for venous thromboembolism. *Blood*. 2019;134:1645-1657. doi: 10.1182/blood.2019000435

17.	Klarin D, Busenkell E, Judy R, Lynch J, Levin M, Haessler J, Aragam K, Chaffin M, Haas M, Lindström S, et al. Genome-wide association analysis of venous thromboembolism identifies new risk loci and genetic overlap with arterial vascular disease. *Nat Genet*. 2019;51:1574-1579. doi: 10.1038/s41588-019-0519-3

18.	Deloukas P, Kanoni S, Willenborg C, Farrall M, Assimes TL, Thompson JR, Ingelsson E, Saleheen D, Erdmann J, Goldstein BA, et al. Large-scale association analysis identifies new risk loci for coronary artery disease. *Nat Genet*. 2013;45:25-33. doi: 10.1038/ng.2480

19.	Bowden J, Davey Smith G, Haycock PC, Burgess S. Consistent Estimation in Mendelian Randomization with Some Invalid Instruments Using a Weighted Median Estimator. *Genet Epidemiol*. 2016;40:304-314. doi: 10.1002/gepi.21965

20.	Hartwig FP, Davey Smith G, Bowden J. Robust inference in summary data Mendelian randomization via the zero modal pleiotropy assumption. *Int J Epidemiol*. 2017;46:1985-1998. doi: 10.1093/ije/dyx102

21.	Bowden J, Davey Smith G, Burgess S. Mendelian randomization with invalid instruments: effect estimation and bias detection through Egger regression. *Int J Epidemiol*. 2015;44:512-525. doi: 10.1093/ije/dyv080

22.	Stacey D, Chen L, Howson JM, Mason AM, Burgess S, MacDonald S, Langdown J, McKinney H, Downes K, Farahi N. Elucidating mechanisms of genetic cross-disease associations: an integrative approach implicates protein C as a causal pathway in arterial and venous diseases. *medRxiv*. 2020.

23.	Wright JD, Folsom AR, Coresh J, et al. The ARIC (Atherosclerosis Risk In Communities) Study: JACC Focus Seminar 3/8. *J Am Coll Cardiol.* 2021 Jun 15;77(23):2939-2959)

24.	Pattaro C, Gögele M, Mascalzoni D, Melotti R, Schwienbacher C, De Grandi A, Foco L, D’elia Y, Linder B, Fuchsberger C. The Cooperative Health Research in South Tyrol (CHRIS) study: rationale, objectives, and preliminary results. *Journal of translational medicine*. 2015;13:1-16.

25.	Fried LP, Borhani NO, Enright P, Furberg CD, Gardin JM, Kronmal RA, Kuller LH, Manolio TA, Mittelmark MB, Newman A, et al. The Cardiovascular Health Study: design and rationale. *Ann Epidemiol*. 1991;1:263-276. doi: 10.1016/1047-2797(91)90005-w

26.	Cushman M, Cornell ES, Howard PR, Bovill EG, Tracy RP. Laboratory methods and quality assurance in the Cardiovascular Health Study. *Clin Chem*. 1995;41:264-270.

27.	Desch K, Li J, Kim S, Laventhal N, Metzger K, Siemieniak D, Ginsburg D. Analysis of informed consent document utilization in a minimal-risk genetic study. *Annals of internal medicine*. 2011;155:316-322.

28.	Desch KC, Ozel AB, Siemieniak D, Kalish Y, Shavit JA, Thornburg CD, Sharathkumar AA, McHugh CP, Laurie CC, Crenshaw A, et al. Linkage analysis identifies a locus for plasma von Willebrand factor undetected by genome-wide association. *Proc Natl Acad Sci U S A*. 2013;110:588-593. doi: 10.1073/pnas.1219885110

29.	Souto JC, Almasy L, Borrell M, Garí M, Martínez E, Mateo J, Stone WH, Blangero J, Fontcuberta J. Genetic determinants of hemostasis phenotypes in Spanish families. *Circulation*. 2000;101:1546-1551. doi: 10.1161/01.cir.101.13.1546

30.	Souto JC, Almasy L, Borrell M, Blanco-Vaca F, Mateo J, Soria JM, Coll I, Felices R, Stone W, Fontcuberta J, et al. Genetic susceptibility to thrombosis and its relationship to physiological risk factors: the GAIT study. Genetic Analysis of Idiopathic Thrombophilia. *Am J Hum Genet*. 2000;67:1452-1459. doi: 10.1086/316903

31.	Psaty BM, Heckbert SR, Koepsell TD, Siscovick DS, Raghunathan TE, Weiss NS, Rosendaal FR, Lemaitre RN, Smith NL, Wahl PW, et al. The risk of myocardial infarction associated with antihypertensive drug therapies. *Jama*. 1995;274:620-625.

32.	Psaty BM, Heckbert SR, Atkins D, Lemaitre R, Koepsell TD, Wahl PW, Siscovick DS, Wagner EH. The risk of myocardial infarction associated with the combined use of estrogens and progestins in postmenopausal women. *Arch Intern Med*. 1994;154:1333-1339.

33.	Heckbert SR, Weiss NS, Koepsell TD, Lemaitre RN, Smith NL, Siscovick DS, Lin D, Psaty BM. Duration of estrogen replacement therapy in relation to the risk of incident myocardial infarction in postmenopausal women. *Arch Intern Med*. 1997;157:1330-1336.

34.	Heckbert SR, Wiggins KL, Glazer NL, Dublin S, Psaty BM, Smith NL, Longstreth WT, Jr., Lumley T. Antihypertensive treatment with ACE inhibitors or beta-blockers and risk of incident atrial fibrillation in a general hypertensive population. *Am J Hypertens*. 2009;22:538-544. doi: 10.1038/ajh.2009.33

35.	Lemaitre RN, Heckbert SR, Psaty BM, Smith NL, Kaplan RC, Longstreth WT, Jr. Hormone replacement therapy and associated risk of stroke in postmenopausal women. *Arch Intern Med*. 2002;162:1954-1960. doi: 10.1001/archinte.162.17.1954

36.	Smith NL, Blondon M, Wiggins KL, Harrington LB, van Hylckama Vlieg A, Floyd JS, Hwang M, Bis JC, McKnight B, Rice KM, et al. Lower risk of cardiovascular events in postmenopausal women taking oral estradiol compared with oral conjugated equine estrogens. *JAMA Intern Med*. 2014;174:25-31. doi: 10.1001/jamainternmed.2013.11074

37.	Blondon M, van Hylckama Vlieg A, Wiggins KL, Harrington LB, McKnight B, Rice KM, Rosendaal FR, Heckbert SR, Psaty BM, Smith NL. Differential associations of oral estradiol and conjugated equine estrogen with hemostatic biomarkers. *J Thromb Haemost*. 2014;12:879-886. doi: 10.1111/jth.12560

38.	Tomaschitz A, Pilz S, Ritz E, Meinitzer A, Boehm BO, März W. Plasma aldosterone levels are associated with increased cardiovascular mortality: the Ludwigshafen Risk and Cardiovascular Health (LURIC) study. *European heart journal*. 2010;31:1237-1247.

39.	Antoni G, Oudot-Mellakh T, Dimitromanolakis A, Germain M, Cohen W, Wells P, Lathrop M, Gagnon F, Morange PE, Tregouet DA. Combined analysis of three genome-wide association studies on vWF and FVIII plasma levels. *BMC Med Genet*. 2011;12:102. doi: 10.1186/1471-2350-12-102

40.	Vázquez-Santiago M, Vilalta N, Cuevas B, Murillo J, Llobet D, Macho R, Pujol-Moix N, Carrasco M, Mateo J, Fontcuberta J, et al. Short closure time values in PFA-100® are related to venous thrombotic risk. Results from the RETROVE Study. *Thromb Res*. 2018;169:57-63. doi: 10.1016/j.thromres.2018.07.012

41.	Mills JL, Carter TC, Scott JM, Troendle JF, Gibney ER, Shane B, Kirke PN, Ueland PM, Brody LC, Molloy AM. Do high blood folate concentrations exacerbate metabolic abnormalities in people with low vitamin B-12 status? *The American journal of clinical nutrition*. 2011;94:495-500.

42.	Stone N, Pangilinan F, Molloy AM, Shane B, Scott JM, Ueland PM, Mills JL, Kirke PN, Sethupathy P, Brody LC. Bioinformatic and genetic association analysis of microRNA target sites in one-carbon metabolism genes. *PloS one*. 2011;6:e21851.

**MEGASTROKE CONSORTIUM AUTHORS**

Rainer Malik ^1^, Ganesh Chauhan ^2^, Matthew Traylor ^3^, Muralidharan Sargurupremraj ^4,5^, Yukinori Okada ^6,7,8^, Aniket Mishra ^4,5^, Loes Rutten-Jacobs ^3^, Anne-Katrin Giese ^9^, Sander W van der Laan ^10^, Solveig Gretarsdottir ^11^, Christopher D Anderson ^12,13,14,14^, Michael Chong ^15^, Hieab HH Adams ^16,17^, Tetsuro Ago ^18^, Peter Almgren ^19^, Philippe Amouyel ^20,21^, Hakan Ay ^22,13^, Traci M Bartz ^23^, Oscar R Benavente ^24^, Steve Bevan ^25^, Giorgio B Boncoraglio ^26^, Robert D Brown, Jr.  ^27^, Adam S Butterworth ^28,29^, Caty Carrera ^30,31^, Cara L Carty ^32,33^, Daniel I Chasman ^34,35^, Wei-Min Chen ^36^, John W Cole ^37^, Adolfo Correa ^38^, Ioana Cotlarciuc ^39^, Carlos Cruchaga ^40,41^, John Danesh ^28,42,43,44^, Paul IW de Bakker ^45,46^, Anita L DeStefano ^47,48^, Marcel den Hoed ^49^, Qing Duan ^50^, Stefan T Engelter ^51,52^, Guido J Falcone ^53,54^, Rebecca F Gottesman ^55^, Raji P Grewal ^56^, Vilmundur Gudnason ^57,58^, Stefan Gustafsson ^59^, Jeffrey Haessler ^60^, Tamara B Harris ^61^, Ahamad Hassan ^62^, Aki S Havulinna ^63,64^, Susan R Heckbert ^65^, Elizabeth G Holliday ^66,67^, George Howard ^68^, Fang-Chi Hsu ^69^, Hyacinth I Hyacinth ^70^, M Arfan Ikram ^16^, Erik Ingelsson ^71,72^, Marguerite R Irvin ^73^, Xueqiu Jian ^74^, Jordi Jiménez-Conde ^75^, Julie A Johnson ^76,77^, J Wouter Jukema ^78^, Masahiro Kanai ^6,7,79^, Keith L Keene ^80,81^, Brett M Kissela ^82^, Dawn O Kleindorfer ^82^, Charles Kooperberg ^60^, Michiaki Kubo ^83^, Leslie A Lange ^84^, Carl D Langefeld ^85^, Claudia Langenberg ^86^, Lenore J Launer ^87^, Jin-Moo Lee ^88^, Robin Lemmens ^89,90^, Didier Leys ^91^, Cathryn M Lewis ^92,93^, Wei-Yu Lin ^28,94^, Arne G Lindgren ^95,96^, Erik Lorentzen ^97^, Patrik K Magnusson ^98^, Jane Maguire ^99^, Ani Manichaikul ^36^, Patrick F McArdle ^100^, James F Meschia ^101^, Braxton D Mitchell ^100,102^, Thomas H Mosley ^103,104^, Michael A Nalls ^105,106^, Toshiharu Ninomiya ^107^, Martin J O'Donnell ^15,108^, Bruce M Psaty ^109,110,111,112^, Sara L Pulit ^113,45^, Kristiina Rannikmäe ^114,115^, Alexander P Reiner ^65,116^, Kathryn M Rexrode ^117^, Kenneth Rice ^118^, Stephen S Rich ^36^, Paul M Ridker ^34,35^, Natalia S Rost ^9,13^, Peter M Rothwell ^119^, Jerome I Rotter ^120,121^, Tatjana Rundek ^122^, Ralph L Sacco ^122^, Saori Sakaue ^7,123^, Michele M Sale ^124^, Veikko Salomaa ^63^, Bishwa R Sapkota ^125^, Reinhold Schmidt ^126^, Carsten O Schmidt  ^127^, Ulf Schminke ^128^, Pankaj Sharma ^39^, Agnieszka Slowik ^129^, Cathie LM Sudlow ^114,115^, Christian Tanislav ^130^, Turgut Tatlisumak ^131,132^, Kent D Taylor ^120,121^, Vincent NS Thijs ^133,134^, Gudmar Thorleifsson ^11^, Unnur Thorsteinsdottir ^11^, Steffen Tiedt ^1^, Stella Trompet ^135^, Christophe Tzourio ^5,136,137^, Cornelia M van Duijn ^138,139^, Matthew Walters ^140^, Nicholas J Wareham ^86^, Sylvia Wassertheil-Smoller ^141^, James G Wilson ^142^, Kerri L Wiggins ^109^, Qiong Yang ^47^, Salim Yusuf ^15^, Najaf Amin ^16^, Hugo S Aparicio ^185,48^, Donna K Arnett ^186^, John Attia ^187^, Alexa S Beiser ^47,48^, Claudine Berr ^188^, Julie E Buring ^34,35^, Mariana Bustamante ^189^, Valeria Caso ^190^, Yu-Ching Cheng ^191^, Seung Hoan Choi ^192,48^, Ayesha Chowhan ^185,48^, Natalia Cullell ^31^, Jean-François Dartigues ^193,194^, Hossein Delavaran ^95,96^, Pilar Delgado ^195^, Marcus Dörr ^196,197^, Gunnar Engström ^19^, Ian Ford ^198^, Wander S Gurpreet ^199^, Anders Hamsten ^200,201^, Laura Heitsch ^202^, Atsushi Hozawa ^203^, Laura Ibanez ^204^, Andreea Ilinca ^95,96^, Martin Ingelsson ^205^, Motoki Iwasaki ^206^, Rebecca D Jackson ^207^, Katarina Jood ^208^, Pekka Jousilahti ^63^, Sara Kaffashian ^4,5^, Lalit Kalra ^209^, Masahiro Kamouchi ^210^, Takanari Kitazono ^211^, Olafur Kjartansson ^212^, Manja Kloss ^213^, Peter J Koudstaal ^214^, Jerzy Krupinski ^215^, Daniel L Labovitz ^216^, Cathy C Laurie ^118^, Christopher R Levi ^217^, Linxin Li ^218^, Lars Lind ^219^, Cecilia M Lindgren ^220,221^, Vasileios Lioutas ^222,48^, Yong Mei Liu ^223^, Oscar L Lopez ^224^, Hirata Makoto ^225^, Nicolas Martinez-Majander ^172^, Koichi Matsuda ^225^, Naoko Minegishi ^203^, Joan Montaner  ^226^, Andrew P Morris ^227,228^, Elena Muiño ^31^, Martina Müller-Nurasyid ^229,230,231^, Bo Norrving ^95,96^, Soichi Ogishima ^203^, Eugenio A Parati ^232^, Leema Reddy Peddareddygari ^56^, Nancy L Pedersen ^98,233^, Joanna Pera ^129^, Markus Perola ^63,234^, Alessandro Pezzini ^235^, Silvana Pileggi ^236^, Raquel Rabionet ^237^, Iolanda Riba-Llena ^30^, Marta Ribasés ^238^, Jose R Romero ^185,48^, Jaume Roquer ^239,240^, Anthony G Rudd ^241,242^, Antti-Pekka Sarin ^243,244^, Ralhan Sarju ^199^, Chloe Sarnowski ^47,48^, Makoto Sasaki ^245^, Claudia L Satizabal ^185,48^, Mamoru Satoh ^245^, Naveed Sattar ^246^, Norie Sawada ^206^, Gerli Sibolt ^172^, Ásgeir Sigurdsson ^247^, Albert Smith ^248^, Kenji Sobue ^245^, Carolina Soriano-Tárraga ^240^, Tara Stanne ^249^, O Colin Stine ^250^, David J Stott ^251^, Konstantin Strauch ^229,252^, Takako Takai  ^203^, Hideo Tanaka ^253,254^, Kozo Tanno ^245^, Alexander Teumer ^255^, Liisa Tomppo ^172^, Nuria P Torres-Aguila ^31^, Emmanuel Touze ^256,257^, Shoichiro Tsugane  ^206^, Andre G Uitterlinden ^258^, Einar M Valdimarsson ^259^, Sven J van der Lee ^16^, Henry Völzke ^255^, Kenji Wakai  ^253^, David Weir ^260^, Stephen R Williams ^261^, Charles DA Wolfe ^241,242^, Quenna Wong ^118^, Huichun Xu ^191^, Taiki Yamaji ^206^, Dharambir K Sanghera ^125,169,170^, Olle Melander ^19^, Christina Jern ^171^, Daniel Strbian ^172,173^, Israel Fernandez-Cadenas ^31,30^, W T Longstreth, Jr ^174,65^, Arndt Rolfs ^175^, Jun Hata ^107^, Daniel Woo ^82^, Jonathan Rosand ^12,13,14^, Guillaume Pare ^15^, Jemma C Hopewell ^176^, Danish Saleheen ^177^, Kari Stefansson ^11,178^, Bradford B Worrall ^179^, Steven J Kittner ^37^, Sudha Seshadri ^180,48^, Myriam Fornage ^74,181^, Hugh S Markus ^3^, Joanna MM Howson ^28^, Yoichiro Kamatani ^6,182^, Stephanie Debette ^4,5^, Martin Dichgans ^1,183,184^

1 Institute for Stroke and Dementia Research (ISD), University Hospital, LMU Munich, Munich, Germany

2 Centre for Brain Research, Indian Institute of Science, Bangalore, India

3 Stroke Research Group, Division of Clinical Neurosciences, University of Cambridge, UK

4 INSERM U1219 Bordeaux Population Health Research Center, Bordeaux, France

5 University of Bordeaux, Bordeaux, France

6 Laboratory for Statistical Analysis, RIKEN Center for Integrative Medical Sciences, Yokohama, Japan

7 Department of Statistical Genetics, Osaka University Graduate School of Medicine, Osaka, Japan

8 Laboratory of Statistical Immunology, Immunology Frontier Research Center (WPI-IFReC), Osaka University, Suita, Japan.

9 Department of Neurology, Massachusetts General Hospital, Harvard Medical School, Boston, MA, USA

10 Laboratory of Experimental Cardiology, Division of Heart and Lungs, University Medical Center Utrecht, University of Utrecht, Utrecht,Netherlands

11 deCODE genetics/AMGEN inc, Reykjavik, Iceland

12 Center for Genomic Medicine, Massachusetts General Hospital (MGH), Boston, MA, USA

13 J. Philip Kistler Stroke Research Center, Department of Neurology, MGH, Boston, MA, USA

14 Program in Medical and Population Genetics, Broad Institute, Cambridge, MA, USA

15 Population Health Research Institute, McMaster University, Hamilton, Canada

16 Department of Epidemiology, Erasmus University Medical Center, Rotterdam, Netherlands

17 Department of Radiology and Nuclear Medicine, Erasmus University Medical Center, Rotterdam, Netherlands

18 Department of Medicine and Clinical Science, Graduate School of Medical Sciences, Kyushu University, Fukuoka, Japan

19 Department of Clinical Sciences, Lund University, Malmö, Sweden

20 Univ. Lille, Inserm, Institut Pasteur de Lille, LabEx DISTALZ-UMR1167, Risk factors and molecular determinants of aging-related diseases, F-59000 Lille, France

21 Centre Hosp. Univ Lille, Epidemiology and Public Health Department, F-59000 Lille, France

22 AA Martinos Center for Biomedical Imaging, Department of Radiology, Massachusetts General Hospital, Harvard Medical School, Boston, MA, USA

23 Cardiovascular Health Research Unit, Departments of Biostatistics and Medicine, University of Washington, Seattle, WA, USA

24 Division of Neurology, Faculty of Medicine, Brain Research Center, University of British Columbia, Vancouver, Canada

25 School of Life Science, University of Lincoln, Lincoln, UK

26 Department of Cerebrovascular Diseases, Fondazione IRCCS Istituto Neurologico "Carlo Besta", Milano, Italy

27 Department of Neurology, Mayo Clinic Rochester, Rochester, MN, USA

28 MRC/BHF Cardiovascular Epidemiology Unit, Department of Public Health and Primary Care, University of Cambridge, Cambridge, UK

29 The National Institute for Health Research Blood and Transplant Research Unit in Donor Health and Genomics, University of Cambridge, UK

30 Neurovascular Research Laboratory, Vall d'Hebron Institut of Research, Neurology and Medicine Departments-Universitat Autònoma de Barcelona, Vall d’Hebrón Hospital, Barcelona, Spain

31 Stroke Pharmacogenomics and Genetics, Fundacio Docència i Recerca MutuaTerrassa, Terrassa, Spain

32 Children's Research Institute, Children's National Medical Center, Washington, DC, USA

33 Center for Translational Science, George Washington University, Washington, DC, USA

34 Division of Preventive Medicine, Brigham and Women's Hospital, Boston, MA, USA

35 Harvard Medical School, Boston, MA, USA

36 Center for Public Health Genomics, Department of Public Health Sciences, University of Virginia, Charlottesville, VA, USA

37 Department of Neurology, University of Maryland School of Medicine and Baltimore VAMC, Baltimore, MD, USA

38 Departments of Medicine, Pediatrics and Population Health Science, University of Mississippi Medical Center, Jackson, MS, USA

39 Institute of Cardiovascular Research, Royal Holloway University of London, UK  &  Ashford and St Peters Hospital, Surrey UK

40 Department of Psychiatry,The Hope Center Program on Protein Aggregation and Neurodegeneration (HPAN),Washington University, School of Medicine, St. Louis, MO, USA

41 Department of Developmental Biology, Washington University School of Medicine, St. Louis, MO, USA

42 NIHR Blood and Transplant Research Unit in Donor Health and Genomics, Department of Public Health and Primary Care, University of Cambridge, Cambridge, UK

43 Wellcome Trust Sanger Institute, Wellcome Trust Genome Campus, Hinxton,  Cambridge, UK

44 British Heart Foundation, Cambridge Centre of Excellence, Department of Medicine, University of Cambridge, Cambridge, UK

45 Department of Medical Genetics, University Medical Center Utrecht, Utrecht, Netherlands

46 Department of Epidemiology, Julius Center for Health Sciences and Primary Care, University Medical Center Utrecht, Utrecht, Netherlands

47 Boston University School of Public Health, Boston, MA, USA

48 Framingham Heart Study, Framingham, MA, USA

49 Department of Immunology, Genetics and Pathology and Science for Life Laboratory, Uppsala University, Uppsala, Sweden

50 Department of Genetics, University of North Carolina, Chapel Hill, NC, USA

51 Department of Neurology and Stroke Center, Basel University Hospital, Switzerland

52 Neurorehabilitation Unit, University and University Center for Medicine of Aging and Rehabilitation Basel, Felix Platter Hospital, Basel, Switzerland

53 Department of Neurology, Yale University School of Medicine, New Haven, CT, USA

54 Program in Medical and Population Genetics, The Broad Institute of Harvard and MIT, Cambridge, MA, USA

55 Department of Neurology, Johns Hopkins University School of Medicine, Baltimore, MD, USA

56 Neuroscience Institute, SF Medical Center, Trenton, NJ, USA

57 Icelandic Heart Association Research Institute, Kopavogur, Iceland

58 University of Iceland, Faculty of Medicine, Reykjavik, Iceland

59 Department of Medical Sciences, Molecular Epidemiology and Science for Life Laboratory, Uppsala University, Uppsala, Sweden

60 Division of Public Health Sciences, Fred Hutchinson Cancer Research Center, Seattle, WA, USA

61 Laboratory of Epidemiology and Population Science, National Institute on Aging, National Institutes of Health, Bethesda, MD, USA

62 Department of Neurology, Leeds General Infirmary, Leeds Teaching Hospitals NHS Trust, Leeds, UK

63 National Institute for Health and Welfare, Helsinki, Finland

64 FIMM - Institute for Molecular Medicine Finland, Helsinki, Finland

65 Department of Epidemiology, University of Washington, Seattle, WA, USA

66 Public Health Stream, Hunter Medical Research Institute, New Lambton, Australia

67 Faculty of Health and Medicine, University of Newcastle, Newcastle, Australia

68 School of Public Health, University of Alabama at Birmingham, Birmingham, AL, USA

69 Department of Biostatistical Sciences, Wake Forest School of Medicine, Winston-Salem, NC, USA

70 Aflac Cancer and Blood Disorder Center, Department of Pediatrics, Emory University School of Medicine, Atlanta, GA, USA

71 Department of Medicine, Division of Cardiovascular Medicine, Stanford University School of Medicine, CA, USA

72 Department of Medical Sciences, Molecular Epidemiology and Science for Life Laboratory, Uppsala University, Uppsala, Sweden

73 Epidemiology, School of Public Health, University of Alabama at Birmingham, USA

74 Brown Foundation Institute of Molecular Medicine, University of Texas Health Science Center at Houston, Houston, TX, USA

75 Neurovascular Research Group (NEUVAS), Neurology Department, Institut Hospital del Mar d'Investigació Mèdica, Universitat Autònoma de Barcelona, Barcelona, Spain

76 Department of Pharmacotherapy and Translational Research and Center for Pharmacogenomics, University of Florida, College of Pharmacy, Gainesville, FL, USA

77 Division of Cardiovascular Medicine, College of Medicine, University of Florida, Gainesville, FL, USA

78 Department of Cardiology, Leiden University Medical Center, Leiden, the Netherlands

79 Program in Bioinformatics and Integrative Genomics, Harvard Medical School, Boston, MA, USA

80 Department of Biology, East Carolina University, Greenville, NC, USA

81 Center for Health Disparities, East Carolina University, Greenville, NC, USA

82 University of Cincinnati College of Medicine, Cincinnati, OH, USA

83 RIKEN Center for Integrative Medical Sciences, Yokohama, Japan

84 Department of Medicine, University of Colorado Denver, Anschutz Medical Campus, Aurora, CO, USA

85 Center for Public Health Genomics and Department of Biostatistical Sciences, Wake Forest School of Medicine, Winston-Salem, NC, USA

86 MRC Epidemiology Unit, University of Cambridge School of Clinical Medicine, Institute of Metabolic Science, Cambridge Biomedical Campus, Cambridge, UK

87 Intramural Research Program, National Institute on Aging, National Institutes of Health, Bethesda, MD, USA

88 Department of Neurology, Radiology, and Biomedical Engineering, Washington University School of Medicine, St. Louis, MO, USA

89 KU Leuven – University of Leuven, Department of Neurosciences,  Experimental Neurology, Leuven, Belgium

90 VIB Center for Brain & Disease Research, University Hospitals Leuven, Department of Neurology, Leuven, Belgium

91 Univ.-Lille, INSERM U 1171. CHU Lille. Lille, France

92 Department of Medical and Molecular Genetics, King's College London, London, UK

93 SGDP Centre, Institute of Psychiatry, Psychology & Neuroscience, King's College London, London, UK

94 Northern Institute for Cancer Research, Paul O'Gorman Building, Newcastle University, Newcastle, UK

95 Department of Clinical Sciences Lund, Neurology, Lund University, Lund, Sweden

96 Department of Neurology and Rehabilitation Medicine, Skåne University Hospital, Lund, Sweden

97 Bioinformatics Core Facility, University of Gothenburg, Gothenburg, Sweden

98 Department of Medical Epidemiology and Biostatistics, Karolinska Institutet, Stockholm, Sweden

99 University of Technology Sydney, Faculty of Health, Ultimo, Australia

100 Department of Medicine, University of Maryland School of Medicine, MD, USA

101 Department of Neurology, Mayo Clinic, Jacksonville, FL, USA

102 Geriatrics Research and Education Clinical Center, Baltimore Veterans Administration Medical Center, Baltimore, MD, USA

103 Division of Geriatrics, School of Medicine, University of Mississippi Medical Center, Jackson, MS, USA

104 Memory Impairment and Neurodegenerative Dementia Center, University of Mississippi Medical Center, Jackson, MS, USA

105 Laboratory of Neurogenetics, National Institute on Aging, National institutes of Health, Bethesda, MD, USA

106 Data Tecnica International, Glen Echo MD, USA

107 Department of Epidemiology and Public Health, Graduate School of Medical Sciences, Kyushu University, Fukuoka, Japan

108 Clinical Research Facility, Department of Medicine, NUI Galway, Galway, Ireland

109 Cardiovascular Health Research Unit, Department of Medicine, University of Washington, Seattle, WA, USA

110 Department of Epidemiology, University of Washington, Seattle, WA

111 Department of Health Services, University of Washington, Seattle, WA, USA

112 Kaiser Permanente Washington Health Research Institute, Seattle, WA, USA

113 Brain Center Rudolf Magnus, Department of Neurology, University Medical Center Utrecht, Utrecht, The Netherlands

114 Usher Institute of Population Health Sciences and Informatics, University of Edinburgh, Edinburgh, UK

115 Centre for Clinical Brain Sciences, University of Edinburgh, Edinburgh, UK

116 Fred Hutchinson Cancer Research Center, University of Washington, Seattle, WA, USA

117 Department of Medicine, Brigham and Women's Hospital, Boston, MA, USA

118 Department of Biostatistics, University of Washington, Seattle, WA, USA

119 Nuffield Department of Clinical Neurosciences, University of Oxford, UK

120 Institute for Translational Genomics and Population Sciences, Los Angeles Biomedical Research Institute at  Harbor-UCLA Medical Center, Torrance, CA, USA

121 Division of Genomic Outcomes, Department of Pediatrics, Harbor-UCLA Medical Center, Torrance, CA, USA

122 Department of Neurology, Miller School of Medicine, University of Miami, Miami, FL, USA

123 Department of Allergy and Rheumatology, Graduate School of Medicine, the University of Tokyo, Tokyo, Japan

124 Center for Public Health Genomics, University of Virginia, Charlottesville, VA, USA

125 Department of Pediatrics, College of Medicine, University of Oklahoma Health Sciences Center, Oklahoma City, OK, USA

126 Department of Neurology, Medical University of Graz, Graz, Austria

127 University Medicine  Greifswald, Institute for Community Medicine, SHIP-KEF, Greifswald, Germany

128 University Medicine  Greifswald,  Department of Neurology, Greifswald, Germany

129 Department of Neurology, Jagiellonian University, Krakow, Poland

130 Department of Neurology, Justus Liebig University, Giessen, Germany

131 Department of Clinical Neurosciences/Neurology, Institute of Neuroscience and Physiology, Sahlgrenska Academy at University of Gothenburg, Gothenburg, Sweden

132 Sahlgrenska University Hospital, Gothenburg, Sweden

133 Stroke Division, Florey Institute of Neuroscience and Mental Health, University of Melbourne, Heidelberg, Australia

134 Austin Health, Department of Neurology, Heidelberg, Australia

135 Department of Internal Medicine, Section Gerontology and Geriatrics, Leiden University Medical Center, Leiden, the Netherlands

136 INSERM U1219, Bordeaux, France

137 Department of Public Health, Bordeaux University Hospital, Bordeaux, France

138 Genetic Epidemiology Unit, Department of Epidemiology, Erasmus University Medical Center Rotterdam, Netherlands

139 Center for Medical Systems Biology, Leiden, Netherlands

140 School of Medicine, Dentistry and Nursing at the University of Glasgow, Glasgow, UK

141 Department of Epidemiology and Population Health, Albert Einstein College of Medicine, NY, USA

142 Department of Physiology and Biophysics, University of Mississippi Medical Center, Jackson, MS, USA

143 A full list of members and affiliations appears in the Supplementary Note

144 Department of Human Genetics, McGill University, Montreal, Canada

145 Department of Pathophysiology, Institute of Biomedicine and Translation Medicine, University of Tartu, Tartu, Estonia

146 Department of Cardiac Surgery, Tartu University Hospital, Tartu, Estonia

147 Clinical Gene Networks AB,Stockholm, Sweden

148 Department of Genetics and Genomic Sciences, The Icahn Institute for Genomics and Multiscale Biology Icahn School of Medicine at Mount Sinai, New York, NY , USA

149 Department of Pathophysiology, Institute of Biomedicine and Translation Medicine, University of Tartu, Biomeedikum, Tartu, Estonia

150 Integrated Cardio Metabolic Centre, Department of Medicine, Karolinska Institutet, Karolinska Universitetssjukhuset, Huddinge, Sweden.

151 Clinical Gene Networks AB, Stockholm, Sweden

152 Sorbonne Universités, UPMC Univ. Paris 06, INSERM, UMR_S 1166, Team Genomics & Pathophysiology of Cardiovascular Diseases, Paris, France

153 ICAN Institute for Cardiometabolism and Nutrition, Paris, France

154 Department of Biomedical Engineering, University of Virginia, Charlottesville, VA, USA

155 Group Health Research Institute, Group Health Cooperative, Seattle, WA, USA

156 Seattle Epidemiologic Research and Information Center, VA Office of Research and Development, Seattle, WA, USA

157 Cardiovascular Research Center, Massachusetts General Hospital, Boston, MA, USA

158 Department of Medical Research, Bærum Hospital, Vestre Viken Hospital Trust, Gjettum, Norway

159 Saw Swee Hock School of Public Health, National University of Singapore and National University Health System, Singapore

160 National Heart and Lung Institute, Imperial College London, London, UK

161 Department of Gene Diagnostics and Therapeutics, Research Institute, National Center for Global Health and Medicine, Tokyo, Japan

162 Department of Epidemiology, Tulane University School of Public Health and Tropical Medicine, New Orleans, LA, USA

163 Department of Cardiology,University Medical Center Groningen, University of Groningen, Netherlands

164 MRC-PHE Centre for Environment and Health, School of Public Health, Department of Epidemiology and Biostatistics, Imperial College London, London, UK

165 Department of Epidemiology and Biostatistics, Imperial College London, London, UK

166 Department of Cardiology, Ealing Hospital NHS Trust, Southall, UK

167 National Heart, Lung and Blood Research Institute, Division of Intramural Research, Population Sciences Branch, Framingham, MA, USA

168 A full list of members and affiliations appears at the end of the manuscript

169 Department of Phamaceutical Sciences, Collge of Pharmacy, University of Oklahoma Health Sciences Center, Oklahoma City, OK, USA

170 Oklahoma Center for Neuroscience, Oklahoma City, OK, USA

171 Department of Pathology and Genetics, Institute of Biomedicine, The Sahlgrenska Academy at University of Gothenburg, Gothenburg, Sweden

172 Department of Neurology, Helsinki University Hospital, Helsinki, Finland

173 Clinical Neurosciences, Neurology, University of Helsinki, Helsinki, Finland

174 Department of Neurology, University of Washington, Seattle, WA, USA

175 Albrecht Kossel Institute, University Clinic of Rostock, Rostock, Germany

176 Clinical Trial Service Unit and Epidemiological Studies Unit, Nuffield Department of Population Health, University of Oxford, Oxford, UK

177 Department of Genetics, Perelman School of Medicine, University of Pennsylvania, PA, USA

178 Faculty of Medicine, University of Iceland, Reykjavik, Iceland

179 Departments of Neurology and Public Health Sciences, University of Virginia School of Medicine, Charlottesville, VA, USA

180 Department of Neurology, Boston University School of Medicine, Boston, MA, USA

181 Human Genetics Center, University of Texas Health Science Center at Houston, Houston, TX, USA

182 Center for Genomic Medicine, Kyoto University Graduate School of Medicine, Kyoto, Japan

183 Munich Cluster for Systems Neurology (SyNergy), Munich, Germany

184 German Center for Neurodegenerative Diseases (DZNE), Munich, Germany

185 Boston University School of Medicine, Boston, MA, USA

186 University of Kentucky College of Public Health, Lexington, KY, USA

187 University of Newcastle and Hunter Medical Research Institute, New Lambton, Australia

188 Univ. Montpellier, Inserm, U1061, Montpellier, France

189 Centre for Research in Environmental Epidemiology, Barcelona, Spain

190 Department of Neurology, Università degli Studi di Perugia, Umbria, Italy

191 Department of Medicine, University of Maryland School of Medicine, Baltimore, MD, USA

192 Broad Institute, Cambridge, MA, USA

193 Univ. Bordeaux, Inserm, Bordeaux Population Health Research Center, UMR 1219, Bordeaux, France

194 Bordeaux University Hospital, Department of Neurology, Memory Clinic, Bordeaux, France

195 Neurovascular Research Laboratory. Vall d'Hebron Institut of Research, Neurology and Medicine Departments-Universitat Autònoma de Barcelona. Vall d’Hebrón Hospital, Barcelona, Spain

196 University Medicine Greifswald, Department of Internal Medicine B, Greifswald, Germany

197 DZHK, Greifswald, Germany

198 Robertson Center for Biostatistics, University of Glasgow, Glasgow, UK

199 Hero DMC Heart Institute, Dayanand Medical College & Hospital, Ludhiana, India

200 Atherosclerosis Research Unit, Department of Medicine Solna, Karolinska Institutet, Stockholm, Sweden

201 Karolinska Institutet, Stockholm, Sweden

202 Division of Emergency Medicine, and Department of Neurology, Washington University School of Medicine, St. Louis, MO, USA

203 Tohoku Medical Megabank Organization, Sendai, Japan

204 Department of Psychiatry, Washington University School of Medicine, St. Louis, MO, USA

205 Department of Public Health and Caring Sciences / Geriatrics, Uppsala University, Uppsala, Sweden

206 Epidemiology and Prevention Group, Center for Public Health Sciences, National Cancer Center, Tokyo, Japan

207 Department of Internal Medicine and the Center for Clinical and Translational Science, The Ohio State University, Columbus, OH, USA

208 Institute of Neuroscience and Physiology, the Sahlgrenska Academy at University of Gothenburg, Goteborg, Sweden

209 Department of Basic and Clinical Neurosciences, King's College London, London, UK

210 Department of Health Care Administration and Management, Graduate School of Medical Sciences, Kyushu University, Japan

211 Department of Medicine and Clinical Science, Graduate School of Medical Sciences, Kyushu University, Japan

212 Landspitali National University Hospital, Departments of Neurology & Radiology, Reykjavik, Iceland

213 Department of Neurology, Heidelberg University Hospital, Germany

214 Department of Neurology, Erasmus University Medical Center

215 Hospital Universitari Mutua Terrassa, Terrassa (Barcelona), Spain

216 Albert Einstein College of Medicine, Montefiore Medical Center, New York, NY, USA

217 John Hunter Hospital, Hunter Medical Research Institute and University of Newcastle, Newcastle, NSW, Australia

218 Centre for Prevention of Stroke and Dementia, Nuffield Department of Clinical Neurosciences, University of Oxford, UK

219 Department of Medical Sciences, Uppsala University, Uppsala, Sweden

220 Genetic and Genomic Epidemiology Unit, Wellcome Trust Centre for Human Genetics, University of Oxford, Oxford, UK

221 The Wellcome Trust Centre for Human Genetics, Oxford, UK

222 Beth Israel Deaconess Medical Center, Boston, MA, USA

223 Wake Forest School of Medicine, Wake Forest, NC, USA

224 Department of Neurology, University of Pittsburgh, Pittsburgh, PA, USA

225 BioBank Japan, Laboratory of Clinical Sequencing, Department of Computational biology and medical Sciences, Graduate school of Frontier Sciences, The University of Tokyo, Tokyo, Japan

226 Neurovascular Research Laboratory, Vall d'Hebron Institut of Research, Neurology and Medicine Departments-Universitat Autònoma de Barcelona. Vall d’Hebrón Hospital, Barcelona, Spain

227 Department of Biostatistics, University of Liverpool, Liverpool, UK

228 Wellcome Trust Centre for Human Genetics, University of Oxford, Oxford, UK

229 Institute of Genetic Epidemiology, Helmholtz Zentrum München - German Research Center for Environmental Health, Neuherberg, Germany

230 Department of Medicine I, Ludwig-Maximilians-Universität, Munich, Germany

231 DZHK (German Centre for Cardiovascular Research), partner site Munich Heart Alliance, Munich, Germany

232 Department of Cerebrovascular Diseases, Fondazione IRCCS Istituto Neurologico “Carlo Besta”, Milano, Italy

233 Karolinska Institutet, MEB, Stockholm, Sweden

234 University of Tartu, Estonian Genome Center, Tartu, Estonia, Tartu, Estonia

235 Department of Clinical and Experimental Sciences, Neurology Clinic, University of Brescia, Italy

236 Translational Genomics Unit, Department of Oncology, IRCCS Istituto di Ricerche Farmacologiche Mario Negri, Milano, Italy

237 Department of Genetics, Microbiology and Statistics, University of Barcelona, Barcelona, Spain

238 Psychiatric Genetics Unit, Group of Psychiatry, Mental Health and Addictions, Vall d’Hebron Research Institute (VHIR), Universitat Autònoma de Barcelona, Biomedical Network Research Centre on Mental Health (CIBERSAM), Barcelona, Spain

239 Department of Neurology, IMIM-Hospital del Mar, and Universitat Autònoma de Barcelona, Spain

240 IMIM (Hospital del Mar Medical Research Institute), Barcelona, Spain

241 National Institute for Health Research Comprehensive Biomedical Research Centre, Guy's & St. Thomas' NHS Foundation Trust and King's College London, London, UK

242 Division of Health and Social Care Research, King's College London, London, UK

243 FIMM-Institute for Molecular Medicine Finland, Helsinki, Finland

244 THL-National Institute for Health and Welfare, Helsinki, Finland

245 Iwate Tohoku Medical Megabank Organization, Iwate Medical University, Iwate, Japan

246 BHF Glasgow Cardiovascular Research Centre, Faculty of Medicine, Glasgow, UK

247 deCODE Genetics/Amgen, Inc., Reykjavik, Iceland

248 Icelandic Heart Association, Reykjavik, Iceland

249 Institute of Biomedicine, the Sahlgrenska Academy at University of Gothenburg, Goteborg, Sweden

250 Department of Epidemiology, University of Maryland School of Medicine, Baltimore, MD, USA

251 Institute of Cardiovascular and Medical Sciences, Faculty of Medicine, University of Glasgow, Glasgow, UK

252 Chair of Genetic Epidemiology, IBE, Faculty of Medicine, LMU Munich, Germany

253 Division of Epidemiology and Prevention, Aichi Cancer Center Research Institute, Nagoya, Japan

254 Department of Epidemiology, Nagoya University Graduate School of Medicine, Nagoya, Japan

255 University Medicine Greifswald, Institute for Community Medicine, SHIP-KEF, Greifswald, Germany

256 Department of Neurology, Caen University Hospital, Caen, France

257 University of Caen Normandy, Caen, France

258 Department of Internal Medicine, Erasmus University Medical Center, Rotterdam, Netherlands

259 Landspitali University Hospital, Reykjavik, Iceland

260 Survey Research Center, University of Michigan, Ann Arbor, MI, USA

261 University of Virginia Department of Neurology, Charlottesville, VA, USA
